# Post-pandemic social contact patterns in the United Kingdom: the Reconnect survey

**DOI:** 10.1101/2025.08.13.25333584

**Authors:** Lucy Goodfellow, Billy J. Quilty, Kevin van Zandvoort, W. John Edmunds

## Abstract

Close-contact and respiratory infectious diseases are spread through social interactions, which were affected by the COVID-19 pandemic and wider demographic and cultural changes. To estimate post-pandemic social contact patterns in the United Kingdom, we conducted a cross-sectional survey of 13,238 participants from November 2024 to March 2025. We calculated the mean number of daily contacts and contact matrices stratified by age, ethnicity, and socioeconomic status (SES). The mean number of daily contacts was 9.11 (95% confidence interval (CI): 8.73 - 9.50). Age-assortativity was high, while assortativity between ethnic groups was strongest in the home, and between SES groups in the workplace. In a novel respiratory pathogen outbreak, we found 2.29 times higher infection risk for Black people compared to White (95% CI: 2.08-2.55). This study provides crucial data to inform post-pandemic models of infectious disease transmission, and incorporate ethnicity and SES into such models.

## Introduction

The control of pathogens which spread through close contact and respiratory routes, such as influenza, respiratory syncytial virus, and SARS-CoV-2, is imperative for public health. These viruses are transmitted via social interactions, with strong correlations between measured social contacts and pathogen transmission [1–3]. The accuracy of mathematical models of infectious disease transmission has been transformed over the last two decades by the inclusion of data on social contacts. However, many of the studies which provide such data were performed before the COVID-19 pandemic (e.g. [4–6]). Many countries, including the UK, have experienced large-scale shifts in work and transport patterns since the pandemic, which would be expected to alter social contacts [7,8]. Furthermore, there is an evidence gap around social contact patterns within and between socioeconomic and ethnic groups, which limits the ability of modelling studies to investigate the impact of interventions on the distribution of diseases across these groups [9,10]. While the importance of this social structuring is well-recognised, its precise impact on epidemic potential is often difficult to quantify due to a lack of sufficiently detailed data capturing contacts between these groups. We conducted the Reconnect survey to address these gaps in empirical knowledge, by collecting contemporary data on social interactions across a representative sample of the UK population. We provide updated social contact matrices stratified by age, ethnicity, and socioeconomic status (SES), for the parameterisation of transmission models and study of infectious disease inequalities. Using these matrices, we further quantified infection risk of a novel close-contact pathogen in a completely susceptible population.

## Results

### Survey participants

We recruited 13,238 participants, who recorded the characteristics of 50,665 individual and 75,006 large group contacts over a 24-hour period; 11,303 of these participants were recruited in the first study period, and 1,935 in the boost period. 3,019 (22.8%) of our study population were aged under 18 years old, 501 of whom were aged under 5 years old. The median age of the study participants was 38 years old. 54.3% of the study sample was female, 45.5% male, and 0.1% ‘other’. 78.9% of study participants identified their ethnic group as White, 9.5% Black/African/Caribbean/Black British, 8.4% Asian/Asian British, 2.5% Mixed/multiple ethnic groups, 0.6% Other ethnic group, and 0.2% responded with ‘Prefer not to say’. The age breakdown of each ethnicity within the study sample is shown in Figure S2, with comparison to data from ONS data on ethnic group-specific age structure. We found a 70% dropoff rate between the demographic survey and the contact survey the following day.

37.9% of the study sample were employed full-time (2.9% self-employed), and 13.7% were employed part-time (1.9% self-employed), while 13% were retired. In comparison, 54.3% of contacts were recorded as employed, 18.2% as not employed (including retired individuals), 20.2% as students, and 7.4% recorded as ‘unknown’. Employed study participants slightly overrepresented National Statistics Socioeconomic Classification (NS-SEC) classes 1-3, and underrepresented classes 4-7. Further breakdowns of the study sample demography, alongside population proportions from the Office for National Statistics (ONS), are in Table 1.

**Table 1:** Study sample breakdown. Shown by various factors, with comparison to population-level data from the Office for National Statistics (ONS). Some ONS estimates are only calculated using Census 2021 data from England and Wales (ethnicity, highest qualification, employment status, household members, NS-SEC classes), and urban/rural proportions are calculated using data from England only; urban/rural survey proportions exclude participants whose urbanicity was not known.

| Variable | Category | % (N) | ONS % |
| --- | --- | --- | --- |
| Total |  | 13 238 | - |
| Adult/child | Adult | 77.2% (10 219) | 79.4% |
| Adult/child | Child | 22.8% (3019) | 20.6% |
| Age | 0-4 | 3.8% (501) | 5.30% |
| Age | 5-9 | 7.0% (931) | 5.8% |
| Age | 10-14 | 7.4% (977) | 6.1% |
| Age | 15-19 | 6.5% (855) | 5.8% |
| Age | 20-24 | 5.2% (694) | 6% |
| Age | 25-29 | 7.0% (932) | 6.5% |
| Age | 30-34 | 7.6% (1001) | 6.9% |
| Age | 35-39 | 8.1% (1070) | 6.7% |
| Age | 40-44 | 7.0% (932) | 6.4% |
| Age | 45-49 | 7.6% (1005) | 6.1% |
| Age | 50-54 | 7.1% (935) | 6.8% |
| Age | 55-59 | 6.4% (853) | 6.8% |
| Age | 60-64 | 6.1% (804) | 6% |
| Age | 65-69 | 4.7% (620) | 5.1% |
| Age | 70-74 | 4.7% (627) | 4.7% |
| Age | 75+ | 3.8% (501) | 9.1% |
| Gender | Female | 54.3% (7194) | 51% |
| Gender | Male | 45.5% (6026) | 49% |
| Gender | Other | 0.1% (17) | - |
| Ethnicity | White | 78.9% (10439) | 81.7% |
| Ethnicity | Asian | 8.4% (1112) | 9.3% |
| Ethnicity | Black | 9.5% (1255) | 4% |
| Ethnicity | Mixed | 2.5% (333) | 2.9% |
| Ethnicity | Prefer not to say | 0.2% (21) | - |
| Ethnicity | Other | 0.6% (78) | 2.1% |
| Highest qualification | Apprenticeship | 1.9% (258) | 4.3% |
| Highest qualification | Level 1 (1-4 GCSEs, O-levels (any), NVQ level 1, etc.) | 8.0% (1056) | 7.9% |
| Highest qualification | Level 2 (5+ GCSEs, O-levels (passes), NVQ level 2, etc.) | 10.9% (1444) | 10.9% |
| Highest qualification | Level 3 (A-level, BTEC, NVQ level 3, etc.) | 12.2% (1611) | 13.8% |
| Highest qualification | Level 4+ (University degree and above) | 39.0% (5165) | 27.5% |
| Highest qualification | No qualifications | 4.9% (652) | 14.8% |
| Highest qualification | Other | 0.2% (33) | 2.2% |
| Employment status | Employed full-time (35+ hours per week) | 35.0% (4631) | 38.8% |
| Employment status | Employed part-time | 11.8% (1565) | - |
| Employment status | Self-employed full time | 2.9% (390) | 7.8% |
| Employment status | Self-employed part time | 1.9% (250) | - |
| Employment status | Long-term sick or disabled | 2.5% (335) | 3.4% |
| Employment status | Looking after home or family | 3.0% (395) | 3.9% |
| Employment status | Retired | 13.0% (1722) | 17.6% |
| Employment status | Student | 2.5% (336) | 4.6% |
| Employment status | Unemployed (currently looking for work) | 2.8% (370) | 2.8% |
| Employment status | Unemployed (not currently looking for work) | 1.4% (180) | 2.6% |
| Employment status | Other | 0.3% (45) | - |
| Country | England | 85.6% (113 28) | 84.5% |
| Country | Northern Ireland | 2.1% (280) | 2.8% |
| Country | Scotland | 7.6% (1007) | 8% |
| Country | Wales | 4.7% (6 23) | 4.6% |
| Household members | 1 | 14.5% (19 22) | 28.9% |
| Household members | 2 | 25.8% (3 409) | 20.3% |
| Household members | 3 | 22.9% (3 030) | 21.8% |
| Household members | 4 | 23.8% (3 147) | 9.5% |
| Household members | 5 | 9.3% (1 23 4) | 3.8% |
| Household members | 6 | 2.7% (3 62) | 1.6% |
| Household members | 7 | 0.7% (9 2) | 1.3% |
| Household members | 8+ | 0.3% (40) | 1.3% |
| NS-SEC class (employed participants) | 1 (Higher managerial, administrative and professional occupations) | 26.0% (1731) | 15.6% |
| NS-SEC class (employed participants) | 2 (Lower managerial, administrative and professional occupations) | 27.9% (1857) | 23.8% |
| NS-SEC class (employed participants) | 3 (Intermediate occupations) | 18.2% (1 213) | 13.6% |
| NS-SEC class (employed participants) | 4 (Small employers and own account workers) | 5.6% (373) | 12.6% |
| NS-SEC class (employed participants) | 5 (Lower supervisory and technical occupations) | 3.7% (248) | 6.4% |
| NS-SEC class (employed participants) | 6 (Semi-routine occupations) | 10.5% (697) | 13.6% |
| NS-SEC class (employed participants) | 7 (Routine occupations) | 8.0% (533) | 14.4% |
| Urban/rural | Rural | 12.9% (1607) | 16.9% |
| Urban/rural | Urban | 87.1% (10895) | 83.1% |

### Mean contacts

We fitted negative binomial regression models using maximum-likelihood estimation (MLE) to calculate the mean number of daily contacts across various stratifications of the survey population. We compared the overall mean number of daily contacts to that found by the POLYMOD survey, which collected data from 1,012 participants in the UK in 2005-06 [4], and the final CoMix surveys conducted after the acute phase of the COVID-19 pandemic between November - December 2022 when there were no formal restrictions in place on social contacts [7].

The mean number of daily contacts was 9.11 (95% CI: 8.73 - 9.50) (Figure 1). This is significantly higher than the mean number of contacts found by Jarvis et al. at the end of the COVID-19 pandemic (6.5; 95% CI: 6.0 - 7.0) [7], but significantly lower than the mean number of daily contacts found by the POLYMOD survey in 2005-06 (11.74) [4]. We found that the data was overdispersed, therefore justifying the negative binomial assumption over a Poisson distribution, with a mean dispersion parameter estimated via MLE of 1.26 (95% CI: 1.20 - 1.33), where high values indicate greater heterogeneity and low values approximate a Poisson distribution. This is greater than the overdispersion value of 0.36 found by the POLYMOD survey, and below the value of 1.72 found by Jarvis et al. in 2022 [4,7]. We found that 6% of contacts occurred in multiple settings (home/work/school/other).

**Figure 1:**
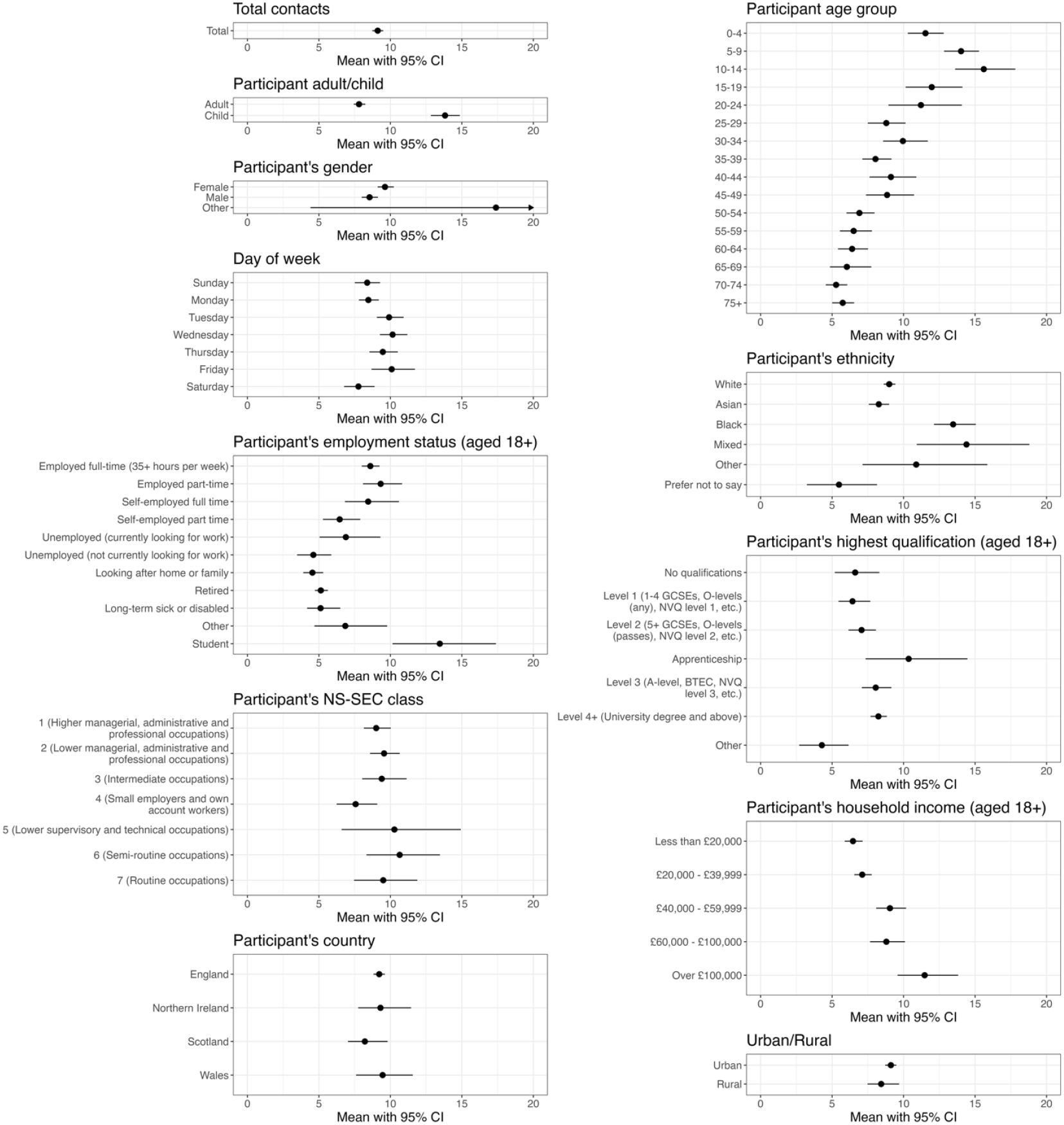
Mean daily number of social contacts and associated 95% confidence intervals. Shown across various stratifications, estimated using a negative binomial regression model weighted by participants’ age group, gender, ethnicity, and weekday/weekend survey day. Models for employment status were not weighted by age, and models for country were not weighted by ethnicity.

On average, children had 13.82 (95% CI: 12.83 - 14.86) daily contacts, compared to 7.81 (95% CI: 7.44 - 8.25) for adults. The number of daily contacts increased with age until 10-14 years old, and generally decreased thereafter (Fig. 1). The daily number of contacts differed across ethnicities: 9.00 (95% CI: 8.61 - 9.46) for participants identifying as White, 13.47 (95% CI: 12.13 - 15.05) for those identifying as Black/African/Caribbean/Black British, 8.27 (95% CI: 7.57 - 8.98) for Asian/Asian British, 14.40 (95% CI: 10.92 - 18.80) for Mixed/multiple ethnic groups, and 10.88 (95% CI: 7.13 - 15.86) for Other ethnic groups. While the age structure of each ethnicity differs widely (Fig. S1), differences in recorded contacts by ethnic group are maintained when standardising by age (Fig. S2).

Contact levels typically increased with participants’ household income and highest level of qualification, with the exception of those with an apprenticeship (Fig. 1). Employed (full- or part-time) or self-employed (full time) individuals had the greatest number of daily contacts after students; while individuals who were long-term sick or disabled, retired, unemployed and not looking for work, or looking after home or family, the fewest. There was no clear trend in contact levels when stratified by employed participants’ NS-SeC class. Mean daily contacts typically increased with household size: individuals living alone reported 5.54 (95% CI: 5.10 - 6.01) contacts on average, compared to 12.81 (95% CI: 8.22 - 18.60) for individuals in households of eight or more people (Table S7).

The mean number of daily recorded contacts varied across regions of England, from 8.15 (95% CI: 6.69 - 9.80) in the North East to 10.63 (95% CI: 9.36 - 11.96) in the East of England, and across countries in the UK, from 8.23 (95% CI: 7.15 - 9.80) in Scotland to 9.46 (95% CI: 7.68 - 11.72) in Wales (Fig. S6-7). We did not find a significant difference in the mean number of daily contacts between participants living in urban and rural areas (Fig. 1).

Weekdays were associated with 14.8% more contacts for adults than weekends (95% CI: 8.4% - 20.7%), with corresponding figures of 18.9% (95% CI: 10.3% - 26.6%) for children. Contacts of longer duration or increased intensity were more likely to involve physical touch (Fig. 2). 75% of contacts occurring in the home involved physical touch, compared to 44% at school and 27% in the workplace. 70% of contacts made on a daily basis lasted over an hour, whereas 81% of contacts between people who have not met before lasted under 15 minutes.

**Figure 2:**
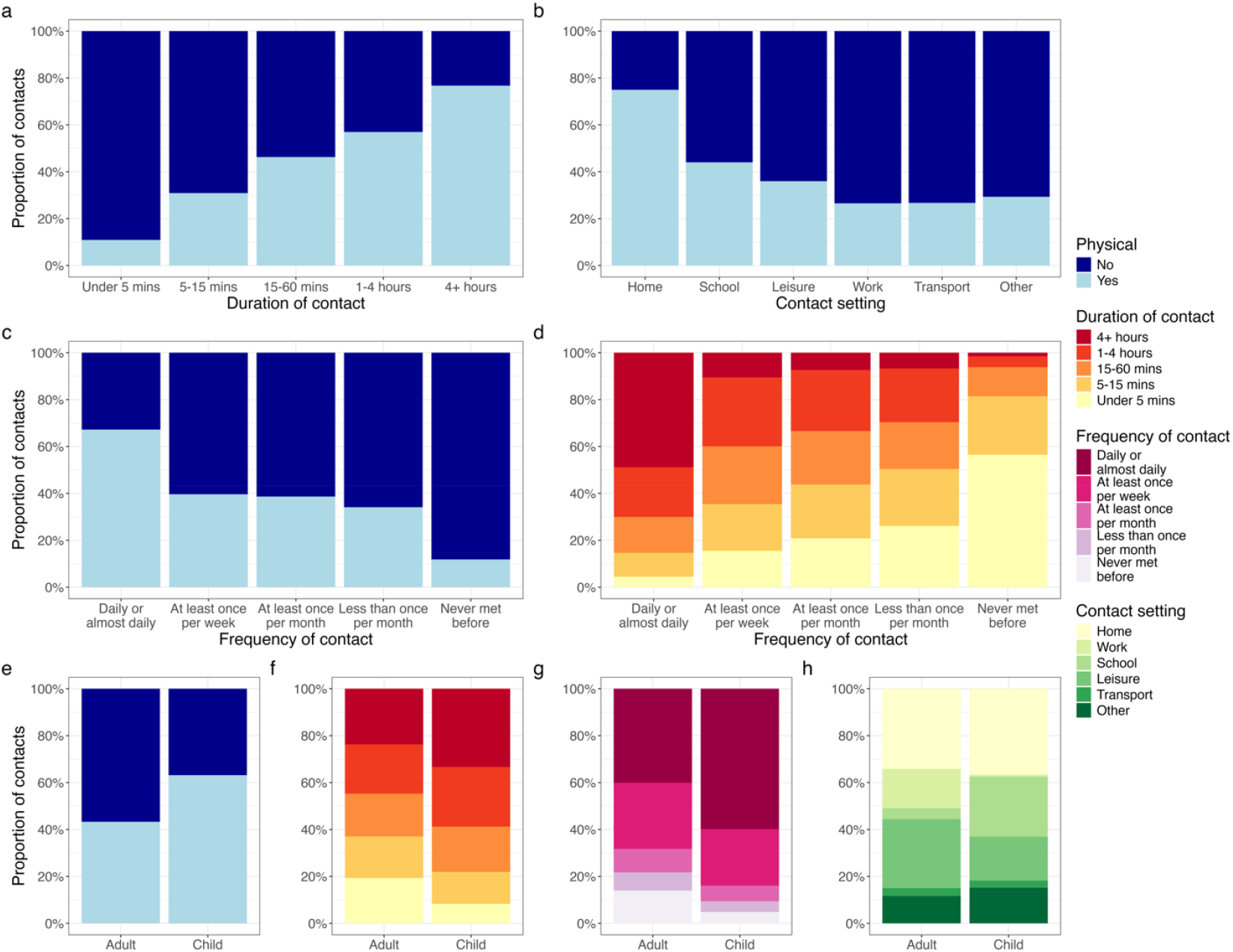
Breakdown of contact types. All proportions weighted by age group, gender, ethnicity, and weekday/weekend. **(A-C)** The proportion of contacts (excluding large group contacts) that involved physical contact, by duration, setting, and frequency of contact. **(D)** The correlation between duration and frequency of contact. **(E-H)** The proportion of contacts in adults and children split by physical contact, duration of contact, frequency of contact, and contact setting.

Children reported substantially more contacts during term time (mean daily contacts: 13.16; 95% CI: 12.43 - 13.93) compared to school holidays (9.32; 95% CI: 8.46 - 10.31). School holidays were associated with a 29.1% (95% CI: 20.7% - 36.9%) reduction in children’s mean number of contacts. We found similar changes in the mean number of contacts for the elderly but not adults, with relative reductions of 20.2% (95% CI: 6.1% - 31.7%) and 7.8% (95% CI: -8.8% - 23%), respectively (Fig. S3).

### Contact matrices

Assortativity of mixing was calculated using the measure of assortativity Q, as described by Keeling and Rohani [11], where Q = 1 describes complete assortativity, *Q* = 0 proportionate mixing, and *Q* < 0 disassortative mixing. Measures of assortativity among NS-SeC classes were restricted to interactions between employed individuals, i.e. NS-SEC classes 1-7, to reduce the impact of high assortativity between other groups captured by age groups (under 17s, retired individuals etc.).

Contacts were assortative by age group (*Q* = 0.14; 95% CI: 0.13 - 0.15), with assortativity strongest in children and in the school setting, and least pronounced in the workplace setting (Fig. 3, Fig. S10). The highest mean number of contacts between two age groups was between 10-14 year olds (6.71; 95% CI: 5.44 - 8.1). Full contact matrix data and confidence intervals, stratified by setting, can be found in Table S8. There are two parallel diagonals between children and adults, likely reflecting contact with their parents or guardians in the household (Figure 4b). When home contacts are broken down by gender, contacts are both age- and gender-assortative in younger ages, but more disassortative by gender in older ages (Fig. S11).

**Figure 3:**
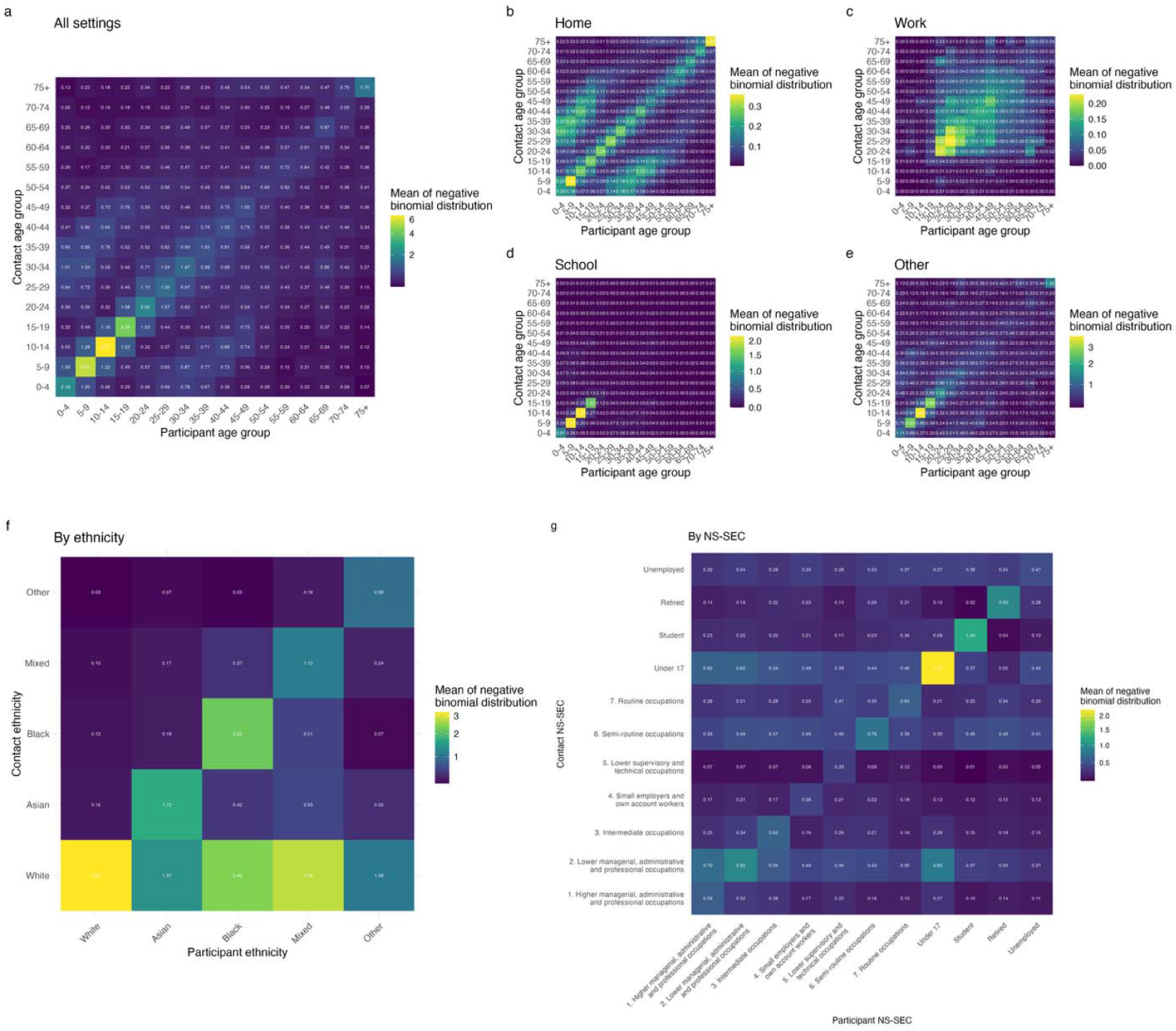
Stratified contact matrices. All contact matrices estimated by the weighted negative binomial regression model. **(A)** Mean number of contacts between individuals of each age group in all settings. **(B-E)** Mean number of contacts between individuals of each age group, stratified by setting. **(F)** Mean number of contacts between individuals of each ethnic group. **(G)** Mean number of contacts between individuals of each NS-SeC class or employment categorisation, not adjusted for population-level reciprocity.

**Figure 4:**
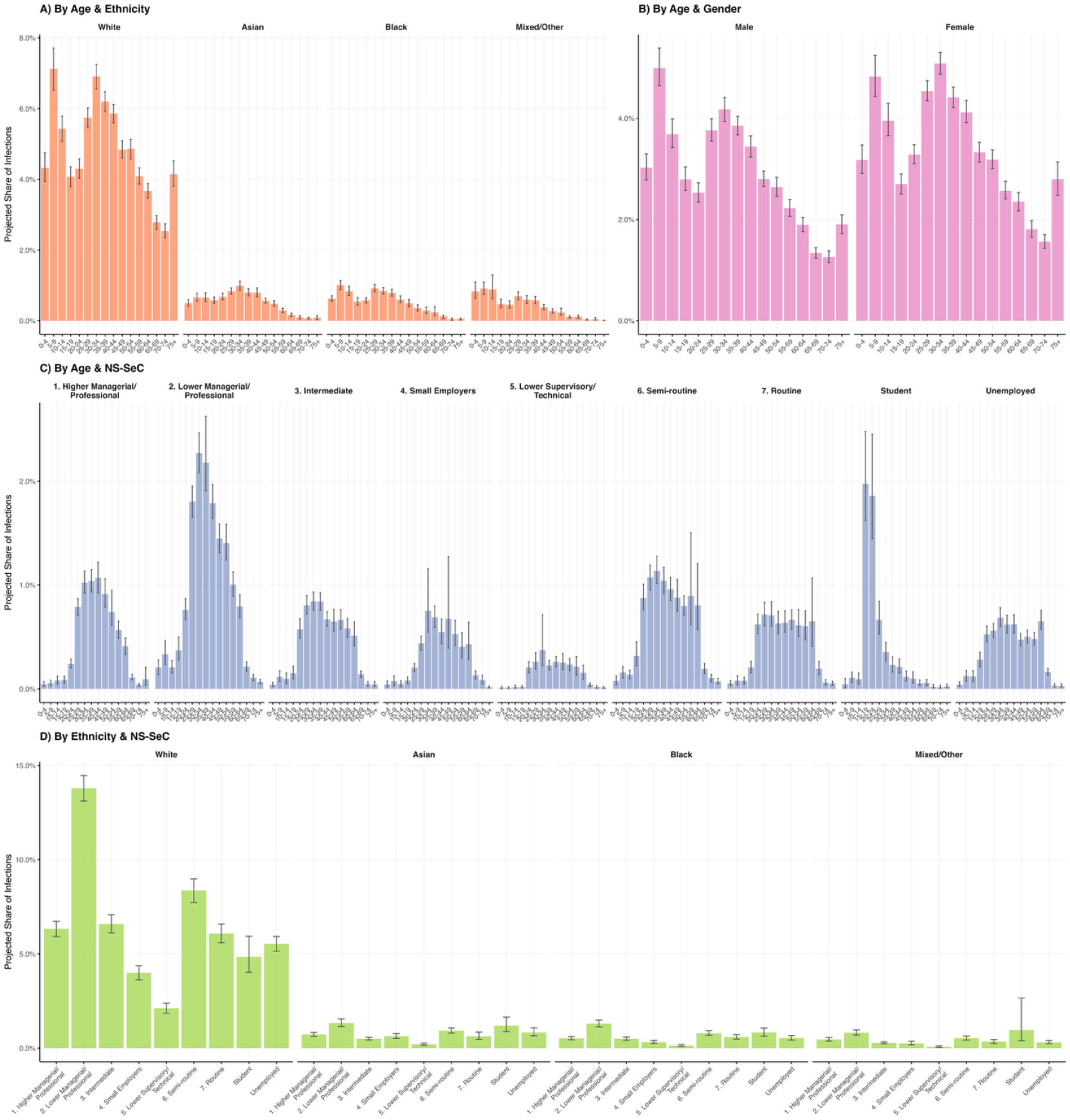
Mean projected share of infections by demographic groups, with 95% confidence intervals. **(A)** Age-ethnicity stratification. **(B)** Age-gender stratification. **(C)** Age-NS-SeC stratification, including all NS-SeC categories: working-age categories (1-7), retired, student, under 17, and unemployed. **(D)** Ethnicity-NS-SeC stratification (all ages, working-age categories (1-7), retired, student, under 17, and unemployed).

The data also displays assortativity between participants’ ethnicity and their contacts’ ethnicity (Q = 0.34; 95% CI: 0.32 - 0.37), as well as relatively high numbers of White contacts for participants of all ethnicities, driven by over 80% of the UK population identifying as White (Fig. 4f, Fig. S10). Assortativity of contacts by ethnicity is strongest in the home (Fig. S8, Fig. S10), but least pronounced in the workplace or school. In the workplace and ‘other’ settings, the majority of all participants’ contacts were White, regardless of the ethnicity of the survey participant. Interactions between NS-SeC classes showed moderate assortativity (*Q* = 0.14; 95% CI: 0.13 - 0.16); daily recorded contacts within NS-SeC classes were highest for class 2 (lower managerial, administrative and professional occupations) and class 6 (semi-routine occupations), respectively, and lowest for class 4 (small employers and own account workers). Assortativity within NS-SeC classes was strongest in the workplace, but less so in other settings (Fig. S9-10).

### Effect on infection risk

We projected the overall and per-capita share of infections of a novel close-contact pathogen in a completely susceptible population using socially-stratified next-generation matrices (NGMs). Analyses were conducted across one-way (age group, gender, ethnic group, and NS-SeC class) and two-way demographic stratifications (age group and ethnic group, age group and gender, age group and NS-SeC class, and ethnic group and NS-SeC class).

The largest projected share of infections occurred in the White ethnic group (76.1%, 95% CI: 74.8-77.4%), and N-SeC class 2: Lower managerial, administrative and professional occupations (14.7%, 95% CI: 14.2-15.3%) (Fig. 4). Relative infection risk was highest for the Black (2.29, 95% CI: 2.08-2.55) and Mixed/Other (1.42, 95% CI: 1.26-1.61) ethnic groups (compared to the White ethnic group as reference) and the Student (2.25, 95% CI: 1.96-2.60) and Unemployed (1.57, 95% CI: 1.45-1.68) NS-SeC classes (compared to NS-SeC class 1: Higher managerial and professional occupations as reference) (Fig. 5).

**Figure 5:**
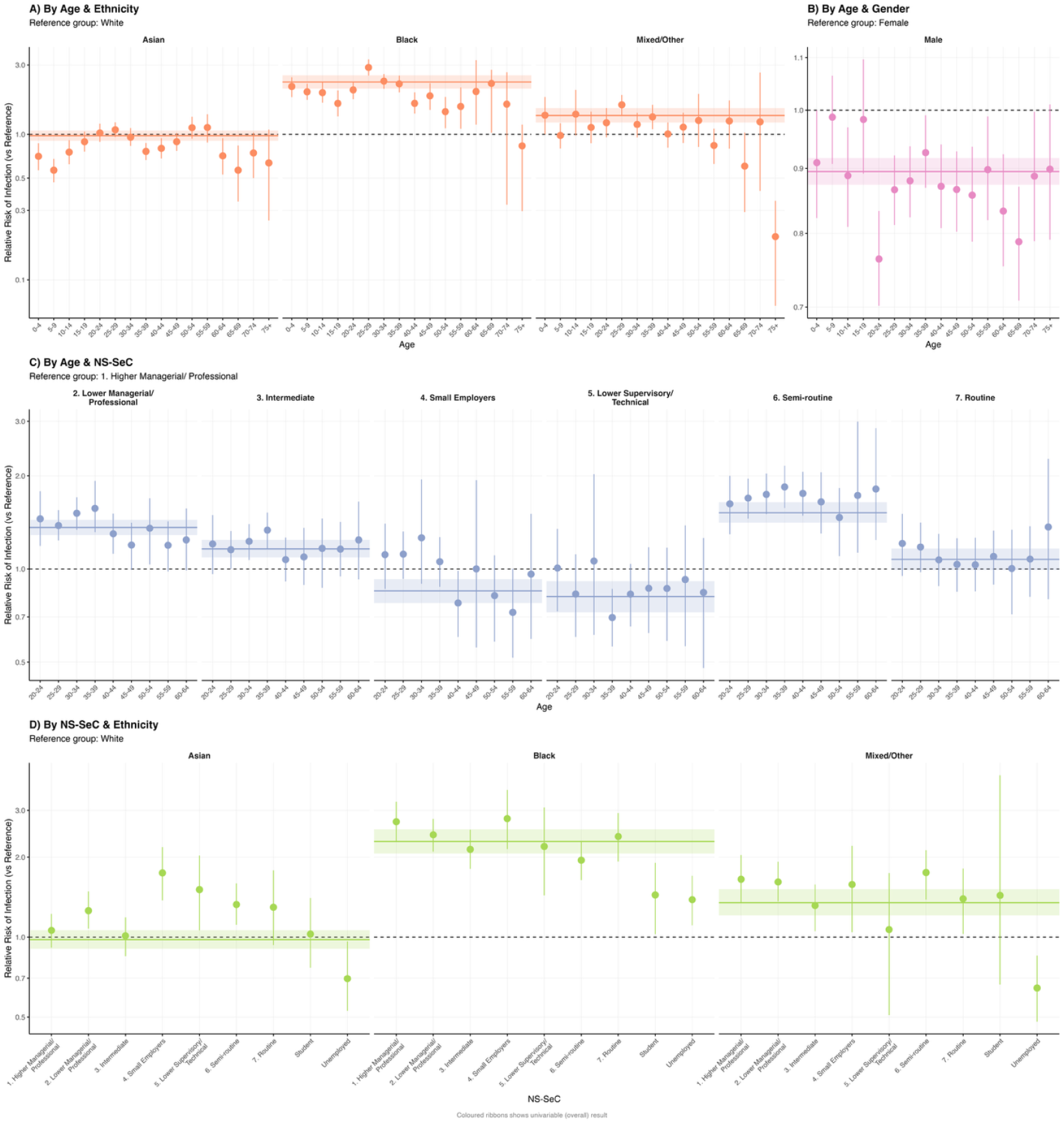
Mean relative risk of infection by demographic groups, with 95% confidence intervals. Relative risks shown on a log scale. Coloured ribbons show univariable mean results with 95% confidence intervals. Reference groups are not shown. **(A)** Age-ethnicity stratification. **(B)** Age-gender stratification. **(C)** Age-NS-SeC stratification, restricted to the working-age population (ages 20-64, NS-SeC categories 1-7). **(D)** Ethnicity-NS-SeC stratification, restricted to the working-age population (ages 20-64, NS-SeC categories 1-7).

In age-stratified analyses, younger and middle-aged groups had the largest projected share of infections across all demographic stratifications. The 5-9, White (7.1%, 95% CI: 6.6-7.7%) and 30-34, White (6.9%, 95% CI: 6.6-7.2%) groups were the largest contributors when stratified by age and ethnic group (Figure 4A). We found that the age-specific relative risk (RR) of infection across ethnic groups compared to the White ethnic group varied across age groups; for example, Black 25-29-year-olds were projected to experience a 2.88 (95% CI: 2.53-3.24) times elevated RR compared to their White counterparts, while the projected RR was 1.46 (95% CI: 1.15-1.86) for Black 50-54-year-olds (Fig. 5A). The same age-dependency in RRs was observed when comparing infection risk between genders (Fig. 5B). Minority ethnic groups were projected to experience higher relative infection risks than the White ethnic group within most NS-SeC categories (Fig. 5D). Further infection share and RR figures are in Table S9 and Supplementary File 2.

## Discussion

The accuracy and utility of mathematical models of infectious disease transmission depend on the availability of up to date, accurate, and specific data on pathogens and the populations through which they spread. Previously available social contact data do not necessarily reflect the impact of demographic changes and post-pandemic societal structures, and are limited in their ability to accurately describe interactions within and between socioeconomic and ethnic groups. Our study provides up-to-date post-pandemic data on social contact patterns in the UK with a large, representative survey sample across all ages, novel insights into the disparities in social involvement through differences in the mean number of daily contacts across age groups, ethnicities, and socioeconomic groups, and, to the best of our knowledge, the first data on mixing within and between ethnic and socioeconomic groups.

We found that reported contacts have decreased by approximately 20% since the POLYMOD survey was conducted in 2005-06, but increased by 40% since late 2022. Mean daily social contacts were greater than those found by POLYMOD in children aged under 5 years old, but lower for all ages above 5; the reduction in contacts was greatest for participants aged 30-50 years old. While contacts were reduced on both weekdays and weekends compared to POLYMOD, we found smaller differences in contact rates between weekdays and weekends for both children and adults than those found by POLYMOD, suggesting a decreased presence of weekday workplace and school contacts. These decreased contact levels suggest that there may be long-term shifts in the epidemiology of a range of close-contact infectious diseases, with corresponding effects on the effectiveness of public health and social measures as well as pharmaceutical interventions such as vaccination programmes. Our overdispersion results indicate that there is substantially greater heterogeneity in contact levels post-pandemic, but less heterogeneity than in 2022.

While the number of recorded social contacts was lower than those found by the POLYMOD survey, age-specific patterns largely followed similar distributions to POLYMOD and other contact surveys, with strong age-assortativity and inter-generational contacts observed. We found that age-assortative contacts in the home are more disassortative by gender in older ages, suggesting that the observed age-assortativity in the home is driven at older ages by heterosexual partnerships. We examined the proportion of contacts which involved physical contact across different settings, durations, and frequencies, which increases risk of transmission; the prevalence of physical contact was very similar to those found by the POLYMOD survey across all stratifications [4]. The proportion of contacts which occurred in multiple settings was also similar to that found by POLYMOD.

A key and novel finding of this study relates to contacts within and between ethnic groups and socioeconomic classes; by comparing assortativity by different attributes across contact settings, this study provides quantitative evidence of assortative mixing between ethnic groups and NS-SEC classes in all settings. While contacts were most assortative between ethnic groups in the home, and between socioeconomic classes in the workplace, these associations were not as strong in other settings. In the workplace or ‘other’ settings, the majority of participants’ contacts were White, regardless of participants’ ethnicity. We found trends in the mean number of daily contacts across socioeconomic gradients as defined by individuals’ highest level of qualification or household income, and typically higher contact levels for employed individuals than those not in work, but less consistent trends across NS-SeC classes.

Our analysis suggests that demographic groups projected to have the largest share of infections are not necessarily those with the highest per-capita risk. The largest share of infections was in from younger age groups and the White ethnic group (who make up 76.1% (95% CI: 74.8-77.4) of projected cases), while per-capita infection risk was highest in minority ethnic groups; we found that the Black ethnic group had a relative risk of infection 2.29 (95% CI: 2.08-2.55) times that of the White ethnic group (reference group). These results suggest that social structure creates epidemiological inequalities through differential contact patterns. The inequalities in exposure risk that our findings quantify are consistent with observations during the COVID-19 pandemic, where factors such as over-representation in public-facing occupations and residence in crowded, multi-generational households were significant drivers of infection risk for ethnic minority communities [12,13]. Our findings suggest that these risk disparities persist within NS-SeC strata, indicating more complex drivers beyond socioeconomic factors. The elevated exposure risk identified through contact patterns could be compounded by other vulnerabilities. Evidence from the pandemic showed that some communities facing higher exposure risk also had greater prevalence of comorbidities that worsened COVID-19 outcomes, and lower vaccination coverage [14–16]. The contact-driven risk we quantify may be one component of morbidity and mortality risk in epidemics and pandemics.

Our data collection methodology had some limitations. Contacts were defined as a contact involving a face-to-face conversation or physical touch. This definition does not necessarily capture all close proximity contacts, such as those in crowded public transport or elevators, or fomite transmission [17]. These transmission events remain difficult to capture through contact diaries. We used online contact diaries and allowed for the capture of large group contacts, similar to the CoMix survey, but in contrast to the POLYMOD survey which used paper-based contact diaries [7,18]. By inferring the age structure of large group contacts according to the setting-specific age distribution of contacts as found by POLYMOD, we implicitly assumed that the age distribution of contacts is similar to that found by the POLYMOD survey, although the magnitude of contacts is assumed to be different.

Self-reported contacts may have been subject to recall bias and brief interactions with strangers may have been forgotten by participants. We attempted to minimise the effect of recall bias by recruiting individuals one day ahead of the survey date, and prompting participants to write down their interactions through the survey day; 76% of children and 58% of adults reported recording contacts in a list throughout the day. It is also possible that reporting fatigue may have affected participants at higher rates than in the POLYMOD survey, as we asked participants to record more characteristics for each contact. Some of these characteristics may have been estimated by the participant, such as contacts’ age, ethnicity, and occupation, but aggregating information such as ages and ethnicities minimised the impact of estimation. Parents and guardians often filled out the survey on behalf of their child, or helped their child with the survey, potentially leading to lower accuracy for young age groups.

The data analysis was also subject to limitations. Analyses stratified by contacts’ characteristics were limited by the fact that we did not have information on characteristics other than setting and age group for large group contacts, restricting these analyses to individually-recorded contacts. Due to an error in data collection, data on physical interaction was missing for 4% of contacts, but we do not believe that this is significantly impactful on data analysis. In our analysis of social mixing within and between socioeconomic groups, we used the NS-SeC classification system. This represented the most pragmatic method of studying contact SES, but is only one definition of SES and does not encompass the wide range of factors which contribute to an individual’s SES. We found disparities in participants’ levels of social interaction by other relevant factors, such as highest level of qualification and household income, but these are difficult to feasibly evaluate for contacts without enrolling all possible contacts in the study. As our socioeconomic classification was based on occupation, it was not possible to analyse contacts across socioeconomic strata within non-working age groups: children and the elderly. Demographic group sizes were estimated using the Census 2021 data for England and Wales, which may not wholly represent the UK population [19]. Census data were not available at sufficient granularity for multi-variable demographic combinations due to anonymisation concerns, preventing full population weighting in multi-variable stratifications.

Our study provides novel data to improve and expand our quantitative understanding of infectious disease transmission, demonstrating disparities in contact levels between various social strata and quantifying mixing patterns by age groups, ethnicity, and socioeconomic classes. Social contact levels are lower than in 2005-06 but have increased since 2022. This suggests that sociocultural changes in the last two decades have led to lower levels of social interaction, but that as a society, we have returned to levels of interactions that more closely reflect pre-pandemic times than daily life three years ago. This study has also quantified, for the first time, variation in infection risk between ethnic and socioeconomic groups, and demonstrated increased infection risk in minority ethnic groups. These data can inform post-pandemic infectious disease models, which can incorporate ethnicity and SES, to improve understanding of infectious disease transmission and inequalities.

## Materials and Methods

### Study design

We conducted a cross-sectional social contact survey in the United Kingdom from 13th December 2024 to 10th February 2025. Participants were recruited from a UK-based internet panel, using quota sampling to ensure a close representation of the UK population in terms of gender, country, region (as defined by the Office for National Statistics (ONS), within England), household income, and education. We aimed to recruit at least 10,000 adult and 1,000 child participants, deliberately oversampling non-White participants and children in order to reduce data scarcity when conducting stratified analyses. As recruitment rates for children and individuals aged 70+ were lower than expected, we conducted a second round of the survey from 22nd February to 10th March 2025 to increase representation in these age groups.

Participants were invited to the study via email and, upon acceptance, were asked to complete a two-stage prospective diary-based survey using a web page accessible from desktop or mobile devices. They recorded demographic information about themselves and their households in the first stage, and were then instructed to document all contacts they would make on the following day (the ‘survey day’). Participants were encouraged to note down contacts as they occurred throughout the survey day to maximise recall. A contact was defined as someone a participant met in person and whom they spoke with or physically touched. At the end of the survey day, participants completed the second stage of the survey, in which they recorded detailed information for each person they had interacted with. Invitations were distributed evenly across all days of the week to capture representative contact patterns for weekdays and weekends. Participants received a small financial incentive for their participation.

### Data collection

For each participant, we collected comprehensive demographic information including age, gender, geographical location (first half of postcode), ethnicity, highest qualification, household income, occupation, housing status, household size, health-related quality of life (using EQ-5D), high-risk status for respiratory diseases, current symptoms, vaccine confidence, and recent public transport use.

For each contact reported, participants recorded the time and setting of the contact (from home, work, school, or other), duration of the contact, whether physical contact occurred, the estimated age, gender, ethnicity, and occupation of the individual contacted, their relationship to the participant, and frequency of typical interaction (days per week). For situations involving numerous contacts that would be difficult to record individually (e.g., large gatherings), participants had the option to record these as aggregate contacts at the end of the questionnaire, with broad estimation of those contact groups’ ages (under 18 years old, 18-64, or over 65), and setting (work, school, other).

### Sample size

We employed negative binomial regression models to estimate the mean number of daily contacts while accounting for heterogeneity between individuals. Sample size calculations indicated that with 10,000 participants, we would have substantial power to detect a 10% change in the total mean number of daily contacts compared to the POLYMOD survey [4], and the final CoMix surveys [7] (Supplementary Section 1).

### Ethics and data management

The Reconnect survey received ethical approval from the London School of Hygiene & Tropical Medicine Research Ethics Committee (LSHTM Ethics Ref: 30212). Informed consent was obtained from all participants before survey completion. Participants had the right to withdraw from the study at any time without providing a reason. All data were stored securely in compliance with the Data Protection Act, ensuring participant confidentiality. Anonymised data has been made publicly available to support broader research efforts.

### Analysis

#### Data preparation

Contact data was cleaned and categorised by setting and age groups of participants and their contacts. Participant and contact ages were aggregated into 16 groups: 0-4, 5-9, 10-14, …, 65-69, 70-74, 75+. We defined school holidays as 21st December 2024 to 5th January 2025, 15th to 23rd February 2025 for participants not living in Wales, and 22nd February - 2nd March 2025 for participants living in Wales; all other survey dates were labelled as ‘term time’. Due to an error in data collection, information on whether interactions involved physical contact were not recorded for 4% of contacts. These data were excluded when presenting physical contacts, but included otherwise. Where the same individual was contacted in multiple settings, we assigned their contact setting preferentially to ‘home’ if present, then ‘school’, then ‘work’, then ‘other’. Further details of data cleaning are in Supplementary Section 2. We truncated broad age- and setting-specific large group contacts at 300, to reduce the impact of outliers above 300; we also conducted a sensitivity analysis in which the total number of contacts was right-truncated at 100 (Supplementary Section 6b).

We determined participants’ SES using the Cascot (Computer Assisted Structured Coding Tool) programme [20]. All participants and contacts with job titles were categorised using the National Statistics Socioeconomic Classification (NS-SeC) as defined by the ONS, which is based on the Standard Occupational Classification 2020 (SOC 2020) and measures employment relations and the conditions of occupations [21]. This classification system aggregates occupations into seven analytic classes, from *Higher managerial, administrative, and professional occupations* to *Routine occupations*, and uses an eighth category for *Never worked and long-term unemployed*, which was not used in this analysis. The additional categories ‘Retired’, ‘Student’, ‘Under 17’, ‘Unemployed’, and ‘Unknown’ were used for participants and contacts without job titles, based on the information provided. Further details of our approach are detailed in Supplementary Section 3. We followed the 2021 Census in England and Wales to categorise 19 ethnic groups into five high-level ethnic groups, which we will hereon refer to as ‘Asian’, ‘Black’, ‘Mixed’, ‘White’, and ‘Other’.

#### Survey weighting

To adjust for potential sampling biases and ensure our sample is representative of the UK population, we applied post-stratification weights with respect to participants’ age group, gender, and ethnicity. These weights were based on the joint age-, gender-, and ethnicity-specific structure in England and Wales, as reported by the 2021 Census [22]. For those who reported their gender as other than male or female, or their ethnicity as ‘Prefer not to say’, no weighting by gender or ethnicity, respectively, was used. Additional weights were used with respect to participants’ survey day, to ensure that weekdays and weekends were represented proportionally. We truncated participants’ weights below and above the 5% and 95% quantiles, respectively, to reduce the impact of outliers with respect to representation.

Analyses of the mean number of contacts stratified by employment status were not weighted by age group due to large differences in the age structure of each strata (Table S5). For the same reason, geographically-stratified analyses (including participant’s country) were not weighted by participant ethnicity.

#### Mean contacts

We assumed that reported contacts follow a negative binomial distribution [4,7,23], and calculated the total and attribute-specific mean number of contacts and associated confidence intervals (CIs) using maximum-likelihood estimation (MLE) with 1000 bootstrap samples. For each bootstrap sample, participants were sampled with replacement according to the post-stratification weights, and a negative binomial model fitted to the total number of contacts (individually-reported and large group contacts) of the sampled participants. We then reported the mean maximum likelihood estimate for μ (the mean of the negative binomial distribution) and the associated 95% CI, for each of these analyses.

#### Contact matrices

We calculated contact matrices using the mean daily number of contacts between age groups, ethnic groups, and NS-SEC classes, using similar negative binomial MLE with 1000 bootstrap samples weighted by post-stratification weights. These matrices were stratified by setting: home, work, school, other, and all settings. All contact matrices were weighted with respect to weekday/weekend, while contacts between age groups were further weighted by gender and ethnicity, and contacts between ethnicities were weighted by gender and age group.

In the case of age-stratified contact matrices, we distributed participants’ large group contacts to age groups according to the setting-specific contact age distribution found by POLYMOD (adjusted for 2022 age structure) [4]; we chose not to use the individually recorded contact data from this survey as a sampling distribution as it is likely that these large group contacts systematically differ from those individually recorded. In other analyses, we excluded large group contacts. For contacts recorded with ethnicity ‘Prefer not to say’ by a participant of ethnicity *x* (0.93% of contacts), we imputed their ethnicity for the ethnicity-stratified matrix using the distribution of contacts’ ethnicities reported by participants of ethnicity *x*; we repeated this for the NS-SEC class-stratified matrix for contacts whose NS-SEC class was unknown (9.50% of contacts), using distributions based on their corresponding participant’s NS-SEC class. These imputations were sampled independently in each of the 1000 bootstrap samples used for the contact matrix fitting.

To ensure reciprocity in contacts, i.e. that the number of daily contacts from group *i* to group *j* is equal to the number of daily contacts from group *j* to group *i*, we normalised the age-stratified and ethnicity-stratified matrices with respect to the underlying population structure to produce reciprocal contact matrices, calculating each element by [4,18,23,24]:

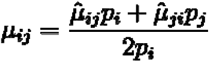

where 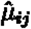 is the mean number of daily contacts from group *i* to group *j* in the unadjusted matrix, and *p*_*i*_ is the proportion of the population in group *i*, based on Census 2021 data [19]. We present the mean μ normalised values.

Assortativity of mixing was calculated using the measure of assortativity *Q*, as described by Keeling and Rohani [11], which was calculated using:

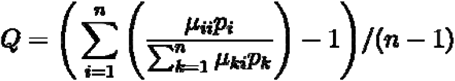

where *n* is the total number of groups in the population. *Q* = 1 describes complete assortativity, *Q* = 0 proportionate mixing, and *Q* < 0 disassortative mixing. We calculated assortativity in overall and setting-specific contact matrices stratified by age group, ethnicity, and NS-SEC class, where NS-SEC class was restricted to interactions between employed individuals, i.e. NS-SEC classes 1-7, to reduce the impact of high assortativity between other groups captured by age groups (under 17s, retired individuals etc.).

### The effect of social structure on infection risk

To calculate next generation matrices (NGMs), participants were weighted by weekday/weekend to ensure proportionate representation. For analyses involving multiple demographic stratification variables, only weekday/weekend weighting was applied as census data were not available at that level of granularity due to anonymisation concerns. Values for contacts with unknown ethnicity or NS-SeC class were imputed as above. For gender-specific analyses, we excluded participants and contacts with ‘Other’ gender. We aggregated the Mixed and Other ethnic groups for these analyses, to minimise the effect of small group sizes. Large group contacts were excluded, as ethnicity and NS-SeC data were not collected for these contacts.

The contact rate between groups *i* and *j* (μ_*ij*_) was adjusted for reciprocity using the same methods as described previously. The NGM *K*_*ij*_element was then defined as the expected number of new infections in group *j* caused by a single infectious individual from group *j* (*K* ∝ *C*^*T*^). We used this NGM *K* to derive the dominant right eigenvector (*w*), which represents the distribution of infections across the demographic strata, assuming uniform susceptibility and infectiousness within each stratum [4]. We defined the projected infection share *S*_*i*_ = *w*_*i*_, and calculated the per-capita infection risk for each demographic strata *i* as *S*_*i*_ /*p*_*i*_, *p*_*i*_ where is the population size based on Census 2021 data [19].

To compare the per-capita risk of infection across demographic strata, we calculated relative risk as *RR*_*i*_ = (*S*_*i*_/*P*_*i*_) / (*S*_*ref*_/*P*_*ref*_) against a reference group. Analyses were conducted using 1000 weighted bootstrap samples.

## Author contributions

Conceptualization: BJQ, WJE

Methodology: LG, BJQ, KvZ, WJE

Formal analysis: LG, BJQ

Writing - original draft: LG, BJQ

Writing - review & editing: LG, BJQ, KvZ, WJE

Visualization: LG, BJQ

Supervision: KvZ, WJE

Funding acquisition: WJE

## Data availability

The code and data used to conduct these analyses are found at https://github.com/cmmid/reconnect_uk_social_contact_survey and 10.5281/zenodo.16845075.

