## Supplementary Materials for "Post-pandemic social contact patterns in the United Kingdom: the Reconnect survey"

**Short title:** Post-pandemic social contact patterns in the UK

Lucy Goodfellow^1^ *^†^, Billy J. Quilty^1,2 †^, Kevin van Zandvoort^1‡^, W. John Edmunds^1‡^

^†^ equal contribution

^‡^ equal contribution

^1^ Centre for the Mathematical Modelling of Infectious Diseases, London School of Hygiene & Tropical Medicine, London, United Kingdom

^2^ Charité Centre for Global Health, Charité Universitätsmedizin Berlin, Berlin, Germany

### 1. Sample size calculation

We defined an appropriate minimum sample size for this study as a sample size such that a difference in overall mean contacts can be detected compared to previous surveys conducted in GB / the UK - POLYMOD (Mossong et al., fieldwork 2005-6) and CoMix (Jarvis et al., final survey Nov-Dec 2022). There were substantial differences in the overall mean daily contacts reported in each of these two surveys (11.74 and 6.5, respectively) with contributing factors being the lasting impacts of the COVID-19 pandemic and rise of widespread digital communication and social media. Thus, we predicted that overall contact rates will have decreased compared to POLYMOD, and increased compared to the final CoMix survey. As such, we specified a meaningful change as being at least a 10% decrease in contacts versus POLYMOD and at least a 10% increase versus the final CoMix survey. Another difference is that of the degree of overdispersion in daily contact rates; POLYMOD reported an overdispersion parameter (the reciprocal of the theta overdispersion parameter) of 0.36, whereas the last round of CoMix found much greater overdispersion (1.72); higher values indicating greater overdispersion.

With a sample size of at least 463 we would be be able to detect a 10% decrease with 80% power compared to the POLYMOD contact survey conducted in 2005-6 (Table 1), assuming overdispersion parameter of 2.78; a sample size of at least 3517 was required to detect a difference compared to the final CoMix survey in 2022, assuming an overdispersion parameter of 0.58.

The final sample size was subject to budget, though we aimed for over 10,000 individuals to reduce the risk of sparsity in the contact matrices. Quotas on age, gender, ethnicity, and geographic location were used to ensure that a representative sample of the UK population was reached and exceeded for minority ethnic groups.

| **Previous survey** | **Number of participants** | **Overall mean contacts** | **Relative change in number of reported contacts compared to previous survey** | **Overdispersion parameter** | **Sample size required to achieve power** |
| --- | --- | --- | --- | --- | --- |
| POLYMOD (2005-06) | 1012 | 11.74 | -10% | 0.36 | 463 |
| CoMix (final survey, Nov-Dec 2022) | 2991 | 6.5 | +10% | 1.72 | 3517 |

**Table S1:** Minimum sample sizes required to detect changes in overall contacts compared to previous surveys, given assumptions on magnitude of change and overdispersion.

### 2. Data cleaning

For any individuals who recorded their occupation as a teacher or other occupation which involved working in a school (e.g. Learning Support Assistant, Classroom Behaviour Support), we manually reallocated any contacts recorded as occurring at ‘work’ to ‘school’, so that all interactions occurring in schools were identified as such, even if that is an individual’s workplace too.

Some participants under age 18 recorded contacts in the workplace, which upon investigation was typically either their parent or guardian’s workplace, or a misallocated setting for interactions with siblings or school friends. We therefore manually reallocated the setting of any children’s contacts recorded as occurring in the workplace to ‘other’, unless the child was aged over 13 and reported having a job.

We removed five participants from the dataset whose recorded contacts were unfeasible, for example having many spouses under the age of 18, or repeatedly filling in open-ended questions with nonsensical text.

We truncated broad age- and setting-specific large group contacts at 300, to reduce the impact of outliers above 300. This led to changes in the reported large group contacts of 14 participants (Table S2).

| **p_age_group** | **add_u18_work** | **add_18_64_work** | **add_65_work** | **add_u18_school** | **add_18_64_school** | **add_65_school** | **add_u18_other** | **add_18_64_other** | **add_65_other** |
| --- | --- | --- | --- | --- | --- | --- | --- | --- | --- |
| 35-39 | 0 | 0 | 0 | 0 | 0 | 0 | 5 | 1000 | 100 |
| 15-19 | 0 | 0 | 0 | 200 | 800 | 30 | 20 | 30 | 5 |
| 65-69 | 0 | 0 | 0 | 0 | 0 | 0 | 10 | 1000 | 25 |
| 55-59 | 0 | 0 | 0 | 0 | 0 | 0 | 0 | 800 | 50 |
| 30-34 | 0 | 0 | 0 | 0 | 0 | 0 | 10 | 500 | 300 |
| 20-24 | 50 | 400 | 100 | 0 | 0 | 0 | 0 | 60 | 15 |
| 45-49 | 0 | 0 | 0 | 40 | 550 | 5 | 0 | 0 | 0 |
| 10-14 | 0 | 0 | 0 | 0 | 0 | 0 | 500 | 40 | 0 |
| 10-14 | 0 | 0 | 0 | 0 | 0 | 0 | 500 | 20 | 0 |
| 25-29 | 0 | 0 | 0 | 0 | 0 | 0 | 1 | 510 | 0 |
| 40-44 | 0 | 0 | 0 | 0 | 0 | 0 | 0 | 480 | 0 |
| 20-24 | 0 | 0 | 0 | 0 | 0 | 0 | 400 | 30 | 0 |
| 10-14 | 0 | 0 | 0 | 400 | 2 | 0 | 0 | 0 | 0 |
| 40-44 | 0 | 0 | 0 | 0 | 0 | 0 | 0 | 400 | 0 |

**Table S2:** Participants reporting over 300 contacts in one of the broad age- and setting-specific large group contact options, before truncation at 300, and participant age group.

In the survey, participants were asked their gender, whereas they were asked to report the sex of their contacts. Both questions had the options: ‘Male’, ‘Female’, ‘Other’, and ‘Prefer not to say’. We present results as ‘gender’ for both participants and contacts, for simplicity.

### 3. National Statistics Socioeconomic Classification (NS-SEC) derivations

To assign each participant and contact with a job title reported to an NS-SEC class, we first coded the free-text values of job title into four-digit categories of the Standard Occupational Classification 2020 (SOC 2020) using the Computer Assisted Structured Coding Tool (CASCOT) [20]. CASCOT is a software programme that assigns SOC 2020 codes to job titles with an associated certainty score (0-100), with the option of added qualifiers such as sectors. We used the following algorithm to consistently code job titles:

1. For each job title, attempt to classify using CASCOT with a certainty score of 60 or higher,
2. If unsuccessful, attach sector where possible as a qualifier:
   a. Occupational sector was recorded for survey participants,
   b. If a contact is recorded to be a colleague of the participant, the participant’s sector is attached,
3. Attempt to classify job with sector using CASCOT with a certainty score of 50 or higher,
4. Otherwise manually classify where possible.

These occupations were then converted into NS-SEC classes using the simplified derivation table (as size of organisation was unknown) [21]. We used the eight-class version of the NS-SEC (Table S3), excluding class 8 (never worked and long-term unemployed), and added classifications of those not employed based on survey responses. Self-employed individuals were either classified by their job title, when this resulted in NS-SEC analytic class 1 or 2, or coded as NS-SEC analytic class 4 in accordance with NS-SEC specifications. As we did not collect data on if participants’ contacts were retired, we attributed ‘Retired’ status to contacts recorded as ‘not employed’ who were aged over 65, or whose occupation was recorded as ‘Retired’.

| **NS-SEC Class** | **Number of employed survey participants in category (%)** | **Example job titles** |
| --- | --- | --- |
| 1. Higher managerial, administrative and professional occupations | 1731 (26.02%) | Hotel manager, Doctor, Auditor, CEO, Accountant, Solicitor, Marketing director |
| 2. Lower managerial, administrative and professional occupations | 1857 (27.92%) | Teacher, Staff nurse, Assistant manager, TV presenter, Private tutor, Social worker |
| 3. Intermediate occupations | 1213 (18.24%) | Library assistant, Civil servant, Administrator, Call handler, Ambulance crew |
| 4. Small employers and own account workers | 373 (5.61%) | Massage therapist, Interior designer, Driver, Plumber, Therapist, Hairdresser, Sports coach |
| 5. Lower supervisory and technical occupations | 248 (3.73%) | Chef, Mechanic, Grounds staff, Electrician, Train driver, Baker, Gardener |
| 6. Semi-routine occupations | 697 (10.48%) | Customer service assistant, Retail assistant, Housekeeper, Support worker, Cashier |
| 7. Routine occupations | 533 (8.01%) | HGV driver, Cleaner, Exam invigilator, Bar staff, Barista, Waitress |
| 8. Never worked and long-term unemployed | NA | NA |

**Table S3:** NS-SEC class titles, with the number and percentage of the employed survey population in each class, and example job titles for each class.

### 4. Urban/rural classifications

To assign urban/rural to participants based on the first half of their postcode, we merged the first half of postcodes, hereon called ‘pcd1’, with small area local-level measures (Lower layer Super Output Areas (LSOAs) in England and Wales, Data Zones in Scotland, and Small Areas in Northern Ireland), and combined with the most recent population estimates for each small area. We further appended the most recent urban/rural assignment of each small area, and totalled the urban and rural populations associated with each pcd1. This method therefore assigned a reasonable estimate of urban/rural status for all pcd1s:

1. Pcd1 contains or overlaps with multiple small areas
   a. The formula will assign the pcd1 the most populated urban/rural status of those small areas
2. Pcd1 is contained within a small area
   a. The formula will assign the pcd1 the urban/rural status of that small area

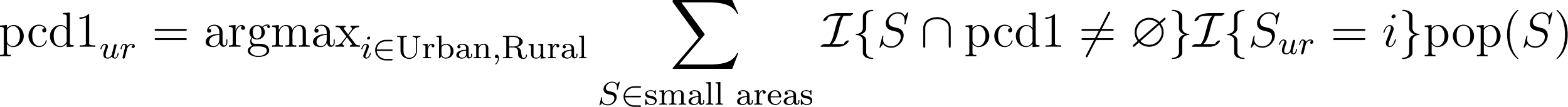
<https://www.codecogs.com/eqnedit.php?latex=%5Ctext%7Bpcd1%7D_%7Bur%7D%20%3D%20%5Ctext%7Bargmax%7D_%7Bi%20%5Cin%20%5Ctext%7BUrban%7D%2C%20%5Ctext%7BRural%7D%7D%20%5Csum_%7BS%20%5Cin%20%5Ctext%7Bsmall%20areas%7D%7D%20%5Cmathcal%7BI%7D%5C%7BS%20%5Ccap%20%5Ctext%7Bpcd1%7D%20%5Cneq%20%5Cvarnothing%20%5C%7D%20%20%5Cmathcal%7BI%7D%5C%7BS_%7Bur%7D%20%3D%20i%20%5C%7D%20%5Ctext%7Bpop%7D(S)#0>

Where the ‘ur’ subscript denotes urban or rural status.

We then assigned urban/rural to each participant based on their pcd1 input. In some cases participants had clearly either missed the final character off of the first half of their postcode (most commonly for London postcodes) or included the first character of the second half of their postcode; we manually corrected these issues. Overall, 12.1% of survey participants lived in rural areas, 82.3% in urban areas, and 5.6% did not provide the first half of their postcode or provided an unmatchable postcode.

### 5. Weights for stratified analyses

| **Stratifying attribute** | **Weighting variables** | | | |
| --- | --- | --- | --- | --- |
|  | **Age group** | **Ethnicity** | **Gender** | **Weekday/weekend** |
| **Total** | Yes | Yes | Yes | Yes |
| **Adult/child** | Yes | Yes | Yes | Yes |
| **Age group** | No | Yes | Yes | Yes |
| **Ethnicity** | Yes | No | Yes | Yes |
| **Gender** | Yes | Yes | No | Yes |
| **Day of week** | Yes | Yes | Yes | No |
| **Employment status** | No | Yes | Yes | Yes |
| **Highest qualification** | Yes | Yes | Yes | Yes |
| **Country** | Yes | No | Yes | Yes |
| **Housing status** | Yes | Yes | Yes | Yes |
| **Urban/Rural** | Yes | Yes | Yes | Yes |

**Table S4:** Variables used for weighting in each stratified analysis.

| **Participant’s employment status** | **Median age** |
| --- | --- |
| Child (Not Applic.) | 10 |
| Employed full-time (35+ hours per week) | 39 |
| Employed part-time | 45 |
| Long-term sick or disabled | 50 |
| Looking after home or family | 48 |
| Other | 55 |
| Retired | 70 |
| Self-employed full time | 46.5 |
| Self-employed part time | 55.5 |
| Student | 21 |
| Unemployed (currently looking for work) | 41 |
| Unemployed (not currently looking for work) | 42 |

**Table S5:** Median age in the survey sample, stratified by employment status.

### 6. Results

#### a. Study sample composition

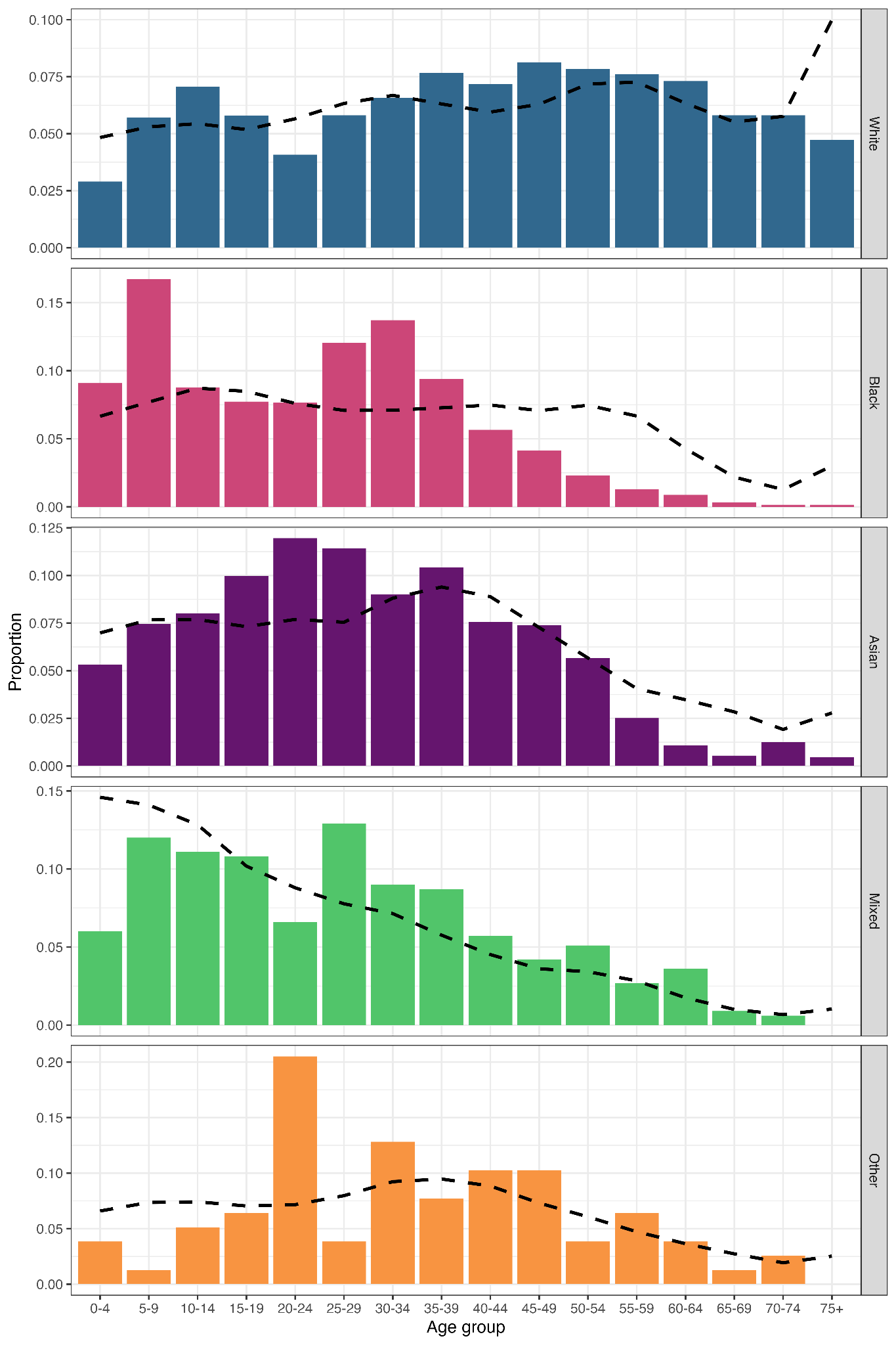

**Figure S1:** Proportion of the survey sample in each age group, stratified by ethnicity, compared to Census 2021 age- and ethnicity-specific population proportions (dotted lines). Census 2021 data is calculated from England and Wales populations only.

| **Contact Setting** | **Proportion** |
| --- | --- |
| Healthcare setting | 2.05% |
| Home | 35.04% |
| Hospitality venue | 4.76% |
| Other | 4.46% |
| Other house | 9.44% |
| Outside | 3.59% |
| Personal care establishment | 0.48% |
| Place of worship | 1.59% |
| Private transport | 0.92% |
| Public transport | 3.24% |
| Retail venue | 8.01% |
| School | 10.23% |
| Sports venue | 2.68% |
| Work | 13.49% |

**Table S6:** The proportion of recorded contacts (excluding large group contacts) taking place in each setting.

#### b. Mean contacts

| **Variable** | **Category** | **Mean weighted negative binomial regression estimate (95% UI)** | **Mean weighted negative binomial regression estimate with right-truncation at 100 (95% UI)** |
| --- | --- | --- | --- |
| Total contacts | Total | 9.11 (8.73, 9.5) | 8.31 (8.06, 8.53) |
| Participant adult/child | Adult | 7.81 (7.44, 8.25) | 7.12 (6.89, 7.34) |
| Participant adult/child | Child | 13.82 (12.83, 14.86) | 12.78 (12.15, 13.39) |
| Participant age group | 0-4 | 11.53 (10.3, 12.81) | 11.43 (10.27, 12.63) |
| Participant age group | 5-9 | 14.02 (12.83, 15.28) | 13.71 (12.66, 14.79) |
| Participant age group | 10-14 | 15.61 (13.61, 17.82) | 13.63 (12.39, 14.88) |
| Participant age group | 15-19 | 11.97 (10.14, 14.12) | 10.48 (9.49, 11.45) |
| Participant age group | 20-24 | 11.21 (8.94, 14.08) | 9.07 (7.88, 10.34) |
| Participant age group | 25-29 | 8.79 (7.5, 10.14) | 7.94 (7.22, 8.73) |
| Participant age group | 30-34 | 9.95 (8.57, 11.7) | 9.04 (8.17, 9.97) |
| Participant age group | 35-39 | 8.04 (7.11, 9.16) | 7.6 (6.84, 8.36) |
| Participant age group | 40-44 | 9.11 (7.62, 10.9) | 7.83 (6.97, 8.73) |
| Participant age group | 45-49 | 8.85 (7.37, 10.73) | 7.57 (6.68, 8.48) |
| Participant age group | 50-54 | 6.91 (6.01, 7.97) | 6.57 (5.92, 7.24) |
| Participant age group | 55-59 | 6.51 (5.55, 7.79) | 6.06 (5.4, 6.78) |
| Participant age group | 60-64 | 6.4 (5.42, 7.52) | 6.09 (5.31, 6.96) |
| Participant age group | 65-69 | 6.03 (4.85, 7.74) | 5.44 (4.71, 6.28) |
| Participant age group | 70-74 | 5.28 (4.56, 6.06) | 5.27 (4.61, 6.03) |
| Participant age group | 75+ | 5.74 (5, 6.54) | 5.73 (5.01, 6.59) |
| Participant's gender | Female | 9.62 (9.11, 10.24) | 8.8 (8.48, 9.13) |
| Participant's gender | Male | 8.54 (7.99, 9.13) | 7.77 (7.49, 8.1) |
| Participant's gender | Other | 17.39 (4.41, 35.77) | 15.13 (4.65, 29.53) |
| Participant's ethnicity | White | 9 (8.61, 9.43) | 8.14 (7.9, 8.4) |
| Participant's ethnicity | Asian | 8.27 (7.57, 8.98) | 8.21 (7.55, 8.93) |
| Participant's ethnicity | Black | 13.47 (12.13, 15.05) | 12.47 (11.56, 13.45) |
| Participant's ethnicity | Mixed | 14.4 (10.92, 18.8) | 11.72 (9.88, 13.68) |
| Participant's ethnicity | Prefer not to say | 5.48 (3.24, 8.14) | 5.48 (3.38, 8.05) |
| Participant's ethnicity | Other | 10.88 (7.13, 15.86) | 10.66 (7.22, 14.82) |
| Participant's employment status (aged 18+) | Employed full-time (35+ hours per week) | 8.6 (8, 9.24) | 7.84 (7.48, 8.19) |
| Participant's employment status (aged 18+) | Employed part-time | 9.32 (8.08, 10.82) | 8.01 (7.35, 8.69) |
| Participant's employment status (aged 18+) | Self-employed full time | 8.45 (6.82, 10.6) | 7.83 (6.68, 9.12) |
| Participant's employment status (aged 18+) | Self-employed part time | 6.45 (5.28, 7.89) | 6.4 (5.26, 7.68) |
| Participant's employment status (aged 18+) | Long-term sick or disabled | 5.12 (4.17, 6.5) | 4.9 (4.12, 5.78) |
| Participant's employment status (aged 18+) | Looking after home or family | 4.55 (3.91, 5.3) | 4.52 (3.95, 5.23) |
| Participant's employment status (aged 18+) | Retired | 5.13 (4.72, 5.63) | 5.06 (4.69, 5.46) |
| Participant's employment status (aged 18+) | Student | 13.45 (10.15, 17.39) | 11.15 (9.4, 13.15) |
| Participant's employment status (aged 18+) | Unemployed (currently looking for work) | 6.89 (5.05, 9.31) | 6.07 (4.89, 7.41) |
| Participant's employment status (aged 18+) | Unemployed (not currently looking for work) | 4.61 (3.47, 5.87) | 4.59 (3.5, 5.93) |
| Participant's employment status (aged 18+) | Other | 6.85 (4.69, 9.78) | 6.81 (4.51, 9.87) |
| Participant's highest qualification (aged 18+) | Apprenticeship | 10.35 (7.34, 14.47) | 9.14 (7.03, 11.38) |
| Participant's highest qualification (aged 18+) | Level 1 (1-4 GCSEs, O-levels (any), NVQ level 1, etc.) | 6.43 (5.45, 7.67) | 5.91 (5.3, 6.56) |
| Participant's highest qualification (aged 18+) | Level 2 (5+ GCSEs, O-levels (passes), NVQ level 2, etc.) | 7.06 (6.15, 8.07) | 6.49 (5.91, 7.11) |
| Participant's highest qualification (aged 18+) | Level 3 (A-level, BTEC, NVQ level 3, etc.) | 8.05 (7.08, 9.15) | 7.12 (6.61, 7.68) |
| Participant's highest qualification (aged 18+) | Level 4+ (University degree and above) | 8.24 (7.69, 8.83) | 7.57 (7.22, 7.92) |
| Participant's highest qualification (aged 18+) | No qualifications | 6.61 (5.19, 8.31) | 5.71 (4.85, 6.67) |
| Participant's highest qualification (aged 18+) | Other | 4.29 (2.7, 6.15) | 4.27 (2.73, 6.18) |
| Participant's country | England | 9.22 (8.82, 9.61) | 8.44 (8.2, 8.69) |
| Participant's country | Northern Ireland | 9.31 (7.75, 11.45) | 9.04 (7.68, 10.64) |
| Participant's country | Scotland | 8.22 (7.03, 9.81) | 7.68 (6.94, 8.44) |
| Participant's country | Wales | 9.46 (7.6, 11.57) | 8.27 (7.21, 9.45) |
| Participant's housing status (aged 18+) | Living rent free | 12.88 (8.74, 17.86) | 10.08 (7.97, 12.62) |
| Participant's housing status (aged 18+) | Other private rented | 9.53 (6.41, 13.28) | 9.6 (6.46, 13.55) |
| Participant's housing status (aged 18+) | Owned outright | 7.71 (7.06, 8.38) | 7.16 (6.77, 7.57) |
| Participant's housing status (aged 18+) | Owned with a mortgage or loan | 8.24 (7.55, 9.09) | 7.65 (7.2, 8.13) |
| Participant's housing status (aged 18+) | Rented from Council (Local Authority) | 6.54 (5.6, 7.66) | 6 (5.41, 6.68) |
| Participant's housing status (aged 18+) | Rented from a relative or friend of household member | 5.07 (4.22, 6) | 5.06 (4.2, 6.01) |
| Participant's housing status (aged 18+) | Rented from housing association, housing co-operative, charitable trust, or registered social landlord | 7.24 (5.85, 9.03) | 6.56 (5.65, 7.52) |
| Participant's housing status (aged 18+) | Rented from private landlord or letting agency | 7.79 (6.88, 8.9) | 6.93 (6.41, 7.45) |
| Participant's housing status (aged 18+) | Shared ownership - with a mortgage and paying rent | 9.06 (6.94, 12.05) | 8.56 (6.8, 10.66) |
| Participant's housing status (aged 18+) | Tied accommodation (accommodation provided by employer of a household member) | 8.59 (3.2, 18.9) | 8.19 (3.1, 17.05) |
| Participant's NS-SEC class | 1 (Higher managerial, administrative and professional occupations) | 9.01 (8.15, 10.03) | 8.31 (7.72, 8.97) |
| Participant's NS-SEC class | 2 (Lower managerial, administrative and professional occupations) | 9.56 (8.58, 10.65) | 8.6 (8.06, 9.19) |
| Participant's NS-SEC class | 3 (Intermediate occupations) | 9.4 (8.03, 11.14) | 8.13 (7.37, 9) |
| Participant's NS-SEC class | 4 (Small employers and own account workers) | 7.57 (6.24, 9.08) | 7.35 (6.09, 8.77) |
| Participant's NS-SEC class | 5 (Lower supervisory and technical occupations) | 10.28 (6.58, 14.92) | 7.98 (6.19, 9.86) |
| Participant's NS-SEC class | 6 (Semi-routine occupations) | 10.65 (8.33, 13.47) | 8.79 (7.72, 9.97) |
| Participant's NS-SEC class | 7 (Routine occupations) | 9.5 (7.46, 11.89) | 7.96 (6.76, 9.41) |
| Urban/Rural | Rural | 8.43 (7.48, 9.69) | 7.57 (7.02, 8.18) |
| Urban/Rural | Urban | 9.11 (8.71, 9.49) | 8.32 (8.08, 8.59) |
| Day of week | Monday | 8.46 (7.8, 9.19) | 8.09 (7.59, 8.62) |
| Day of week | Tuesday | 9.91 (9.05, 10.92) | 9.1 (8.51, 9.68) |
| Day of week | Thursday | 9.47 (8.53, 10.52) | 9.03 (8.29, 9.88) |
| Day of week | Wednesday | 10.15 (9.26, 11.19) | 9.27 (8.65, 9.95) |
| Day of week | Friday | 10.09 (8.68, 11.72) | 8.42 (7.73, 9.12) |
| Day of week | Saturday | 7.76 (6.77, 8.9) | 7.01 (6.51, 7.52) |
| Day of week | Sunday | 8.38 (7.51, 9.28) | 7.57 (6.99, 8.19) |
| Participant's household size | 1 | 5.54 (5.1, 6.01) | 5.47 (5.05, 5.89) |
| Participant's household size | 2 | 8.33 (7.58, 9.12) | 7.29 (6.86, 7.76) |
| Participant's household size | 3 | 8.88 (8.07, 9.85) | 7.98 (7.51, 8.48) |
| Participant's household size | 4 | 11.3 (10.42, 12.24) | 10.23 (9.7, 10.77) |
| Participant's household size | 5 | 11.04 (9.92, 12.33) | 10.26 (9.52, 11.04) |
| Participant's household size | 6 | 11.94 (9.91, 14.24) | 11.09 (9.62, 12.61) |
| Participant's household size | 7 | 16.69 (10.92, 25.19) | 14.06 (10.8, 18.1) |
| Participant's household size | 8+ | 12.81 (8.22, 18.6) | 12.81 (8.25, 18.12) |
| Participant's household income (aged 18+) | Less than £20,000 | 6.46 (5.88, 7.13) | 6.15 (5.64, 6.65) |
| Participant's household income (aged 18+) | £20,000 - £39,999 | 7.11 (6.55, 7.77) | 6.64 (6.28, 7.02) |
| Participant's household income (aged 18+) | £40,000 - £59,999 | 9.05 (8.08, 10.18) | 8 (7.45, 8.6) |
| Participant's household income (aged 18+) | £60,000 - £100,000 | 8.79 (7.65, 10.1) | 7.76 (7.16, 8.34) |
| Participant's household income (aged 18+) | Over £100,000 | 11.47 (9.58, 13.82) | 9.98 (8.92, 11.13) |

**Table S7:** Total mean number of daily contacts (95% confidence interval), stratified by various variables of interest, as calculated by the weighted negative binomial regression model, and with right-truncation at 100 contacts. Estimates are weighted by participants’ age group, gender, ethnicity, and weekday/weekend survey day, where not already stratified by these variables, except employment status (not weighted by age) and country (not weighted by ethnicity).

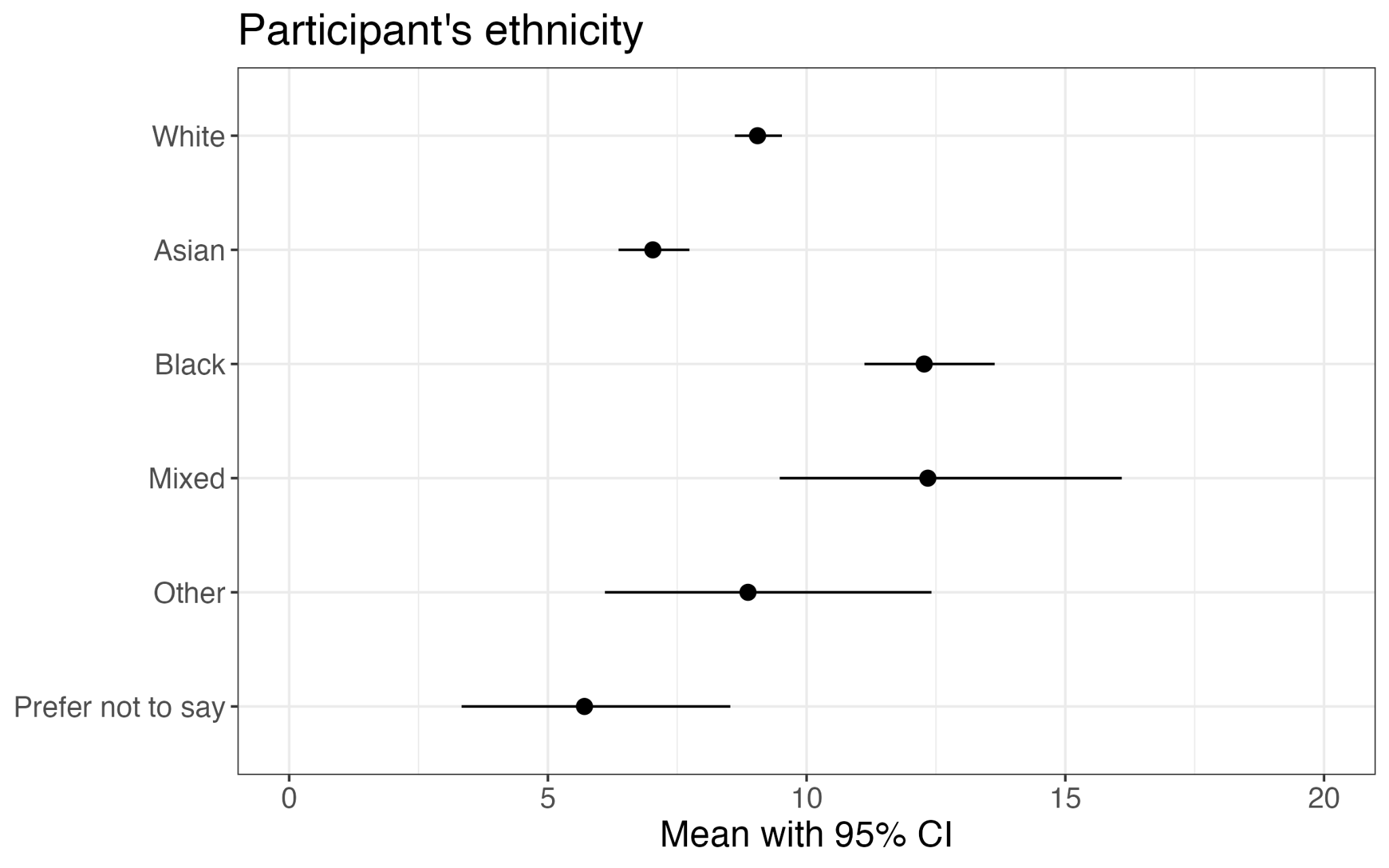

**Figure S2:** Age-standardised mean daily number of social contacts (with 95% confidence interval) across ethnic groups, calculated using the weighted negative binomial regression model. Estimates are weighted by participants’ age group, gender, and weekday/weekend survey day. Age group weighting is applied independently of participant ethnicity, to produce age-standardised results.

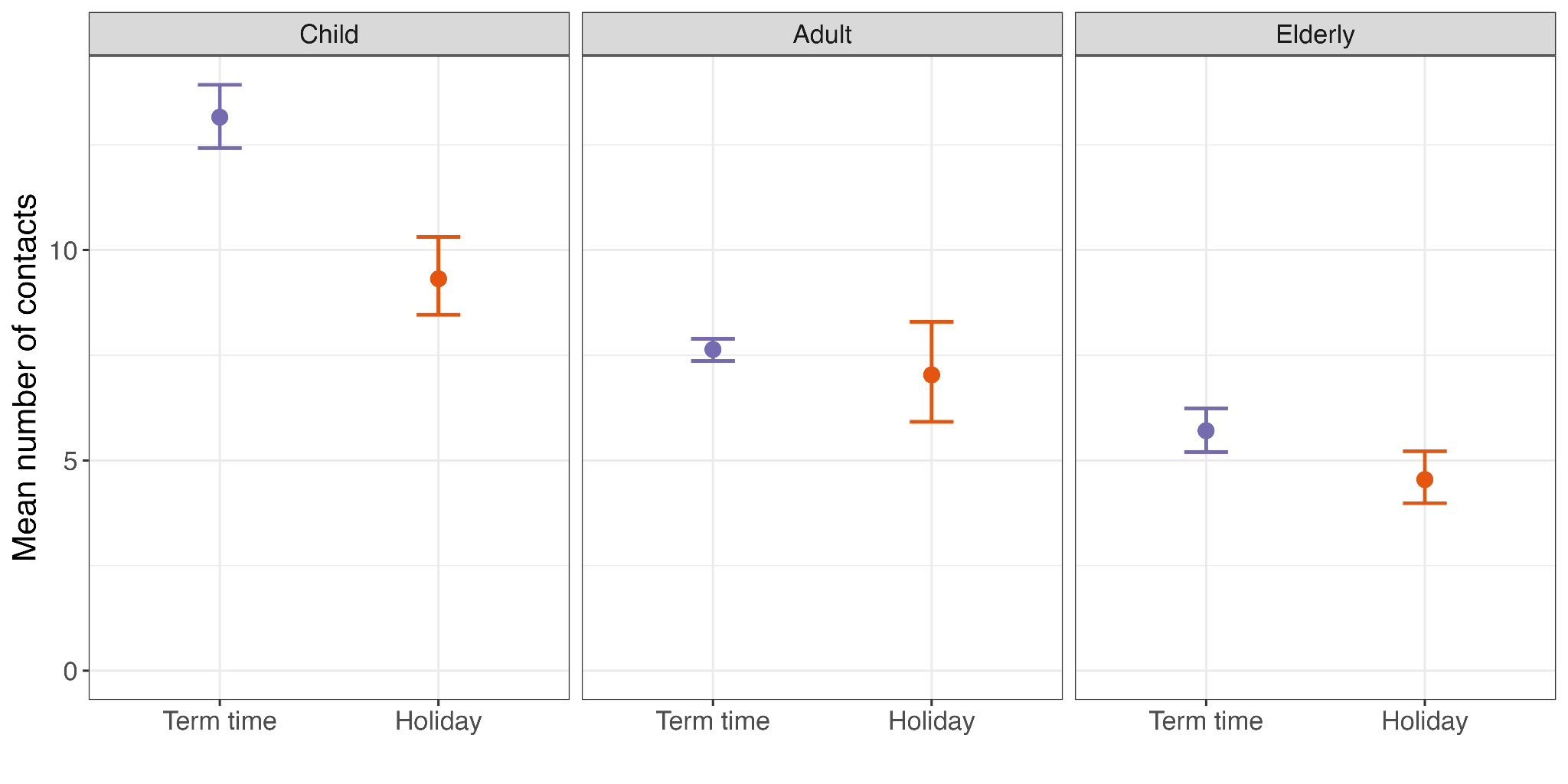

**Figure S3:** Mean daily number of contacts in term time and school holidays, shown for children (aged under 18), adults (aged 18-64), and elderly (aged 65+). All data were weighted by participant age group, gender, ethnicity, and weekday/weekend.

#### c. Age distribution of contacts compared to POLYMOD

We compared the setting-specific age distribution of contacts, stratified by participant age group, between the POLYMOD survey and this survey (weighted by participant gender, ethnicity, and weekday/weekend, and excluding large group contacts). Age distributions are largely similar, although the POLYMOD dataset is more subject to sparsity due to the smaller sample size (1,012 participants, compared to 13,238 participants in this survey).

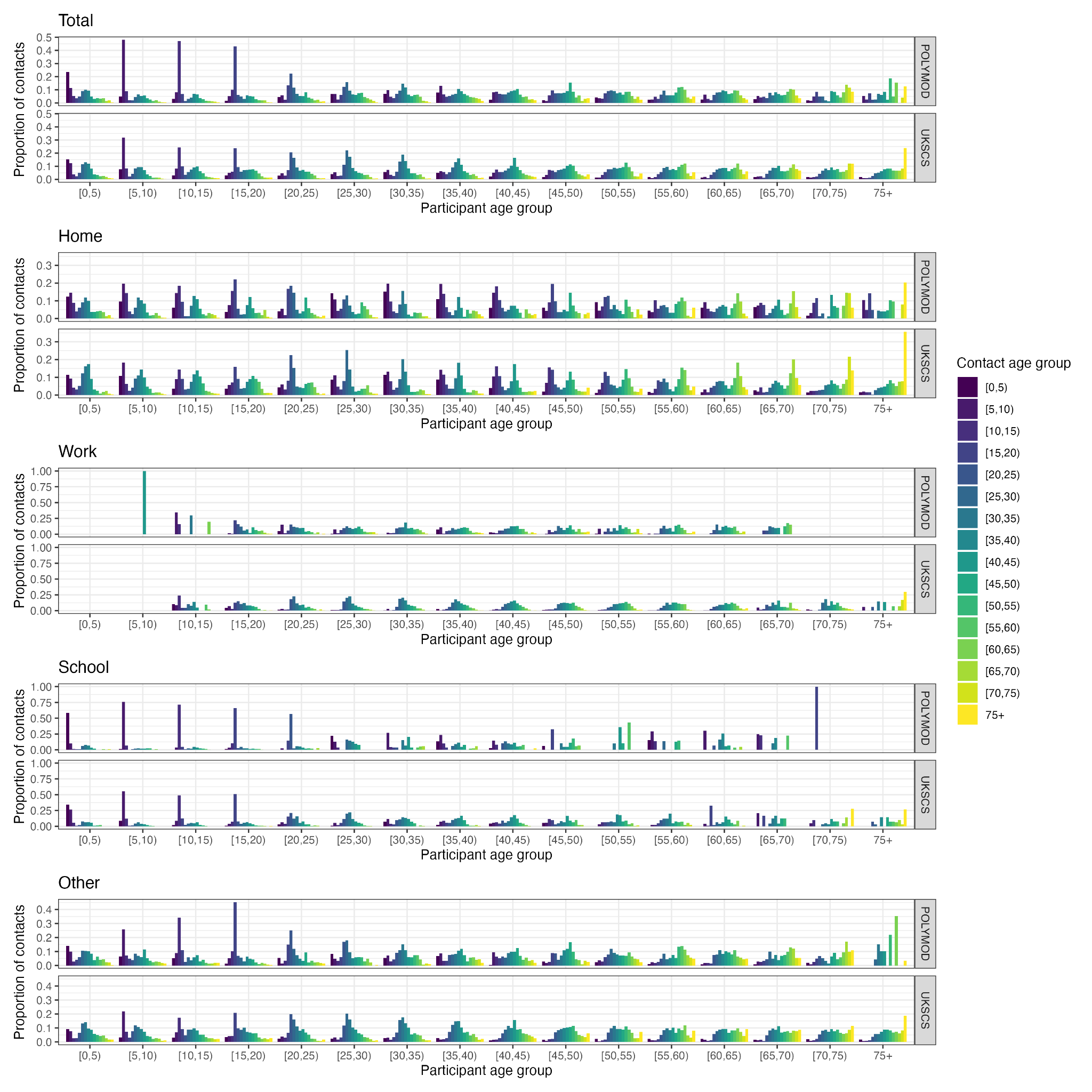

**Figure S4:** The age distribution of setting-specific contacts, stratified by participant age group, in the POLYMOD survey and this survey (weighted by participant gender, ethnicity, and weekday/weekend, and excluding large group contacts).

#### d. Degree distribution

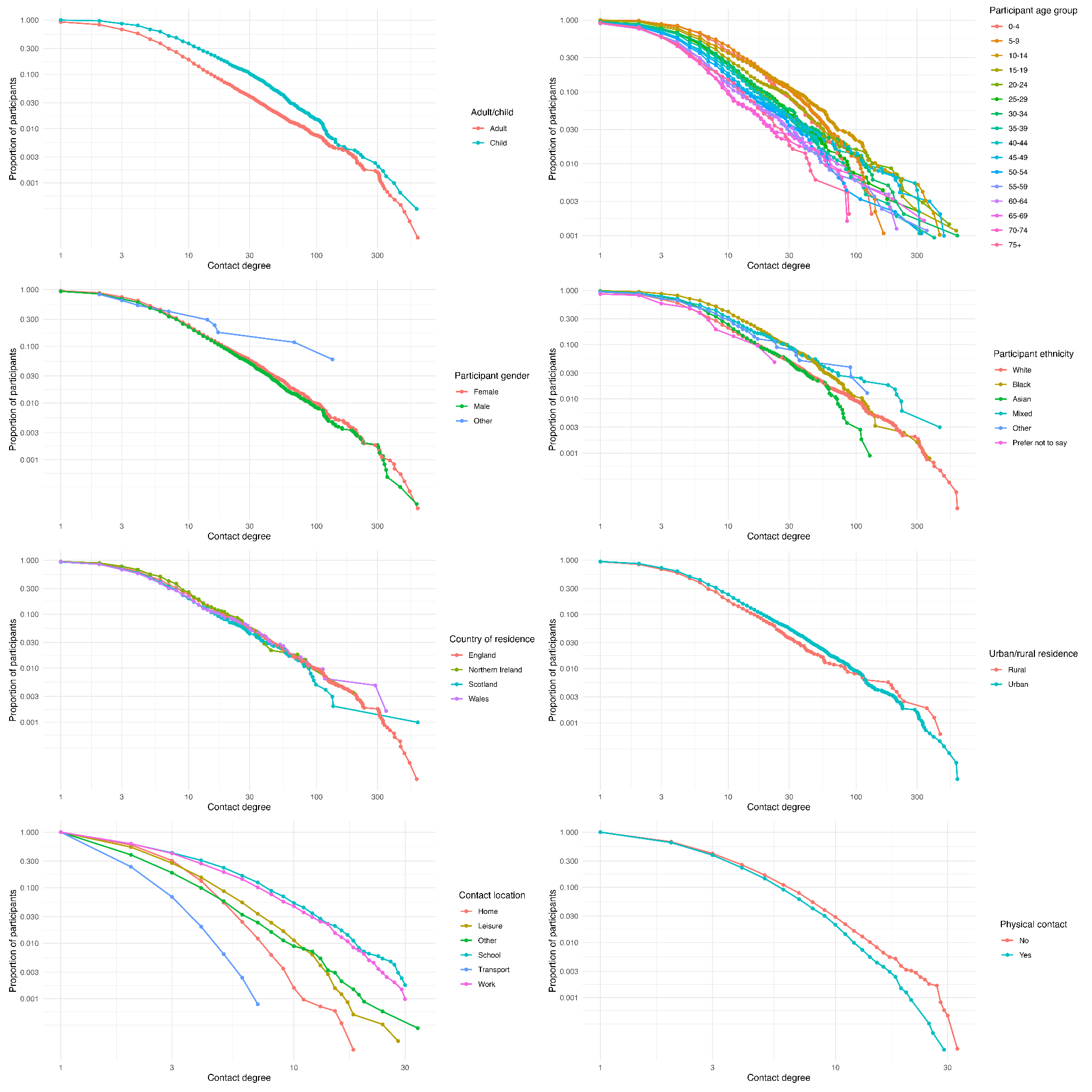

**Figure S5:** Degree distribution of participants, grouped by various participant and contact characteristics. Proportions shown on a log-log scale.

#### e. Geographical analyses

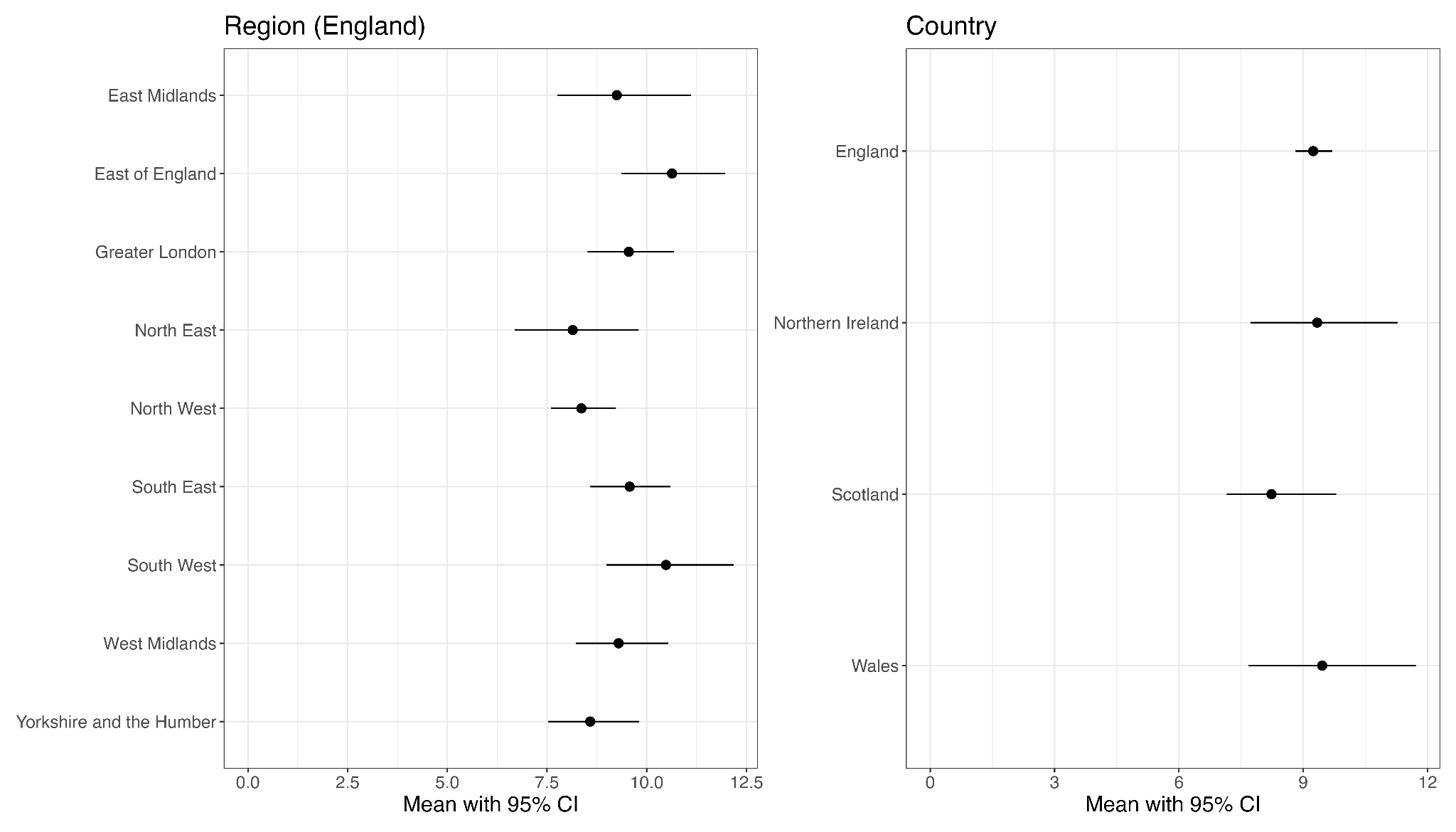

**Figure S6:** Mean daily number of social contacts (with bootstrapped 95% confidence interval) across regions of England and countries in the UK, from the weighted negative binomial regression model. Estimates are weighted by participants’ age group, gender, and weekday/weekend survey day.

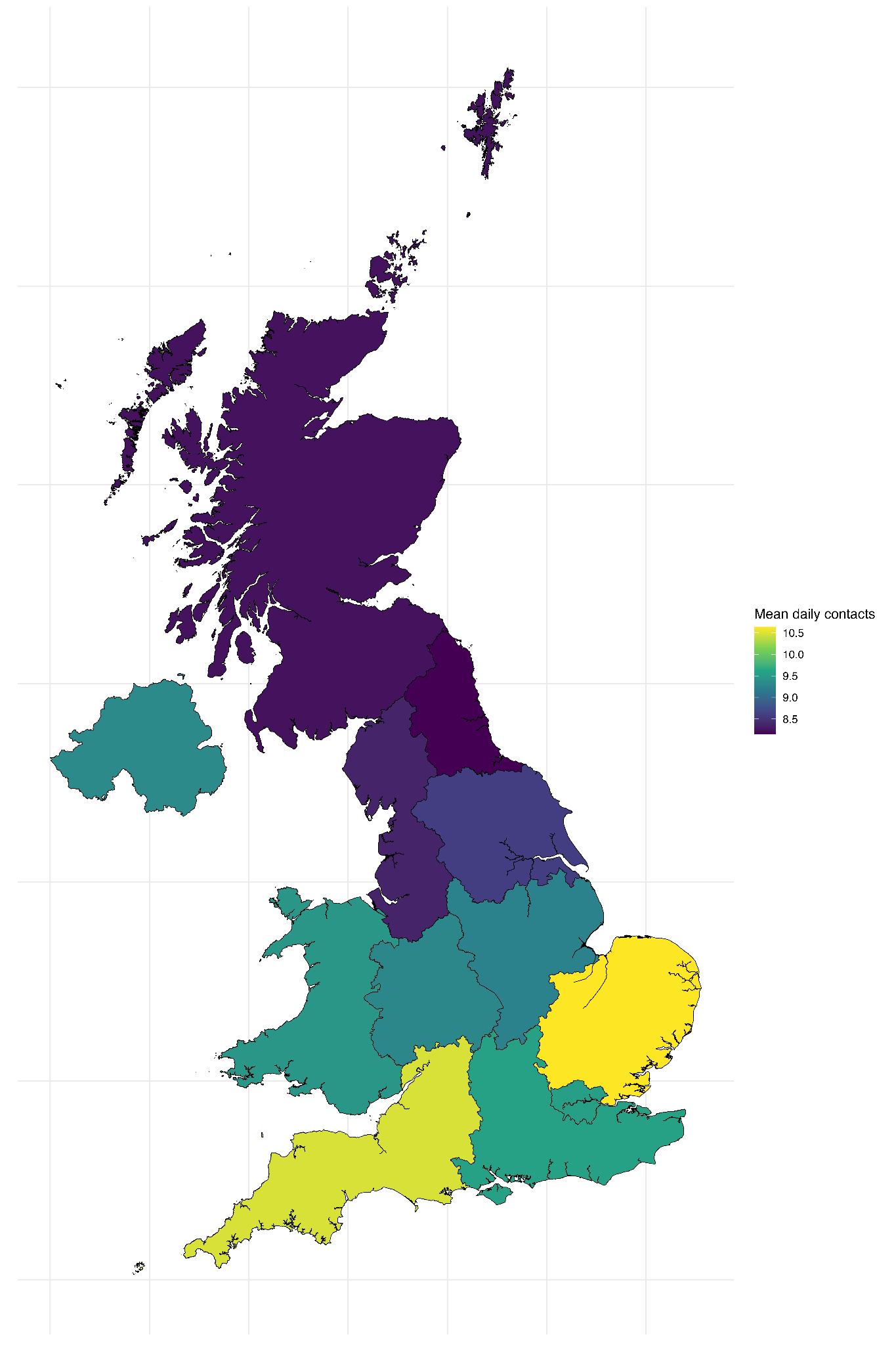

**Figure S7:** Mean daily number of social contacts across regions of England and countries in the UK, from the weighted negative binomial regression model. Estimates are weighted by participants’ age group, gender, and weekday/weekend survey day.

#### f. Mixing patterns

| **Stratifying variable** | **Participant variable** | **Contact variable** | **Setting** | | | | |
| --- | --- | --- | --- | --- | --- | --- | --- |
|  |  |  | **All settings** | **Home** | **Work** | **School** | **Other** |
| Age groups | 0-4 | 0-4 | 2.16 (1.8 - 2.57) | 0.2 (0.16 - 0.24) | 0 (0 - 0) | 0.91 (0.57 - 1.35) | 1.11 (0.92 - 1.34) |
| Age groups | 0-4 | 5-9 | 1.09 (0.97 - 1.22) | 0.2 (0.17 - 0.22) | 0 (0 - 0) | 0.28 (0.22 - 0.35) | 0.75 (0.65 - 0.84) |
| Age groups | 0-4 | 10-14 | 0.55 (0.47 - 0.63) | 0.07 (0.06 - 0.09) | 0 (0 - 0) | 0.05 (0.03 - 0.08) | 0.43 (0.35 - 0.51) |
| Age groups | 0-4 | 15-19 | 0.32 (0.27 - 0.37) | 0.06 (0.04 - 0.08) | 0.01 (0 - 0.01) | 0.02 (0.01 - 0.04) | 0.21 (0.18 - 0.25) |
| Age groups | 0-4 | 20-24 | 0.56 (0.42 - 0.72) | 0.08 (0.06 - 0.1) | 0.01 (0 - 0.04) | 0.02 (0.01 - 0.03) | 0.49 (0.34 - 0.68) |
| Age groups | 0-4 | 25-29 | 0.84 (0.74 - 0.94) | 0.17 (0.14 - 0.19) | 0.01 (0 - 0.01) | 0.09 (0.05 - 0.15) | 0.62 (0.54 - 0.71) |
| Age groups | 0-4 | 30-34 | 1.01 (0.91 - 1.13) | 0.24 (0.21 - 0.27) | 0 (0 - 0.01) | 0.07 (0.05 - 0.1) | 0.64 (0.55 - 0.76) |
| Age groups | 0-4 | 35-39 | 0.85 (0.76 - 0.93) | 0.22 (0.19 - 0.25) | 0.02 (0.01 - 0.04) | 0.07 (0.05 - 0.1) | 0.56 (0.48 - 0.64) |
| Age groups | 0-4 | 40-44 | 0.47 (0.41 - 0.54) | 0.11 (0.09 - 0.13) | 0.01 (0 - 0.02) | 0.06 (0.03 - 0.09) | 0.3 (0.25 - 0.36) |
| Age groups | 0-4 | 45-49 | 0.32 (0.27 - 0.38) | 0.04 (0.03 - 0.06) | 0.01 (0 - 0.01) | 0.03 (0.01 - 0.05) | 0.31 (0.24 - 0.4) |
| Age groups | 0-4 | 50-54 | 0.37 (0.29 - 0.49) | 0.03 (0.02 - 0.04) | 0 (0 - 0.01) | 0.01 (0 - 0.02) | 0.31 (0.24 - 0.41) |
| Age groups | 0-4 | 55-59 | 0.26 (0.21 - 0.31) | 0.02 (0.01 - 0.03) | 0.01 (0 - 0.02) | 0.01 (0 - 0.01) | 0.2 (0.16 - 0.24) |
| Age groups | 0-4 | 60-64 | 0.26 (0.2 - 0.33) | 0.02 (0.02 - 0.03) | 0 (0 - 0) | 0 (0 - 0.01) | 0.22 (0.17 - 0.29) |
| Age groups | 0-4 | 65-69 | 0.25 (0.18 - 0.33) | 0.03 (0.02 - 0.04) | 0 (0 - 0) | 0 (0 - 0.01) | 0.24 (0.15 - 0.34) |
| Age groups | 0-4 | 70-74 | 0.26 (0.18 - 0.35) | 0.01 (0.01 - 0.02) | 0 (0 - 0.01) | 0 (0 - 0.01) | 0.23 (0.15 - 0.31) |
| Age groups | 0-4 | 75+ | 0.13 (0.08 - 0.17) | 0.02 (0.01 - 0.03) | 0 (0 - 0) | 0 (0 - 0) | 0.13 (0.09 - 0.17) |
| Age groups | 5-9 | 0-4 | 1 (0.88 - 1.12) | 0.18 (0.15 - 0.2) | 0 (0 - 0) | 0.26 (0.2 - 0.32) | 0.68 (0.59 - 0.77) |
| Age groups | 5-9 | 5-9 | 5.51 (4.89 - 6.16) | 0.36 (0.32 - 0.4) | 0 (0 - 0) | 2.17 (1.71 - 2.65) | 2.63 (2.28 - 3.02) |
| Age groups | 5-9 | 10-14 | 1.28 (1.14 - 1.42) | 0.15 (0.13 - 0.17) | 0 (0 - 0) | 0.26 (0.22 - 0.31) | 0.91 (0.79 - 1.05) |
| Age groups | 5-9 | 15-19 | 0.49 (0.41 - 0.58) | 0.07 (0.06 - 0.09) | 0.01 (0 - 0.02) | 0.06 (0.03 - 0.09) | 0.38 (0.32 - 0.44) |
| Age groups | 5-9 | 20-24 | 0.59 (0.43 - 0.79) | 0.08 (0.06 - 0.1) | 0.04 (0.01 - 0.09) | 0.05 (0.03 - 0.07) | 0.25 (0.2 - 0.31) |
| Age groups | 5-9 | 25-29 | 0.72 (0.65 - 0.8) | 0.16 (0.14 - 0.18) | 0.02 (0.01 - 0.03) | 0.08 (0.05 - 0.12) | 0.46 (0.4 - 0.52) |
| Age groups | 5-9 | 30-34 | 1.04 (0.92 - 1.17) | 0.21 (0.19 - 0.24) | 0.02 (0.01 - 0.04) | 0.14 (0.09 - 0.21) | 0.59 (0.49 - 0.7) |
| Age groups | 5-9 | 35-39 | 0.89 (0.81 - 0.99) | 0.24 (0.22 - 0.27) | 0.02 (0.01 - 0.02) | 0.08 (0.06 - 0.12) | 0.5 (0.44 - 0.57) |
| Age groups | 5-9 | 40-44 | 0.8 (0.66 - 0.97) | 0.18 (0.15 - 0.2) | 0.01 (0.01 - 0.02) | 0.11 (0.05 - 0.26) | 0.68 (0.53 - 0.9) |
| Age groups | 5-9 | 45-49 | 0.37 (0.33 - 0.43) | 0.09 (0.08 - 0.11) | 0.01 (0 - 0.02) | 0.05 (0.03 - 0.06) | 0.25 (0.21 - 0.31) |
| Age groups | 5-9 | 50-54 | 0.34 (0.28 - 0.43) | 0.05 (0.04 - 0.07) | 0.02 (0 - 0.04) | 0.04 (0.02 - 0.05) | 0.26 (0.19 - 0.39) |
| Age groups | 5-9 | 55-59 | 0.17 (0.14 - 0.22) | 0.02 (0.01 - 0.03) | 0 (0 - 0) | 0.01 (0.01 - 0.02) | 0.18 (0.14 - 0.24) |
| Age groups | 5-9 | 60-64 | 0.32 (0.21 - 0.46) | 0.03 (0.02 - 0.04) | 0 (0 - 0) | 0.02 (0.01 - 0.03) | 0.21 (0.14 - 0.32) |
| Age groups | 5-9 | 65-69 | 0.26 (0.2 - 0.33) | 0.02 (0.02 - 0.03) | 0.01 (0 - 0.02) | 0.01 (0 - 0.02) | 0.2 (0.15 - 0.25) |
| Age groups | 5-9 | 70-74 | 0.15 (0.11 - 0.19) | 0.02 (0.01 - 0.02) | 0 (0 - 0.01) | 0.01 (0 - 0.01) | 0.12 (0.08 - 0.15) |
| Age groups | 5-9 | 75+ | 0.23 (0.17 - 0.3) | 0.02 (0.01 - 0.03) | 0 (0 - 0.01) | 0.01 (0 - 0.02) | 0.2 (0.15 - 0.27) |
| Age groups | 10-14 | 0-4 | 0.48 (0.41 - 0.55) | 0.07 (0.05 - 0.08) | 0 (0 - 0) | 0.05 (0.03 - 0.07) | 0.37 (0.31 - 0.44) |
| Age groups | 10-14 | 5-9 | 1.22 (1.09 - 1.35) | 0.14 (0.12 - 0.16) | 0 (0 - 0) | 0.25 (0.21 - 0.3) | 0.86 (0.75 - 1) |
| Age groups | 10-14 | 10-14 | 6.71 (5.44 - 8.1) | 0.27 (0.23 - 0.3) | 0.01 (0 - 0.01) | 2.18 (1.42 - 3.07) | 3.84 (2.97 - 4.77) |
| Age groups | 10-14 | 15-19 | 1.16 (0.99 - 1.34) | 0.14 (0.12 - 0.16) | 0 (0 - 0.01) | 0.25 (0.17 - 0.36) | 0.88 (0.76 - 1.04) |
| Age groups | 10-14 | 20-24 | 0.32 (0.25 - 0.41) | 0.05 (0.04 - 0.06) | 0.01 (0 - 0.01) | 0.02 (0.02 - 0.03) | 0.32 (0.22 - 0.43) |
| Age groups | 10-14 | 25-29 | 0.39 (0.35 - 0.44) | 0.06 (0.05 - 0.07) | 0.01 (0 - 0.01) | 0.05 (0.03 - 0.07) | 0.37 (0.32 - 0.42) |
| Age groups | 10-14 | 30-34 | 0.59 (0.52 - 0.67) | 0.13 (0.11 - 0.14) | 0.01 (0 - 0.02) | 0.06 (0.04 - 0.08) | 0.34 (0.29 - 0.4) |
| Age groups | 10-14 | 35-39 | 0.78 (0.7 - 0.87) | 0.19 (0.17 - 0.21) | 0.01 (0 - 0.01) | 0.08 (0.06 - 0.1) | 0.39 (0.34 - 0.45) |
| Age groups | 10-14 | 40-44 | 0.94 (0.79 - 1.12) | 0.24 (0.22 - 0.27) | 0.01 (0 - 0.03) | 0.1 (0.06 - 0.18) | 0.54 (0.43 - 0.68) |
| Age groups | 10-14 | 45-49 | 0.73 (0.6 - 0.88) | 0.2 (0.17 - 0.22) | 0.04 (0.01 - 0.07) | 0.04 (0.03 - 0.05) | 0.3 (0.23 - 0.38) |
| Age groups | 10-14 | 50-54 | 0.42 (0.35 - 0.51) | 0.1 (0.09 - 0.12) | 0.01 (0 - 0.01) | 0.03 (0.02 - 0.04) | 0.31 (0.24 - 0.4) |
| Age groups | 10-14 | 55-59 | 0.27 (0.22 - 0.35) | 0.04 (0.03 - 0.06) | 0.01 (0 - 0.02) | 0.01 (0 - 0.02) | 0.17 (0.14 - 0.22) |
| Age groups | 10-14 | 60-64 | 0.2 (0.15 - 0.28) | 0.03 (0.02 - 0.04) | 0 (0 - 0) | 0 (0 - 0.01) | 0.17 (0.1 - 0.26) |
| Age groups | 10-14 | 65-69 | 0.2 (0.15 - 0.25) | 0.03 (0.02 - 0.04) | 0 (0 - 0.01) | 0 (0 - 0.01) | 0.12 (0.09 - 0.15) |
| Age groups | 10-14 | 70-74 | 0.19 (0.14 - 0.24) | 0.02 (0.01 - 0.02) | 0 (0 - 0) | 0.01 (0 - 0.01) | 0.15 (0.11 - 0.2) |
| Age groups | 10-14 | 75+ | 0.18 (0.14 - 0.23) | 0.02 (0.01 - 0.03) | 0 (0 - 0) | 0.01 (0 - 0.01) | 0.22 (0.16 - 0.28) |
| Age groups | 15-19 | 0-4 | 0.29 (0.25 - 0.34) | 0.06 (0.04 - 0.07) | 0 (0 - 0.01) | 0.02 (0.01 - 0.04) | 0.2 (0.17 - 0.23) |
| Age groups | 15-19 | 5-9 | 0.49 (0.41 - 0.58) | 0.07 (0.06 - 0.09) | 0.01 (0 - 0.02) | 0.06 (0.03 - 0.09) | 0.38 (0.32 - 0.45) |
| Age groups | 15-19 | 10-14 | 1.22 (1.05 - 1.41) | 0.15 (0.13 - 0.17) | 0 (0 - 0.01) | 0.27 (0.18 - 0.38) | 0.93 (0.8 - 1.1) |
| Age groups | 15-19 | 15-19 | 4.29 (3.42 - 5.25) | 0.27 (0.24 - 0.31) | 0.04 (0.02 - 0.07) | 1.52 (0.73 - 2.6) | 2.82 (2.31 - 3.42) |
| Age groups | 15-19 | 20-24 | 1.08 (0.84 - 1.37) | 0.13 (0.11 - 0.15) | 0.04 (0.02 - 0.07) | 0.13 (0.06 - 0.23) | 0.89 (0.64 - 1.2) |
| Age groups | 15-19 | 25-29 | 0.49 (0.42 - 0.57) | 0.05 (0.04 - 0.06) | 0.04 (0.02 - 0.05) | 0.05 (0.02 - 0.09) | 0.38 (0.33 - 0.43) |
| Age groups | 15-19 | 30-34 | 0.42 (0.37 - 0.48) | 0.06 (0.05 - 0.07) | 0.02 (0.01 - 0.04) | 0.05 (0.03 - 0.07) | 0.32 (0.28 - 0.38) |
| Age groups | 15-19 | 35-39 | 0.52 (0.45 - 0.59) | 0.11 (0.09 - 0.12) | 0.03 (0.02 - 0.04) | 0.05 (0.03 - 0.08) | 0.31 (0.27 - 0.36) |
| Age groups | 15-19 | 40-44 | 0.63 (0.54 - 0.73) | 0.16 (0.14 - 0.18) | 0.03 (0.02 - 0.04) | 0.03 (0.02 - 0.05) | 0.34 (0.27 - 0.42) |
| Age groups | 15-19 | 45-49 | 0.79 (0.65 - 0.94) | 0.19 (0.17 - 0.21) | 0.03 (0.01 - 0.05) | 0.09 (0.04 - 0.19) | 0.5 (0.38 - 0.67) |
| Age groups | 15-19 | 50-54 | 0.53 (0.44 - 0.64) | 0.16 (0.14 - 0.18) | 0.02 (0.01 - 0.03) | 0.02 (0.01 - 0.04) | 0.29 (0.22 - 0.42) |
| Age groups | 15-19 | 55-59 | 0.3 (0.26 - 0.34) | 0.11 (0.1 - 0.13) | 0.01 (0.01 - 0.02) | 0.01 (0 - 0.03) | 0.14 (0.11 - 0.18) |
| Age groups | 15-19 | 60-64 | 0.21 (0.17 - 0.25) | 0.05 (0.04 - 0.07) | 0.02 (0.01 - 0.05) | 0.03 (0.01 - 0.05) | 0.13 (0.09 - 0.19) |
| Age groups | 15-19 | 65-69 | 0.32 (0.21 - 0.47) | 0.02 (0.01 - 0.02) | 0.02 (0 - 0.07) | 0.01 (0 - 0.02) | 0.23 (0.15 - 0.35) |
| Age groups | 15-19 | 70-74 | 0.18 (0.13 - 0.24) | 0.02 (0.01 - 0.03) | 0.01 (0 - 0.02) | 0.01 (0 - 0.02) | 0.16 (0.11 - 0.22) |
| Age groups | 15-19 | 75+ | 0.22 (0.16 - 0.27) | 0.02 (0.01 - 0.03) | 0.01 (0 - 0.02) | 0.01 (0 - 0.03) | 0.14 (0.1 - 0.19) |
| Age groups | 20-24 | 0-4 | 0.49 (0.37 - 0.63) | 0.07 (0.05 - 0.08) | 0.01 (0 - 0.03) | 0.02 (0.01 - 0.02) | 0.43 (0.3 - 0.6) |
| Age groups | 20-24 | 5-9 | 0.57 (0.41 - 0.76) | 0.08 (0.06 - 0.09) | 0.04 (0.01 - 0.09) | 0.05 (0.03 - 0.07) | 0.24 (0.19 - 0.3) |
| Age groups | 20-24 | 10-14 | 0.32 (0.25 - 0.41) | 0.05 (0.04 - 0.06) | 0.01 (0 - 0.02) | 0.02 (0.02 - 0.03) | 0.32 (0.23 - 0.44) |
| Age groups | 20-24 | 15-19 | 1.03 (0.81 - 1.31) | 0.13 (0.11 - 0.15) | 0.04 (0.02 - 0.06) | 0.12 (0.06 - 0.22) | 0.85 (0.61 - 1.15) |
| Age groups | 20-24 | 20-24 | 2.04 (1.66 - 2.54) | 0.28 (0.23 - 0.33) | 0.21 (0.09 - 0.41) | 0.16 (0.07 - 0.25) | 1.55 (1.23 - 1.93) |
| Age groups | 20-24 | 25-29 | 1.15 (1 - 1.32) | 0.14 (0.12 - 0.16) | 0.2 (0.13 - 0.29) | 0.03 (0.02 - 0.04) | 0.86 (0.73 - 1.02) |
| Age groups | 20-24 | 30-34 | 0.71 (0.61 - 0.82) | 0.06 (0.05 - 0.08) | 0.11 (0.06 - 0.19) | 0.04 (0.03 - 0.06) | 0.55 (0.47 - 0.64) |
| Age groups | 20-24 | 35-39 | 0.52 (0.45 - 0.61) | 0.05 (0.04 - 0.06) | 0.1 (0.05 - 0.16) | 0.02 (0.01 - 0.03) | 0.38 (0.32 - 0.44) |
| Age groups | 20-24 | 40-44 | 0.55 (0.47 - 0.63) | 0.06 (0.05 - 0.07) | 0.1 (0.05 - 0.17) | 0.03 (0.01 - 0.04) | 0.32 (0.26 - 0.39) |
| Age groups | 20-24 | 45-49 | 0.59 (0.5 - 0.7) | 0.08 (0.06 - 0.09) | 0.06 (0.03 - 0.09) | 0.01 (0 - 0.01) | 0.44 (0.37 - 0.53) |
| Age groups | 20-24 | 50-54 | 0.53 (0.46 - 0.6) | 0.11 (0.09 - 0.13) | 0.06 (0.03 - 0.1) | 0.01 (0.01 - 0.02) | 0.27 (0.23 - 0.31) |
| Age groups | 20-24 | 55-59 | 0.39 (0.33 - 0.45) | 0.09 (0.07 - 0.1) | 0.05 (0.03 - 0.08) | 0.01 (0 - 0.01) | 0.22 (0.18 - 0.27) |
| Age groups | 20-24 | 60-64 | 0.27 (0.23 - 0.32) | 0.06 (0.04 - 0.07) | 0.04 (0.02 - 0.05) | 0 (0 - 0.01) | 0.19 (0.15 - 0.22) |
| Age groups | 20-24 | 65-69 | 0.34 (0.24 - 0.45) | 0.02 (0.01 - 0.02) | 0.09 (0.02 - 0.21) | 0.03 (0 - 0.07) | 0.23 (0.17 - 0.31) |
| Age groups | 20-24 | 70-74 | 0.19 (0.15 - 0.25) | 0.01 (0.01 - 0.02) | 0.01 (0 - 0.02) | 0 (0 - 0) | 0.19 (0.14 - 0.24) |
| Age groups | 20-24 | 75+ | 0.34 (0.24 - 0.45) | 0.01 (0 - 0.01) | 0.03 (0.01 - 0.07) | 0 (0 - 0.01) | 0.23 (0.17 - 0.31) |
| Age groups | 25-29 | 0-4 | 0.69 (0.61 - 0.77) | 0.14 (0.12 - 0.16) | 0 (0 - 0.01) | 0.07 (0.04 - 0.12) | 0.51 (0.44 - 0.58) |
| Age groups | 25-29 | 5-9 | 0.65 (0.59 - 0.72) | 0.14 (0.12 - 0.16) | 0.01 (0.01 - 0.03) | 0.07 (0.05 - 0.1) | 0.41 (0.36 - 0.46) |
| Age groups | 25-29 | 10-14 | 0.37 (0.33 - 0.41) | 0.06 (0.05 - 0.07) | 0.01 (0 - 0.01) | 0.05 (0.03 - 0.06) | 0.35 (0.3 - 0.4) |
| Age groups | 25-29 | 15-19 | 0.44 (0.38 - 0.51) | 0.05 (0.04 - 0.06) | 0.03 (0.02 - 0.05) | 0.04 (0.02 - 0.08) | 0.34 (0.3 - 0.38) |
| Age groups | 25-29 | 20-24 | 1.07 (0.93 - 1.24) | 0.13 (0.12 - 0.15) | 0.18 (0.12 - 0.27) | 0.02 (0.01 - 0.03) | 0.8 (0.68 - 0.95) |
| Age groups | 25-29 | 25-29 | 1.5 (1.28 - 1.77) | 0.29 (0.26 - 0.33) | 0.23 (0.16 - 0.33) | 0.04 (0.02 - 0.06) | 1.01 (0.81 - 1.27) |
| Age groups | 25-29 | 30-34 | 1.04 (0.95 - 1.15) | 0.15 (0.13 - 0.16) | 0.2 (0.17 - 0.25) | 0.03 (0.02 - 0.05) | 0.68 (0.6 - 0.79) |
| Age groups | 25-29 | 35-39 | 0.62 (0.56 - 0.71) | 0.05 (0.04 - 0.06) | 0.13 (0.1 - 0.18) | 0.03 (0.02 - 0.04) | 0.4 (0.35 - 0.46) |
| Age groups | 25-29 | 40-44 | 0.52 (0.45 - 0.6) | 0.03 (0.02 - 0.04) | 0.11 (0.08 - 0.15) | 0.02 (0.01 - 0.03) | 0.35 (0.29 - 0.41) |
| Age groups | 25-29 | 45-49 | 0.52 (0.44 - 0.6) | 0.04 (0.03 - 0.05) | 0.13 (0.08 - 0.19) | 0.01 (0 - 0.02) | 0.27 (0.23 - 0.31) |
| Age groups | 25-29 | 50-54 | 0.56 (0.48 - 0.64) | 0.05 (0.04 - 0.07) | 0.07 (0.04 - 0.1) | 0.01 (0 - 0.01) | 0.41 (0.36 - 0.48) |
| Age groups | 25-29 | 55-59 | 0.46 (0.39 - 0.53) | 0.07 (0.06 - 0.09) | 0.06 (0.04 - 0.09) | 0 (0 - 0.01) | 0.27 (0.23 - 0.34) |
| Age groups | 25-29 | 60-64 | 0.35 (0.31 - 0.4) | 0.07 (0.06 - 0.08) | 0.05 (0.03 - 0.08) | 0 (0 - 0.01) | 0.25 (0.21 - 0.29) |
| Age groups | 25-29 | 65-69 | 0.38 (0.28 - 0.49) | 0.02 (0.02 - 0.03) | 0.07 (0.02 - 0.13) | 0 (0 - 0) | 0.36 (0.28 - 0.47) |
| Age groups | 25-29 | 70-74 | 0.22 (0.18 - 0.28) | 0.01 (0.01 - 0.02) | 0.05 (0.01 - 0.12) | 0 (0 - 0) | 0.1 (0.08 - 0.12) |
| Age groups | 25-29 | 75+ | 0.22 (0.18 - 0.26) | 0.02 (0.01 - 0.03) | 0.01 (0 - 0.02) | 0 (0 - 0.01) | 0.19 (0.15 - 0.23) |
| Age groups | 30-34 | 0-4 | 0.78 (0.69 - 0.86) | 0.18 (0.16 - 0.21) | 0 (0 - 0) | 0.06 (0.04 - 0.08) | 0.49 (0.42 - 0.58) |
| Age groups | 30-34 | 5-9 | 0.87 (0.77 - 0.99) | 0.18 (0.16 - 0.2) | 0.02 (0.01 - 0.03) | 0.12 (0.08 - 0.18) | 0.49 (0.41 - 0.58) |
| Age groups | 30-34 | 10-14 | 0.52 (0.46 - 0.59) | 0.11 (0.1 - 0.13) | 0.01 (0 - 0.02) | 0.05 (0.04 - 0.07) | 0.3 (0.26 - 0.36) |
| Age groups | 30-34 | 15-19 | 0.35 (0.31 - 0.4) | 0.05 (0.04 - 0.06) | 0.02 (0.01 - 0.03) | 0.04 (0.02 - 0.06) | 0.27 (0.23 - 0.31) |
| Age groups | 30-34 | 20-24 | 0.62 (0.53 - 0.72) | 0.05 (0.04 - 0.07) | 0.1 (0.06 - 0.17) | 0.04 (0.02 - 0.05) | 0.47 (0.41 - 0.55) |
| Age groups | 30-34 | 25-29 | 0.97 (0.88 - 1.08) | 0.14 (0.12 - 0.15) | 0.19 (0.15 - 0.23) | 0.03 (0.02 - 0.04) | 0.64 (0.56 - 0.73) |
| Age groups | 30-34 | 30-34 | 1.47 (1.28 - 1.67) | 0.27 (0.24 - 0.3) | 0.17 (0.14 - 0.21) | 0.05 (0.03 - 0.08) | 0.94 (0.79 - 1.14) |
| Age groups | 30-34 | 35-39 | 0.95 (0.86 - 1.05) | 0.14 (0.13 - 0.16) | 0.15 (0.12 - 0.17) | 0.04 (0.03 - 0.05) | 0.58 (0.51 - 0.67) |
| Age groups | 30-34 | 40-44 | 0.64 (0.57 - 0.72) | 0.06 (0.05 - 0.07) | 0.09 (0.08 - 0.11) | 0.04 (0.02 - 0.05) | 0.46 (0.39 - 0.55) |
| Age groups | 30-34 | 45-49 | 0.46 (0.4 - 0.53) | 0.03 (0.02 - 0.04) | 0.11 (0.07 - 0.14) | 0.04 (0.01 - 0.08) | 0.3 (0.26 - 0.35) |
| Age groups | 30-34 | 50-54 | 0.54 (0.48 - 0.6) | 0.04 (0.03 - 0.05) | 0.07 (0.05 - 0.08) | 0.01 (0.01 - 0.02) | 0.43 (0.37 - 0.49) |
| Age groups | 30-34 | 55-59 | 0.47 (0.41 - 0.53) | 0.05 (0.04 - 0.06) | 0.07 (0.05 - 0.09) | 0.01 (0 - 0.01) | 0.33 (0.28 - 0.39) |
| Age groups | 30-34 | 60-64 | 0.39 (0.34 - 0.44) | 0.06 (0.05 - 0.07) | 0.04 (0.03 - 0.06) | 0.01 (0 - 0.01) | 0.29 (0.24 - 0.34) |
| Age groups | 30-34 | 65-69 | 0.48 (0.35 - 0.65) | 0.04 (0.03 - 0.05) | 0.03 (0.02 - 0.06) | 0.01 (0 - 0.01) | 0.21 (0.16 - 0.29) |
| Age groups | 30-34 | 70-74 | 0.31 (0.22 - 0.43) | 0.02 (0.01 - 0.03) | 0.02 (0.01 - 0.03) | 0 (0 - 0) | 0.29 (0.2 - 0.42) |
| Age groups | 30-34 | 75+ | 0.36 (0.28 - 0.45) | 0.03 (0.02 - 0.04) | 0.01 (0 - 0.01) | 0 (0 - 0.01) | 0.52 (0.39 - 0.73) |
| Age groups | 35-39 | 0-4 | 0.67 (0.6 - 0.74) | 0.17 (0.15 - 0.2) | 0.02 (0 - 0.03) | 0.06 (0.04 - 0.08) | 0.44 (0.38 - 0.51) |
| Age groups | 35-39 | 5-9 | 0.77 (0.7 - 0.85) | 0.21 (0.19 - 0.23) | 0.01 (0.01 - 0.02) | 0.07 (0.05 - 0.1) | 0.43 (0.38 - 0.49) |
| Age groups | 35-39 | 10-14 | 0.71 (0.64 - 0.79) | 0.17 (0.15 - 0.19) | 0.01 (0 - 0.01) | 0.07 (0.05 - 0.09) | 0.35 (0.31 - 0.41) |
| Age groups | 35-39 | 15-19 | 0.45 (0.39 - 0.51) | 0.09 (0.08 - 0.11) | 0.02 (0.02 - 0.03) | 0.04 (0.03 - 0.07) | 0.27 (0.24 - 0.31) |
| Age groups | 35-39 | 20-24 | 0.47 (0.4 - 0.55) | 0.05 (0.04 - 0.06) | 0.09 (0.05 - 0.15) | 0.02 (0.01 - 0.02) | 0.34 (0.29 - 0.39) |
| Age groups | 35-39 | 25-29 | 0.6 (0.54 - 0.68) | 0.05 (0.04 - 0.06) | 0.13 (0.09 - 0.17) | 0.03 (0.02 - 0.04) | 0.39 (0.33 - 0.45) |
| Age groups | 35-39 | 30-34 | 0.98 (0.89 - 1.09) | 0.15 (0.13 - 0.17) | 0.15 (0.12 - 0.18) | 0.04 (0.03 - 0.05) | 0.6 (0.53 - 0.7) |
| Age groups | 35-39 | 35-39 | 1.05 (0.92 - 1.21) | 0.22 (0.2 - 0.25) | 0.13 (0.1 - 0.17) | 0.04 (0.03 - 0.06) | 0.58 (0.48 - 0.71) |
| Age groups | 35-39 | 40-44 | 0.78 (0.69 - 0.88) | 0.12 (0.1 - 0.13) | 0.12 (0.1 - 0.15) | 0.04 (0.03 - 0.05) | 0.54 (0.46 - 0.64) |
| Age groups | 35-39 | 45-49 | 0.53 (0.46 - 0.6) | 0.05 (0.04 - 0.05) | 0.11 (0.08 - 0.15) | 0.03 (0.02 - 0.05) | 0.37 (0.32 - 0.42) |
| Age groups | 35-39 | 50-54 | 0.48 (0.43 - 0.54) | 0.03 (0.02 - 0.04) | 0.09 (0.07 - 0.11) | 0.02 (0.01 - 0.03) | 0.29 (0.25 - 0.33) |
| Age groups | 35-39 | 55-59 | 0.37 (0.32 - 0.43) | 0.03 (0.02 - 0.04) | 0.07 (0.05 - 0.09) | 0.01 (0 - 0.01) | 0.27 (0.23 - 0.32) |
| Age groups | 35-39 | 60-64 | 0.4 (0.35 - 0.45) | 0.06 (0.04 - 0.07) | 0.05 (0.03 - 0.07) | 0.01 (0 - 0.01) | 0.28 (0.24 - 0.33) |
| Age groups | 35-39 | 65-69 | 0.37 (0.3 - 0.46) | 0.05 (0.04 - 0.06) | 0.04 (0.02 - 0.07) | 0.01 (0 - 0.02) | 0.24 (0.18 - 0.31) |
| Age groups | 35-39 | 70-74 | 0.22 (0.17 - 0.28) | 0.03 (0.02 - 0.03) | 0.03 (0.01 - 0.06) | 0 (0 - 0) | 0.18 (0.14 - 0.23) |
| Age groups | 35-39 | 75+ | 0.34 (0.28 - 0.41) | 0.03 (0.02 - 0.05) | 0.02 (0 - 0.05) | 0 (0 - 0.01) | 0.3 (0.24 - 0.35) |
| Age groups | 40-44 | 0-4 | 0.39 (0.34 - 0.45) | 0.09 (0.07 - 0.11) | 0.01 (0 - 0.01) | 0.05 (0.03 - 0.08) | 0.25 (0.21 - 0.3) |
| Age groups | 40-44 | 5-9 | 0.73 (0.6 - 0.88) | 0.16 (0.14 - 0.18) | 0.01 (0 - 0.02) | 0.1 (0.04 - 0.24) | 0.62 (0.48 - 0.82) |
| Age groups | 40-44 | 10-14 | 0.89 (0.75 - 1.06) | 0.23 (0.21 - 0.26) | 0.01 (0 - 0.03) | 0.1 (0.06 - 0.17) | 0.51 (0.41 - 0.64) |
| Age groups | 40-44 | 15-19 | 0.56 (0.48 - 0.66) | 0.14 (0.12 - 0.16) | 0.02 (0.01 - 0.04) | 0.03 (0.02 - 0.04) | 0.3 (0.25 - 0.37) |
| Age groups | 40-44 | 20-24 | 0.51 (0.44 - 0.59) | 0.05 (0.04 - 0.07) | 0.09 (0.05 - 0.16) | 0.02 (0.01 - 0.04) | 0.3 (0.24 - 0.36) |
| Age groups | 40-44 | 25-29 | 0.53 (0.46 - 0.61) | 0.03 (0.02 - 0.04) | 0.11 (0.08 - 0.15) | 0.02 (0.01 - 0.03) | 0.35 (0.3 - 0.41) |
| Age groups | 40-44 | 30-34 | 0.69 (0.61 - 0.78) | 0.07 (0.05 - 0.08) | 0.1 (0.08 - 0.12) | 0.04 (0.02 - 0.06) | 0.49 (0.42 - 0.59) |
| Age groups | 40-44 | 35-39 | 0.81 (0.72 - 0.92) | 0.12 (0.11 - 0.14) | 0.13 (0.1 - 0.16) | 0.04 (0.03 - 0.05) | 0.57 (0.48 - 0.67) |
| Age groups | 40-44 | 40-44 | 1.03 (0.88 - 1.2) | 0.24 (0.2 - 0.29) | 0.14 (0.11 - 0.18) | 0.04 (0.03 - 0.06) | 0.61 (0.48 - 0.79) |
| Age groups | 40-44 | 45-49 | 0.75 (0.66 - 0.86) | 0.11 (0.09 - 0.12) | 0.13 (0.1 - 0.17) | 0.04 (0.02 - 0.06) | 0.53 (0.44 - 0.64) |
| Age groups | 40-44 | 50-54 | 0.56 (0.5 - 0.63) | 0.06 (0.05 - 0.07) | 0.09 (0.07 - 0.11) | 0.02 (0.01 - 0.03) | 0.39 (0.33 - 0.46) |
| Age groups | 40-44 | 55-59 | 0.41 (0.36 - 0.48) | 0.02 (0.02 - 0.03) | 0.08 (0.06 - 0.1) | 0.01 (0 - 0.01) | 0.32 (0.27 - 0.39) |
| Age groups | 40-44 | 60-64 | 0.41 (0.35 - 0.47) | 0.04 (0.03 - 0.05) | 0.06 (0.04 - 0.09) | 0.01 (0 - 0.02) | 0.32 (0.27 - 0.38) |
| Age groups | 40-44 | 65-69 | 0.37 (0.3 - 0.46) | 0.04 (0.03 - 0.05) | 0.03 (0.02 - 0.05) | 0 (0 - 0) | 0.27 (0.2 - 0.36) |
| Age groups | 40-44 | 70-74 | 0.34 (0.28 - 0.42) | 0.04 (0.03 - 0.05) | 0.02 (0.01 - 0.04) | 0 (0 - 0) | 0.27 (0.21 - 0.35) |
| Age groups | 40-44 | 75+ | 0.49 (0.39 - 0.59) | 0.05 (0.03 - 0.07) | 0.01 (0.01 - 0.02) | 0 (0 - 0.01) | 0.38 (0.3 - 0.47) |
| Age groups | 45-49 | 0-4 | 0.28 (0.23 - 0.33) | 0.04 (0.03 - 0.05) | 0 (0 - 0.01) | 0.02 (0.01 - 0.04) | 0.27 (0.21 - 0.35) |
| Age groups | 45-49 | 5-9 | 0.36 (0.32 - 0.41) | 0.09 (0.07 - 0.1) | 0.01 (0 - 0.02) | 0.04 (0.03 - 0.06) | 0.24 (0.2 - 0.3) |
| Age groups | 45-49 | 10-14 | 0.74 (0.61 - 0.89) | 0.2 (0.17 - 0.22) | 0.04 (0.01 - 0.07) | 0.04 (0.03 - 0.05) | 0.3 (0.23 - 0.38) |
| Age groups | 45-49 | 15-19 | 0.75 (0.62 - 0.89) | 0.18 (0.16 - 0.2) | 0.03 (0.01 - 0.05) | 0.09 (0.03 - 0.18) | 0.48 (0.36 - 0.63) |
| Age groups | 45-49 | 20-24 | 0.59 (0.49 - 0.7) | 0.08 (0.06 - 0.09) | 0.06 (0.03 - 0.09) | 0.01 (0 - 0.01) | 0.44 (0.37 - 0.52) |
| Age groups | 45-49 | 25-29 | 0.55 (0.47 - 0.64) | 0.04 (0.03 - 0.05) | 0.14 (0.09 - 0.2) | 0.01 (0 - 0.02) | 0.28 (0.24 - 0.33) |
| Age groups | 45-49 | 30-34 | 0.53 (0.46 - 0.6) | 0.03 (0.03 - 0.04) | 0.12 (0.08 - 0.16) | 0.04 (0.02 - 0.09) | 0.34 (0.3 - 0.4) |
| Age groups | 45-49 | 35-39 | 0.58 (0.51 - 0.66) | 0.05 (0.04 - 0.06) | 0.13 (0.09 - 0.16) | 0.03 (0.02 - 0.06) | 0.41 (0.35 - 0.47) |
| Age groups | 45-49 | 40-44 | 0.79 (0.7 - 0.9) | 0.11 (0.1 - 0.13) | 0.14 (0.1 - 0.18) | 0.04 (0.02 - 0.06) | 0.56 (0.47 - 0.68) |
| Age groups | 45-49 | 45-49 | 1 (0.81 - 1.24) | 0.17 (0.15 - 0.19) | 0.17 (0.09 - 0.27) | 0.06 (0.02 - 0.15) | 0.58 (0.47 - 0.7) |
| Age groups | 45-49 | 50-54 | 0.64 (0.57 - 0.74) | 0.11 (0.09 - 0.12) | 0.12 (0.09 - 0.16) | 0.05 (0.01 - 0.14) | 0.4 (0.35 - 0.45) |
| Age groups | 45-49 | 55-59 | 0.45 (0.39 - 0.52) | 0.05 (0.04 - 0.06) | 0.1 (0.07 - 0.14) | 0.02 (0.01 - 0.05) | 0.25 (0.21 - 0.3) |
| Age groups | 45-49 | 60-64 | 0.36 (0.31 - 0.42) | 0.03 (0.03 - 0.04) | 0.08 (0.06 - 0.12) | 0.02 (0.01 - 0.05) | 0.22 (0.19 - 0.26) |
| Age groups | 45-49 | 65-69 | 0.25 (0.21 - 0.32) | 0.04 (0.03 - 0.04) | 0.05 (0.02 - 0.11) | 0.01 (0 - 0.02) | 0.16 (0.12 - 0.21) |
| Age groups | 45-49 | 70-74 | 0.3 (0.24 - 0.38) | 0.04 (0.03 - 0.05) | 0.04 (0.01 - 0.09) | 0.01 (0 - 0.01) | 0.24 (0.19 - 0.29) |
| Age groups | 45-49 | 75+ | 0.54 (0.43 - 0.69) | 0.07 (0.05 - 0.09) | 0.07 (0.01 - 0.19) | 0.01 (0 - 0.03) | 0.34 (0.28 - 0.42) |
| Age groups | 50-54 | 0-4 | 0.29 (0.23 - 0.38) | 0.02 (0.01 - 0.03) | 0 (0 - 0) | 0.01 (0 - 0.02) | 0.24 (0.19 - 0.32) |
| Age groups | 50-54 | 5-9 | 0.29 (0.24 - 0.37) | 0.05 (0.04 - 0.06) | 0.01 (0 - 0.03) | 0.03 (0.02 - 0.04) | 0.22 (0.16 - 0.33) |
| Age groups | 50-54 | 10-14 | 0.37 (0.31 - 0.46) | 0.09 (0.08 - 0.11) | 0.01 (0 - 0.01) | 0.03 (0.02 - 0.04) | 0.27 (0.22 - 0.36) |
| Age groups | 50-54 | 15-19 | 0.45 (0.38 - 0.54) | 0.14 (0.12 - 0.15) | 0.02 (0.01 - 0.03) | 0.02 (0.01 - 0.03) | 0.25 (0.19 - 0.35) |
| Age groups | 50-54 | 20-24 | 0.47 (0.41 - 0.53) | 0.1 (0.08 - 0.11) | 0.05 (0.03 - 0.09) | 0.01 (0 - 0.02) | 0.24 (0.2 - 0.27) |
| Age groups | 50-54 | 25-29 | 0.53 (0.46 - 0.6) | 0.05 (0.04 - 0.06) | 0.06 (0.04 - 0.1) | 0.01 (0 - 0.01) | 0.39 (0.34 - 0.46) |
| Age groups | 50-54 | 30-34 | 0.55 (0.49 - 0.61) | 0.04 (0.03 - 0.05) | 0.07 (0.05 - 0.09) | 0.01 (0.01 - 0.02) | 0.43 (0.38 - 0.5) |
| Age groups | 50-54 | 35-39 | 0.47 (0.42 - 0.53) | 0.03 (0.02 - 0.04) | 0.09 (0.06 - 0.11) | 0.02 (0.01 - 0.03) | 0.29 (0.25 - 0.33) |
| Age groups | 50-54 | 40-44 | 0.53 (0.47 - 0.6) | 0.05 (0.04 - 0.06) | 0.09 (0.07 - 0.1) | 0.02 (0.01 - 0.03) | 0.37 (0.31 - 0.44) |
| Age groups | 50-54 | 45-49 | 0.57 (0.51 - 0.66) | 0.1 (0.08 - 0.11) | 0.11 (0.08 - 0.14) | 0.05 (0.01 - 0.12) | 0.35 (0.32 - 0.4) |
| Age groups | 50-54 | 50-54 | 0.69 (0.61 - 0.77) | 0.16 (0.14 - 0.19) | 0.11 (0.08 - 0.14) | 0.01 (0.01 - 0.02) | 0.37 (0.3 - 0.43) |
| Age groups | 50-54 | 55-59 | 0.62 (0.55 - 0.72) | 0.1 (0.08 - 0.12) | 0.09 (0.07 - 0.12) | 0.01 (0 - 0.01) | 0.46 (0.38 - 0.55) |
| Age groups | 50-54 | 60-64 | 0.38 (0.33 - 0.43) | 0.05 (0.04 - 0.06) | 0.07 (0.05 - 0.09) | 0.01 (0 - 0.01) | 0.27 (0.23 - 0.31) |
| Age groups | 50-54 | 65-69 | 0.23 (0.19 - 0.28) | 0.02 (0.02 - 0.03) | 0.03 (0.02 - 0.04) | 0 (0 - 0) | 0.21 (0.16 - 0.27) |
| Age groups | 50-54 | 70-74 | 0.25 (0.22 - 0.29) | 0.03 (0.02 - 0.04) | 0.02 (0.01 - 0.03) | 0 (0 - 0) | 0.18 (0.15 - 0.21) |
| Age groups | 50-54 | 75+ | 0.55 (0.47 - 0.64) | 0.08 (0.06 - 0.09) | 0.01 (0 - 0.02) | 0 (0 - 0.01) | 0.42 (0.35 - 0.5) |
| Age groups | 55-59 | 0-4 | 0.2 (0.17 - 0.24) | 0.02 (0.01 - 0.02) | 0.01 (0 - 0.02) | 0.01 (0 - 0.01) | 0.15 (0.12 - 0.19) |
| Age groups | 55-59 | 5-9 | 0.15 (0.12 - 0.18) | 0.02 (0.01 - 0.02) | 0 (0 - 0) | 0.01 (0.01 - 0.02) | 0.15 (0.11 - 0.21) |
| Age groups | 55-59 | 10-14 | 0.24 (0.2 - 0.31) | 0.04 (0.03 - 0.05) | 0.01 (0 - 0.02) | 0.01 (0 - 0.02) | 0.15 (0.12 - 0.2) |
| Age groups | 55-59 | 15-19 | 0.25 (0.22 - 0.29) | 0.1 (0.08 - 0.11) | 0.01 (0.01 - 0.02) | 0.01 (0 - 0.02) | 0.12 (0.09 - 0.16) |
| Age groups | 55-59 | 20-24 | 0.34 (0.29 - 0.4) | 0.08 (0.06 - 0.09) | 0.04 (0.02 - 0.07) | 0 (0 - 0.01) | 0.2 (0.16 - 0.24) |
| Age groups | 55-59 | 25-29 | 0.43 (0.37 - 0.5) | 0.07 (0.06 - 0.08) | 0.06 (0.04 - 0.09) | 0 (0 - 0.01) | 0.26 (0.21 - 0.32) |
| Age groups | 55-59 | 30-34 | 0.47 (0.41 - 0.54) | 0.05 (0.04 - 0.06) | 0.07 (0.05 - 0.09) | 0.01 (0 - 0.01) | 0.34 (0.29 - 0.4) |
| Age groups | 55-59 | 35-39 | 0.36 (0.32 - 0.42) | 0.03 (0.02 - 0.03) | 0.07 (0.05 - 0.09) | 0.01 (0 - 0.01) | 0.27 (0.23 - 0.32) |
| Age groups | 55-59 | 40-44 | 0.39 (0.34 - 0.45) | 0.02 (0.02 - 0.03) | 0.07 (0.06 - 0.09) | 0.01 (0 - 0.01) | 0.3 (0.25 - 0.36) |
| Age groups | 55-59 | 45-49 | 0.4 (0.35 - 0.47) | 0.04 (0.03 - 0.05) | 0.09 (0.07 - 0.12) | 0.02 (0.01 - 0.05) | 0.22 (0.19 - 0.26) |
| Age groups | 55-59 | 50-54 | 0.62 (0.55 - 0.72) | 0.1 (0.08 - 0.11) | 0.09 (0.07 - 0.12) | 0.01 (0 - 0.01) | 0.46 (0.38 - 0.55) |
| Age groups | 55-59 | 55-59 | 0.72 (0.59 - 0.9) | 0.16 (0.14 - 0.19) | 0.08 (0.05 - 0.13) | 0 (0 - 0.01) | 0.51 (0.39 - 0.69) |
| Age groups | 55-59 | 60-64 | 0.57 (0.5 - 0.65) | 0.12 (0.1 - 0.14) | 0.07 (0.05 - 0.1) | 0 (0 - 0) | 0.39 (0.33 - 0.47) |
| Age groups | 55-59 | 65-69 | 0.31 (0.26 - 0.39) | 0.05 (0.04 - 0.06) | 0.03 (0.01 - 0.05) | 0 (0 - 0) | 0.26 (0.2 - 0.34) |
| Age groups | 55-59 | 70-74 | 0.19 (0.16 - 0.23) | 0.03 (0.02 - 0.03) | 0.02 (0.01 - 0.04) | 0 (0 - 0) | 0.17 (0.14 - 0.21) |
| Age groups | 55-59 | 75+ | 0.47 (0.4 - 0.56) | 0.07 (0.05 - 0.08) | 0.04 (0.01 - 0.1) | 0 (0 - 0.01) | 0.27 (0.22 - 0.33) |
| Age groups | 60-64 | 0-4 | 0.23 (0.17 - 0.29) | 0.02 (0.01 - 0.03) | 0 (0 - 0) | 0 (0 - 0.01) | 0.19 (0.15 - 0.25) |
| Age groups | 60-64 | 5-9 | 0.31 (0.21 - 0.44) | 0.03 (0.02 - 0.04) | 0 (0 - 0) | 0.02 (0.01 - 0.03) | 0.2 (0.14 - 0.31) |
| Age groups | 60-64 | 10-14 | 0.21 (0.15 - 0.29) | 0.03 (0.02 - 0.04) | 0 (0 - 0) | 0 (0 - 0.01) | 0.17 (0.11 - 0.27) |
| Age groups | 60-64 | 15-19 | 0.2 (0.16 - 0.24) | 0.05 (0.04 - 0.06) | 0.02 (0.01 - 0.04) | 0.03 (0.01 - 0.05) | 0.13 (0.09 - 0.18) |
| Age groups | 60-64 | 20-24 | 0.27 (0.23 - 0.32) | 0.06 (0.04 - 0.07) | 0.04 (0.02 - 0.05) | 0 (0 - 0.01) | 0.19 (0.15 - 0.22) |
| Age groups | 60-64 | 25-29 | 0.37 (0.33 - 0.43) | 0.07 (0.06 - 0.09) | 0.06 (0.03 - 0.09) | 0 (0 - 0.01) | 0.26 (0.22 - 0.31) |
| Age groups | 60-64 | 30-34 | 0.45 (0.39 - 0.5) | 0.07 (0.05 - 0.08) | 0.05 (0.03 - 0.07) | 0.01 (0 - 0.01) | 0.33 (0.28 - 0.39) |
| Age groups | 60-64 | 35-39 | 0.44 (0.39 - 0.51) | 0.06 (0.05 - 0.08) | 0.06 (0.04 - 0.08) | 0.01 (0 - 0.01) | 0.31 (0.26 - 0.36) |
| Age groups | 60-64 | 40-44 | 0.43 (0.38 - 0.5) | 0.04 (0.03 - 0.05) | 0.06 (0.04 - 0.09) | 0.01 (0 - 0.02) | 0.34 (0.28 - 0.4) |
| Age groups | 60-64 | 45-49 | 0.36 (0.31 - 0.43) | 0.03 (0.03 - 0.04) | 0.08 (0.06 - 0.12) | 0.02 (0.01 - 0.05) | 0.22 (0.19 - 0.26) |
| Age groups | 60-64 | 50-54 | 0.42 (0.38 - 0.48) | 0.06 (0.05 - 0.07) | 0.07 (0.06 - 0.1) | 0.01 (0 - 0.01) | 0.3 (0.26 - 0.35) |
| Age groups | 60-64 | 55-59 | 0.64 (0.57 - 0.73) | 0.13 (0.12 - 0.15) | 0.08 (0.06 - 0.12) | 0 (0 - 0.01) | 0.44 (0.38 - 0.53) |
| Age groups | 60-64 | 60-64 | 0.61 (0.53 - 0.71) | 0.2 (0.17 - 0.23) | 0.05 (0.03 - 0.07) | 0.01 (0 - 0.02) | 0.28 (0.23 - 0.33) |
| Age groups | 60-64 | 65-69 | 0.46 (0.38 - 0.57) | 0.11 (0.09 - 0.13) | 0.04 (0.02 - 0.06) | 0 (0 - 0) | 0.29 (0.23 - 0.39) |
| Age groups | 60-64 | 70-74 | 0.27 (0.23 - 0.32) | 0.05 (0.04 - 0.06) | 0.03 (0.01 - 0.05) | 0 (0 - 0) | 0.2 (0.16 - 0.24) |
| Age groups | 60-64 | 75+ | 0.54 (0.45 - 0.63) | 0.05 (0.04 - 0.07) | 0.01 (0 - 0.02) | 0 (0 - 0.01) | 0.61 (0.49 - 0.73) |
| Age groups | 65-69 | 0-4 | 0.26 (0.19 - 0.35) | 0.03 (0.02 - 0.05) | 0 (0 - 0) | 0 (0 - 0.01) | 0.25 (0.16 - 0.35) |
| Age groups | 65-69 | 5-9 | 0.3 (0.22 - 0.38) | 0.03 (0.02 - 0.03) | 0.01 (0 - 0.02) | 0.01 (0 - 0.02) | 0.23 (0.17 - 0.29) |
| Age groups | 65-69 | 10-14 | 0.24 (0.18 - 0.3) | 0.04 (0.03 - 0.05) | 0 (0 - 0.01) | 0.01 (0 - 0.01) | 0.15 (0.11 - 0.19) |
| Age groups | 65-69 | 15-19 | 0.37 (0.24 - 0.54) | 0.02 (0.01 - 0.03) | 0.03 (0 - 0.08) | 0.01 (0 - 0.03) | 0.27 (0.17 - 0.4) |
| Age groups | 65-69 | 20-24 | 0.4 (0.29 - 0.54) | 0.02 (0.01 - 0.03) | 0.11 (0.02 - 0.25) | 0.03 (0 - 0.08) | 0.28 (0.2 - 0.37) |
| Age groups | 65-69 | 25-29 | 0.48 (0.36 - 0.63) | 0.03 (0.02 - 0.04) | 0.08 (0.02 - 0.17) | 0 (0 - 0) | 0.46 (0.35 - 0.6) |
| Age groups | 65-69 | 30-34 | 0.66 (0.48 - 0.89) | 0.06 (0.04 - 0.07) | 0.05 (0.02 - 0.08) | 0.01 (0 - 0.02) | 0.29 (0.22 - 0.39) |
| Age groups | 65-69 | 35-39 | 0.49 (0.39 - 0.6) | 0.06 (0.05 - 0.07) | 0.05 (0.02 - 0.1) | 0.02 (0.01 - 0.03) | 0.32 (0.24 - 0.41) |
| Age groups | 65-69 | 40-44 | 0.47 (0.38 - 0.59) | 0.05 (0.04 - 0.07) | 0.04 (0.02 - 0.06) | 0 (0 - 0) | 0.34 (0.25 - 0.45) |
| Age groups | 65-69 | 45-49 | 0.3 (0.25 - 0.38) | 0.04 (0.03 - 0.05) | 0.06 (0.02 - 0.13) | 0.01 (0 - 0.02) | 0.19 (0.15 - 0.25) |
| Age groups | 65-69 | 50-54 | 0.31 (0.25 - 0.38) | 0.03 (0.02 - 0.04) | 0.04 (0.02 - 0.05) | 0 (0 - 0.01) | 0.28 (0.22 - 0.36) |
| Age groups | 65-69 | 55-59 | 0.42 (0.35 - 0.53) | 0.07 (0.05 - 0.08) | 0.04 (0.02 - 0.07) | 0 (0 - 0) | 0.35 (0.27 - 0.45) |
| Age groups | 65-69 | 60-64 | 0.55 (0.45 - 0.68) | 0.13 (0.11 - 0.15) | 0.05 (0.03 - 0.07) | 0 (0 - 0) | 0.35 (0.27 - 0.46) |
| Age groups | 65-69 | 65-69 | 0.87 (0.63 - 1.16) | 0.2 (0.17 - 0.23) | 0.06 (0.01 - 0.12) | 0 (0 - 0) | 0.55 (0.36 - 0.76) |
| Age groups | 65-69 | 70-74 | 0.48 (0.39 - 0.57) | 0.09 (0.07 - 0.1) | 0.04 (0.01 - 0.08) | 0 (0 - 0.01) | 0.33 (0.26 - 0.41) |
| Age groups | 65-69 | 75+ | 0.47 (0.38 - 0.56) | 0.09 (0.07 - 0.12) | 0.06 (0.01 - 0.12) | 0 (0 - 0.01) | 0.37 (0.29 - 0.47) |
| Age groups | 70-74 | 0-4 | 0.29 (0.2 - 0.4) | 0.02 (0.01 - 0.02) | 0 (0 - 0.01) | 0 (0 - 0.02) | 0.25 (0.17 - 0.35) |
| Age groups | 70-74 | 5-9 | 0.18 (0.13 - 0.23) | 0.02 (0.01 - 0.03) | 0 (0 - 0.01) | 0.01 (0 - 0.02) | 0.14 (0.1 - 0.19) |
| Age groups | 70-74 | 10-14 | 0.24 (0.17 - 0.31) | 0.02 (0.01 - 0.03) | 0 (0 - 0) | 0.01 (0 - 0.01) | 0.19 (0.14 - 0.25) |
| Age groups | 70-74 | 15-19 | 0.22 (0.16 - 0.29) | 0.02 (0.02 - 0.03) | 0.01 (0 - 0.03) | 0.01 (0 - 0.02) | 0.2 (0.14 - 0.27) |
| Age groups | 70-74 | 20-24 | 0.25 (0.19 - 0.31) | 0.02 (0.01 - 0.02) | 0.02 (0.01 - 0.03) | 0 (0 - 0) | 0.24 (0.18 - 0.3) |
| Age groups | 70-74 | 25-29 | 0.3 (0.24 - 0.38) | 0.02 (0.01 - 0.03) | 0.07 (0.02 - 0.16) | 0 (0 - 0) | 0.13 (0.11 - 0.16) |
| Age groups | 70-74 | 30-34 | 0.45 (0.33 - 0.62) | 0.03 (0.02 - 0.04) | 0.03 (0.01 - 0.05) | 0 (0 - 0) | 0.43 (0.3 - 0.61) |
| Age groups | 70-74 | 35-39 | 0.31 (0.25 - 0.39) | 0.04 (0.03 - 0.05) | 0.04 (0.01 - 0.08) | 0 (0 - 0) | 0.25 (0.19 - 0.33) |
| Age groups | 70-74 | 40-44 | 0.46 (0.38 - 0.57) | 0.06 (0.04 - 0.07) | 0.03 (0.01 - 0.05) | 0 (0 - 0.01) | 0.37 (0.28 - 0.48) |
| Age groups | 70-74 | 45-49 | 0.39 (0.31 - 0.49) | 0.05 (0.04 - 0.06) | 0.05 (0.02 - 0.11) | 0.01 (0 - 0.02) | 0.31 (0.25 - 0.37) |
| Age groups | 70-74 | 50-54 | 0.36 (0.31 - 0.42) | 0.04 (0.03 - 0.05) | 0.03 (0.01 - 0.05) | 0 (0 - 0.01) | 0.26 (0.22 - 0.3) |
| Age groups | 70-74 | 55-59 | 0.28 (0.24 - 0.33) | 0.04 (0.03 - 0.05) | 0.03 (0.01 - 0.05) | 0 (0 - 0) | 0.25 (0.21 - 0.3) |
| Age groups | 70-74 | 60-64 | 0.34 (0.29 - 0.4) | 0.06 (0.05 - 0.07) | 0.04 (0.02 - 0.07) | 0 (0 - 0) | 0.26 (0.21 - 0.31) |
| Age groups | 70-74 | 65-69 | 0.51 (0.42 - 0.61) | 0.09 (0.08 - 0.11) | 0.04 (0.01 - 0.08) | 0 (0 - 0.01) | 0.35 (0.28 - 0.44) |
| Age groups | 70-74 | 70-74 | 0.59 (0.49 - 0.73) | 0.21 (0.17 - 0.24) | 0.03 (0 - 0.06) | 0 (0 - 0) | 0.31 (0.24 - 0.41) |
| Age groups | 70-74 | 75+ | 0.75 (0.63 - 0.87) | 0.14 (0.11 - 0.17) | 0.05 (0.01 - 0.09) | 0.01 (0 - 0.02) | 0.44 (0.36 - 0.54) |
| Age groups | 75+ | 0-4 | 0.07 (0.05 - 0.1) | 0.01 (0 - 0.02) | 0 (0 - 0) | 0 (0 - 0) | 0.07 (0.05 - 0.1) |
| Age groups | 75+ | 5-9 | 0.15 (0.11 - 0.19) | 0.01 (0.01 - 0.02) | 0 (0 - 0) | 0.01 (0 - 0.01) | 0.13 (0.1 - 0.17) |
| Age groups | 75+ | 10-14 | 0.12 (0.09 - 0.16) | 0.01 (0.01 - 0.02) | 0 (0 - 0) | 0 (0 - 0.01) | 0.15 (0.11 - 0.19) |
| Age groups | 75+ | 15-19 | 0.14 (0.1 - 0.17) | 0.01 (0.01 - 0.02) | 0 (0 - 0.01) | 0.01 (0 - 0.02) | 0.09 (0.06 - 0.12) |
| Age groups | 75+ | 20-24 | 0.22 (0.16 - 0.3) | 0 (0 - 0.01) | 0.02 (0 - 0.05) | 0 (0 - 0.01) | 0.16 (0.11 - 0.21) |
| Age groups | 75+ | 25-29 | 0.15 (0.13 - 0.19) | 0.02 (0.01 - 0.02) | 0.01 (0 - 0.02) | 0 (0 - 0.01) | 0.13 (0.11 - 0.16) |
| Age groups | 75+ | 30-34 | 0.27 (0.21 - 0.35) | 0.02 (0.01 - 0.03) | 0 (0 - 0.01) | 0 (0 - 0) | 0.4 (0.3 - 0.55) |
| Age groups | 75+ | 35-39 | 0.25 (0.21 - 0.3) | 0.02 (0.02 - 0.04) | 0.02 (0 - 0.04) | 0 (0 - 0.01) | 0.22 (0.18 - 0.26) |
| Age groups | 75+ | 40-44 | 0.34 (0.28 - 0.42) | 0.03 (0.02 - 0.05) | 0.01 (0 - 0.01) | 0 (0 - 0.01) | 0.27 (0.21 - 0.34) |
| Age groups | 75+ | 45-49 | 0.36 (0.29 - 0.46) | 0.05 (0.04 - 0.06) | 0.05 (0.01 - 0.13) | 0.01 (0 - 0.02) | 0.23 (0.19 - 0.28) |
| Age groups | 75+ | 50-54 | 0.41 (0.35 - 0.48) | 0.06 (0.04 - 0.07) | 0.01 (0 - 0.01) | 0 (0 - 0.01) | 0.32 (0.26 - 0.37) |
| Age groups | 75+ | 55-59 | 0.36 (0.3 - 0.42) | 0.05 (0.04 - 0.06) | 0.03 (0 - 0.08) | 0 (0 - 0.01) | 0.21 (0.17 - 0.25) |
| Age groups | 75+ | 60-64 | 0.36 (0.3 - 0.42) | 0.04 (0.03 - 0.05) | 0.01 (0 - 0.01) | 0 (0 - 0.01) | 0.4 (0.33 - 0.49) |
| Age groups | 75+ | 65-69 | 0.26 (0.21 - 0.32) | 0.05 (0.04 - 0.07) | 0.03 (0.01 - 0.07) | 0 (0 - 0.01) | 0.21 (0.16 - 0.26) |
| Age groups | 75+ | 70-74 | 0.39 (0.33 - 0.46) | 0.07 (0.06 - 0.09) | 0.02 (0.01 - 0.05) | 0 (0 - 0.01) | 0.23 (0.19 - 0.28) |
| Age groups | 75+ | 75+ | 1.7 (1.36 - 2.08) | 0.35 (0.31 - 0.4) | 0.04 (0.01 - 0.1) | 0.01 (0 - 0.04) | 1.42 (1.05 - 1.86) |
| Ethnicity groups | White | White | 3.24 (3.18 - 3.3) | 1.23 (1.2 - 1.26) | 0.37 (0.34 - 0.4) | 0.32 (0.29 - 0.35) | 1.32 (1.28 - 1.36) |
| Ethnicity groups | White | Asian | 0.16 (0.15 - 0.16) | 0.02 (0.02 - 0.03) | 0.02 (0.02 - 0.03) | 0.02 (0.02 - 0.02) | 0.09 (0.08 - 0.09) |
| Ethnicity groups | White | Black | 0.12 (0.11 - 0.12) | 0.02 (0.02 - 0.02) | 0.02 (0.02 - 0.03) | 0.02 (0.02 - 0.02) | 0.06 (0.05 - 0.06) |
| Ethnicity groups | White | Mixed | 0.1 (0.09 - 0.11) | 0.02 (0.02 - 0.03) | 0.01 (0.01 - 0.01) | 0.02 (0.01 - 0.02) | 0.04 (0.04 - 0.05) |
| Ethnicity groups | White | Other | 0.03 (0.02 - 0.03) | 0.01 (0.01 - 0.01) | 0 (0 - 0.01) | 0 (0 - 0) | 0.01 (0.01 - 0.02) |
| Ethnicity groups | Asian | White | 1.37 (1.3 - 1.44) | 0.22 (0.19 - 0.24) | 0.21 (0.18 - 0.24) | 0.18 (0.15 - 0.21) | 0.77 (0.72 - 0.81) |
| Ethnicity groups | Asian | Asian | 1.72 (1.6 - 1.84) | 0.87 (0.79 - 0.95) | 0.14 (0.11 - 0.17) | 0.17 (0.13 - 0.21) | 0.54 (0.48 - 0.62) |
| Ethnicity groups | Asian | Black | 0.18 (0.16 - 0.2) | 0.03 (0.02 - 0.04) | 0.03 (0.03 - 0.04) | 0.03 (0.02 - 0.03) | 0.09 (0.08 - 0.1) |
| Ethnicity groups | Asian | Mixed | 0.17 (0.15 - 0.2) | 0.05 (0.04 - 0.06) | 0.02 (0.02 - 0.03) | 0.02 (0.01 - 0.03) | 0.08 (0.06 - 0.09) |
| Ethnicity groups | Asian | Other | 0.07 (0.05 - 0.1) | 0.01 (0.01 - 0.02) | 0.01 (0 - 0.02) | 0.02 (0 - 0.03) | 0.04 (0.02 - 0.05) |
| Ethnicity groups | Black | White | 2.42 (2.3 - 2.54) | 0.42 (0.38 - 0.47) | 0.48 (0.42 - 0.54) | 0.39 (0.34 - 0.43) | 1.13 (1.06 - 1.21) |
| Ethnicity groups | Black | Asian | 0.42 (0.37 - 0.47) | 0.06 (0.05 - 0.08) | 0.08 (0.06 - 0.1) | 0.06 (0.05 - 0.08) | 0.21 (0.19 - 0.24) |
| Ethnicity groups | Black | Black | 2.22 (2.09 - 2.36) | 1.1 (1.03 - 1.18) | 0.2 (0.17 - 0.24) | 0.19 (0.16 - 0.23) | 0.72 (0.65 - 0.82) |
| Ethnicity groups | Black | Mixed | 0.37 (0.32 - 0.42) | 0.09 (0.08 - 0.12) | 0.04 (0.03 - 0.05) | 0.07 (0.05 - 0.09) | 0.17 (0.14 - 0.2) |
| Ethnicity groups | Black | Other | 0.03 (0.02 - 0.05) | 0.01 (0 - 0.01) | 0 (0 - 0.01) | 0 (0 - 0.01) | 0.02 (0.01 - 0.03) |
| Ethnicity groups | Mixed | White | 2.78 (2.58 - 3) | 0.68 (0.6 - 0.75) | 0.34 (0.28 - 0.4) | 0.52 (0.41 - 0.64) | 1.26 (1.14 - 1.38) |
| Ethnicity groups | Mixed | Asian | 0.55 (0.47 - 0.64) | 0.17 (0.13 - 0.21) | 0.07 (0.05 - 0.1) | 0.07 (0.05 - 0.1) | 0.24 (0.2 - 0.29) |
| Ethnicity groups | Mixed | Black | 0.51 (0.45 - 0.58) | 0.13 (0.1 - 0.16) | 0.06 (0.04 - 0.07) | 0.09 (0.07 - 0.12) | 0.23 (0.19 - 0.28) |
| Ethnicity groups | Mixed | Mixed | 1.12 (0.96 - 1.31) | 0.6 (0.5 - 0.71) | 0.09 (0.05 - 0.14) | 0.09 (0.05 - 0.13) | 0.33 (0.25 - 0.41) |
| Ethnicity groups | Mixed | Other | 0.18 (0.12 - 0.24) | 0.09 (0.05 - 0.14) | 0.02 (0 - 0.04) | 0.01 (0 - 0.02) | 0.06 (0.03 - 0.09) |
| Ethnicity groups | Other | White | 1.08 (0.92 - 1.25) | 0.3 (0.22 - 0.39) | 0.15 (0.08 - 0.23) | 0.11 (0.05 - 0.18) | 0.52 (0.43 - 0.63) |
| Ethnicity groups | Other | Asian | 0.33 (0.23 - 0.43) | 0.05 (0.02 - 0.09) | 0.04 (0.01 - 0.08) | 0.07 (0.02 - 0.13) | 0.17 (0.1 - 0.24) |
| Ethnicity groups | Other | Black | 0.07 (0.04 - 0.1) | 0.01 (0.01 - 0.02) | 0.01 (0 - 0.01) | 0.01 (0 - 0.03) | 0.04 (0.02 - 0.06) |
| Ethnicity groups | Other | Mixed | 0.24 (0.16 - 0.34) | 0.13 (0.07 - 0.19) | 0.03 (0 - 0.06) | 0.01 (0 - 0.03) | 0.08 (0.04 - 0.13) |
| Ethnicity groups | Other | Other | 0.98 (0.63 - 1.35) | 0.56 (0.31 - 0.85) | 0.04 (0 - 0.1) | 0.03 (0 - 0.06) | 0.35 (0.19 - 0.54) |
| NS-SEC classes | 1 | 1 | 0.59 (0.54 - 0.64) | 0.15 (0.13 - 0.18) | 0.28 (0.24 - 0.33) | 0.02 (0.01 - 0.02) | 0.13 (0.11 - 0.15) |
| NS-SEC classes | 1 | 2 | 0.7 (0.64 - 0.76) | 0.2 (0.18 - 0.22) | 0.24 (0.2 - 0.28) | 0.05 (0.03 - 0.06) | 0.22 (0.19 - 0.24) |
| NS-SEC classes | 1 | 3 | 0.25 (0.22 - 0.29) | 0.07 (0.05 - 0.08) | 0.11 (0.08 - 0.13) | 0.01 (0 - 0.02) | 0.07 (0.06 - 0.09) |
| NS-SEC classes | 1 | 4 | 0.17 (0.15 - 0.2) | 0.04 (0.03 - 0.05) | 0.02 (0.01 - 0.03) | 0 (0 - 0.01) | 0.11 (0.1 - 0.14) |
| NS-SEC classes | 1 | 5 | 0.07 (0.05 - 0.08) | 0.02 (0.01 - 0.03) | 0.03 (0.02 - 0.04) | 0 (0 - 0) | 0.02 (0.01 - 0.03) |
| NS-SEC classes | 1 | 6 | 0.39 (0.35 - 0.43) | 0.06 (0.05 - 0.07) | 0.06 (0.05 - 0.08) | 0 (0 - 0.01) | 0.27 (0.24 - 0.31) |
| NS-SEC classes | 1 | 7 | 0.28 (0.25 - 0.31) | 0.05 (0.04 - 0.06) | 0.05 (0.03 - 0.07) | 0 (0 - 0.01) | 0.19 (0.16 - 0.22) |
| NS-SEC classes | 1 | Under 17 | 0.62 (0.56 - 0.68) | 0.42 (0.38 - 0.46) | 0.04 (0.03 - 0.07) | 0.03 (0.02 - 0.05) | 0.1 (0.08 - 0.12) |
| NS-SEC classes | 1 | Student | 0.23 (0.2 - 0.26) | 0.09 (0.08 - 0.11) | 0.03 (0.02 - 0.04) | 0.03 (0.02 - 0.04) | 0.08 (0.06 - 0.09) |
| NS-SEC classes | 1 | Retired | 0.14 (0.12 - 0.17) | 0.05 (0.04 - 0.06) | 0.01 (0 - 0.02) | 0 (0 - 0) | 0.08 (0.07 - 0.1) |
| NS-SEC classes | 1 | Unemployed | 0.29 (0.25 - 0.32) | 0.12 (0.1 - 0.14) | 0.04 (0.03 - 0.05) | 0.01 (0 - 0.01) | 0.12 (0.1 - 0.14) |
| NS-SEC classes | 2 | 1 | 0.32 (0.29 - 0.36) | 0.1 (0.08 - 0.12) | 0.12 (0.1 - 0.15) | 0.02 (0.01 - 0.02) | 0.08 (0.07 - 0.1) |
| NS-SEC classes | 2 | 2 | 0.92 (0.86 - 0.99) | 0.22 (0.2 - 0.24) | 0.33 (0.29 - 0.38) | 0.13 (0.1 - 0.16) | 0.23 (0.2 - 0.26) |
| NS-SEC classes | 2 | 3 | 0.34 (0.31 - 0.38) | 0.08 (0.07 - 0.1) | 0.13 (0.1 - 0.15) | 0.04 (0.02 - 0.06) | 0.09 (0.08 - 0.11) |
| NS-SEC classes | 2 | 4 | 0.21 (0.19 - 0.24) | 0.05 (0.04 - 0.07) | 0.03 (0.02 - 0.04) | 0.01 (0 - 0.01) | 0.13 (0.11 - 0.15) |
| NS-SEC classes | 2 | 5 | 0.07 (0.06 - 0.09) | 0.03 (0.02 - 0.04) | 0.01 (0.01 - 0.02) | 0 (0 - 0) | 0.03 (0.02 - 0.04) |
| NS-SEC classes | 2 | 6 | 0.49 (0.45 - 0.53) | 0.08 (0.06 - 0.09) | 0.09 (0.07 - 0.11) | 0.01 (0.01 - 0.02) | 0.32 (0.29 - 0.35) |
| NS-SEC classes | 2 | 7 | 0.31 (0.28 - 0.35) | 0.07 (0.06 - 0.09) | 0.05 (0.03 - 0.07) | 0 (0 - 0.01) | 0.19 (0.17 - 0.22) |
| NS-SEC classes | 2 | Under 17 | 0.62 (0.56 - 0.69) | 0.35 (0.32 - 0.39) | 0.04 (0.02 - 0.06) | 0.07 (0.03 - 0.12) | 0.14 (0.11 - 0.18) |
| NS-SEC classes | 2 | Student | 0.25 (0.22 - 0.28) | 0.13 (0.11 - 0.15) | 0.02 (0.01 - 0.03) | 0.02 (0.01 - 0.04) | 0.07 (0.05 - 0.09) |
| NS-SEC classes | 2 | Retired | 0.19 (0.17 - 0.21) | 0.07 (0.05 - 0.08) | 0.01 (0 - 0.01) | 0 (0 - 0) | 0.12 (0.1 - 0.14) |
| NS-SEC classes | 2 | Unemployed | 0.34 (0.3 - 0.38) | 0.13 (0.11 - 0.15) | 0.03 (0.02 - 0.04) | 0.01 (0 - 0.01) | 0.17 (0.15 - 0.2) |
| NS-SEC classes | 3 | 1 | 0.28 (0.24 - 0.32) | 0.08 (0.06 - 0.09) | 0.12 (0.09 - 0.15) | 0.01 (0 - 0.02) | 0.07 (0.06 - 0.09) |
| NS-SEC classes | 3 | 2 | 0.56 (0.5 - 0.62) | 0.13 (0.11 - 0.16) | 0.19 (0.15 - 0.23) | 0.07 (0.05 - 0.1) | 0.16 (0.14 - 0.19) |
| NS-SEC classes | 3 | 3 | 0.62 (0.55 - 0.69) | 0.13 (0.11 - 0.16) | 0.31 (0.25 - 0.37) | 0.05 (0.03 - 0.08) | 0.12 (0.1 - 0.15) |
| NS-SEC classes | 3 | 4 | 0.17 (0.14 - 0.2) | 0.06 (0.04 - 0.07) | 0.02 (0.01 - 0.03) | 0 (0 - 0.01) | 0.09 (0.08 - 0.12) |
| NS-SEC classes | 3 | 5 | 0.07 (0.06 - 0.09) | 0.02 (0.02 - 0.03) | 0.02 (0.01 - 0.03) | 0 (0 - 0) | 0.03 (0.02 - 0.04) |
| NS-SEC classes | 3 | 6 | 0.47 (0.42 - 0.52) | 0.09 (0.07 - 0.1) | 0.07 (0.05 - 0.1) | 0.01 (0 - 0.01) | 0.32 (0.28 - 0.36) |
| NS-SEC classes | 3 | 7 | 0.29 (0.26 - 0.33) | 0.09 (0.07 - 0.1) | 0.04 (0.02 - 0.05) | 0 (0 - 0) | 0.18 (0.14 - 0.21) |
| NS-SEC classes | 3 | Under 17 | 0.54 (0.47 - 0.61) | 0.31 (0.27 - 0.35) | 0.05 (0.02 - 0.08) | 0.05 (0.02 - 0.08) | 0.13 (0.1 - 0.16) |
| NS-SEC classes | 3 | Student | 0.22 (0.18 - 0.26) | 0.11 (0.09 - 0.13) | 0.02 (0.01 - 0.03) | 0.04 (0.02 - 0.07) | 0.04 (0.03 - 0.06) |
| NS-SEC classes | 3 | Retired | 0.22 (0.19 - 0.25) | 0.06 (0.05 - 0.08) | 0.02 (0.01 - 0.03) | 0 (0 - 0) | 0.13 (0.11 - 0.16) |
| NS-SEC classes | 3 | Unemployed | 0.28 (0.24 - 0.32) | 0.12 (0.1 - 0.14) | 0.04 (0.02 - 0.05) | 0.01 (0.01 - 0.03) | 0.11 (0.09 - 0.13) |
| NS-SEC classes | 4 | 1 | 0.17 (0.12 - 0.21) | 0.06 (0.04 - 0.09) | 0.04 (0.02 - 0.06) | 0.01 (0 - 0.02) | 0.06 (0.03 - 0.09) |
| NS-SEC classes | 4 | 2 | 0.4 (0.32 - 0.49) | 0.12 (0.08 - 0.17) | 0.09 (0.05 - 0.13) | 0.03 (0.01 - 0.06) | 0.16 (0.11 - 0.21) |
| NS-SEC classes | 4 | 3 | 0.16 (0.11 - 0.21) | 0.07 (0.04 - 0.1) | 0.02 (0.01 - 0.05) | 0 (0 - 0.01) | 0.06 (0.04 - 0.09) |
| NS-SEC classes | 4 | 4 | 0.38 (0.3 - 0.46) | 0.13 (0.09 - 0.17) | 0.1 (0.06 - 0.15) | 0.01 (0 - 0.03) | 0.13 (0.09 - 0.18) |
| NS-SEC classes | 4 | 5 | 0.08 (0.05 - 0.11) | 0.03 (0.01 - 0.04) | 0.02 (0.01 - 0.04) | 0 (0 - 0.01) | 0.03 (0.01 - 0.06) |
| NS-SEC classes | 4 | 6 | 0.45 (0.37 - 0.54) | 0.08 (0.05 - 0.11) | 0.07 (0.04 - 0.11) | 0 (0 - 0) | 0.32 (0.25 - 0.38) |
| NS-SEC classes | 4 | 7 | 0.25 (0.2 - 0.32) | 0.08 (0.05 - 0.11) | 0.05 (0.02 - 0.08) | 0 (0 - 0.01) | 0.13 (0.09 - 0.18) |
| NS-SEC classes | 4 | Under 17 | 0.49 (0.39 - 0.59) | 0.33 (0.25 - 0.43) | 0.03 (0.01 - 0.06) | 0.01 (0 - 0.03) | 0.1 (0.07 - 0.14) |
| NS-SEC classes | 4 | Student | 0.21 (0.15 - 0.27) | 0.13 (0.08 - 0.17) | 0.02 (0.01 - 0.04) | 0.01 (0 - 0.02) | 0.04 (0.02 - 0.07) |
| NS-SEC classes | 4 | Retired | 0.23 (0.18 - 0.29) | 0.06 (0.04 - 0.1) | 0.02 (0.01 - 0.03) | 0 (0 - 0) | 0.16 (0.1 - 0.21) |
| NS-SEC classes | 4 | Unemployed | 0.35 (0.28 - 0.43) | 0.14 (0.1 - 0.18) | 0.04 (0.02 - 0.07) | 0.02 (0 - 0.04) | 0.16 (0.11 - 0.2) |
| NS-SEC classes | 5 | 1 | 0.22 (0.15 - 0.28) | 0.05 (0.03 - 0.08) | 0.07 (0.04 - 0.11) | 0 (0 - 0) | 0.1 (0.06 - 0.14) |
| NS-SEC classes | 5 | 2 | 0.46 (0.35 - 0.56) | 0.13 (0.08 - 0.19) | 0.17 (0.11 - 0.24) | 0.02 (0 - 0.05) | 0.14 (0.09 - 0.19) |
| NS-SEC classes | 5 | 3 | 0.26 (0.18 - 0.34) | 0.09 (0.05 - 0.13) | 0.1 (0.04 - 0.15) | 0 (0 - 0.02) | 0.07 (0.03 - 0.1) |
| NS-SEC classes | 5 | 4 | 0.21 (0.13 - 0.29) | 0.03 (0.01 - 0.06) | 0.07 (0.02 - 0.14) | 0 (0 - 0) | 0.11 (0.06 - 0.16) |
| NS-SEC classes | 5 | 5 | 0.33 (0.24 - 0.44) | 0.05 (0.02 - 0.08) | 0.21 (0.13 - 0.32) | 0 (0 - 0) | 0.08 (0.04 - 0.12) |
| NS-SEC classes | 5 | 6 | 0.4 (0.31 - 0.49) | 0.09 (0.05 - 0.13) | 0.11 (0.05 - 0.18) | 0 (0 - 0) | 0.2 (0.14 - 0.27) |
| NS-SEC classes | 5 | 7 | 0.41 (0.31 - 0.53) | 0.06 (0.03 - 0.09) | 0.18 (0.1 - 0.28) | 0.01 (0 - 0.02) | 0.18 (0.12 - 0.24) |
| NS-SEC classes | 5 | Under 17 | 0.39 (0.29 - 0.51) | 0.23 (0.15 - 0.31) | 0.03 (0.01 - 0.06) | 0.03 (0.01 - 0.06) | 0.1 (0.06 - 0.15) |
| NS-SEC classes | 5 | Student | 0.17 (0.12 - 0.23) | 0.08 (0.04 - 0.12) | 0.03 (0 - 0.06) | 0.02 (0 - 0.04) | 0.05 (0.02 - 0.08) |
| NS-SEC classes | 5 | Retired | 0.13 (0.07 - 0.21) | 0.02 (0.01 - 0.04) | 0.04 (0 - 0.12) | 0 (0 - 0) | 0.07 (0.03 - 0.11) |
| NS-SEC classes | 5 | Unemployed | 0.28 (0.2 - 0.36) | 0.12 (0.07 - 0.16) | 0.04 (0.01 - 0.06) | 0 (0 - 0) | 0.13 (0.08 - 0.19) |
| NS-SEC classes | 6 | 1 | 0.16 (0.12 - 0.19) | 0.05 (0.04 - 0.07) | 0.04 (0.02 - 0.06) | 0.01 (0 - 0.02) | 0.05 (0.04 - 0.08) |
| NS-SEC classes | 6 | 2 | 0.43 (0.37 - 0.5) | 0.09 (0.07 - 0.11) | 0.17 (0.13 - 0.22) | 0.07 (0.04 - 0.11) | 0.11 (0.08 - 0.13) |
| NS-SEC classes | 6 | 3 | 0.27 (0.21 - 0.34) | 0.06 (0.04 - 0.08) | 0.1 (0.06 - 0.14) | 0.03 (0 - 0.07) | 0.08 (0.06 - 0.11) |
| NS-SEC classes | 6 | 4 | 0.22 (0.15 - 0.29) | 0.05 (0.04 - 0.07) | 0.06 (0.02 - 0.11) | 0.01 (0 - 0.02) | 0.1 (0.08 - 0.13) |
| NS-SEC classes | 6 | 5 | 0.09 (0.07 - 0.12) | 0.04 (0.02 - 0.05) | 0.03 (0.01 - 0.05) | 0 (0 - 0) | 0.03 (0.01 - 0.04) |
| NS-SEC classes | 6 | 6 | 0.76 (0.67 - 0.87) | 0.12 (0.1 - 0.15) | 0.28 (0.21 - 0.35) | 0.02 (0 - 0.06) | 0.35 (0.3 - 0.4) |
| NS-SEC classes | 6 | 7 | 0.35 (0.3 - 0.4) | 0.12 (0.09 - 0.15) | 0.05 (0.03 - 0.08) | 0 (0 - 0.01) | 0.17 (0.14 - 0.21) |
| NS-SEC classes | 6 | Under 17 | 0.44 (0.36 - 0.53) | 0.26 (0.21 - 0.31) | 0.02 (0.01 - 0.05) | 0.06 (0.01 - 0.11) | 0.09 (0.06 - 0.12) |
| NS-SEC classes | 6 | Student | 0.23 (0.19 - 0.28) | 0.09 (0.07 - 0.11) | 0.03 (0.02 - 0.06) | 0.04 (0.02 - 0.07) | 0.07 (0.04 - 0.09) |
| NS-SEC classes | 6 | Retired | 0.26 (0.21 - 0.31) | 0.06 (0.04 - 0.07) | 0.02 (0 - 0.03) | 0 (0 - 0.01) | 0.19 (0.16 - 0.24) |
| NS-SEC classes | 6 | Unemployed | 0.33 (0.28 - 0.39) | 0.14 (0.11 - 0.18) | 0.04 (0.02 - 0.06) | 0.01 (0 - 0.02) | 0.14 (0.11 - 0.18) |
| NS-SEC classes | 7 | 1 | 0.15 (0.11 - 0.2) | 0.03 (0.01 - 0.04) | 0.07 (0.04 - 0.11) | 0.01 (0 - 0.02) | 0.05 (0.03 - 0.08) |
| NS-SEC classes | 7 | 2 | 0.35 (0.29 - 0.42) | 0.1 (0.07 - 0.13) | 0.13 (0.08 - 0.18) | 0.02 (0.01 - 0.04) | 0.1 (0.08 - 0.14) |
| NS-SEC classes | 7 | 3 | 0.16 (0.12 - 0.21) | 0.05 (0.03 - 0.07) | 0.05 (0.02 - 0.08) | 0.01 (0 - 0.01) | 0.05 (0.04 - 0.08) |
| NS-SEC classes | 7 | 4 | 0.18 (0.14 - 0.23) | 0.04 (0.03 - 0.06) | 0.02 (0.01 - 0.05) | 0 (0 - 0) | 0.12 (0.09 - 0.16) |
| NS-SEC classes | 7 | 5 | 0.12 (0.09 - 0.16) | 0.03 (0.01 - 0.05) | 0.07 (0.04 - 0.1) | 0 (0 - 0) | 0.03 (0.01 - 0.04) |
| NS-SEC classes | 7 | 6 | 0.39 (0.33 - 0.47) | 0.07 (0.05 - 0.09) | 0.09 (0.05 - 0.14) | 0 (0 - 0.01) | 0.24 (0.19 - 0.29) |
| NS-SEC classes | 7 | 7 | 0.6 (0.49 - 0.72) | 0.12 (0.08 - 0.15) | 0.3 (0.21 - 0.4) | 0.02 (0 - 0.04) | 0.17 (0.13 - 0.22) |
| NS-SEC classes | 7 | Under 17 | 0.46 (0.36 - 0.57) | 0.29 (0.23 - 0.36) | 0.07 (0.02 - 0.15) | 0.01 (0 - 0.02) | 0.07 (0.04 - 0.11) |
| NS-SEC classes | 7 | Student | 0.26 (0.2 - 0.33) | 0.1 (0.07 - 0.14) | 0.02 (0.01 - 0.04) | 0.05 (0.03 - 0.09) | 0.07 (0.05 - 0.11) |
| NS-SEC classes | 7 | Retired | 0.21 (0.16 - 0.27) | 0.07 (0.05 - 0.09) | 0.02 (0 - 0.04) | 0 (0 - 0) | 0.13 (0.09 - 0.18) |
| NS-SEC classes | 7 | Unemployed | 0.37 (0.31 - 0.45) | 0.17 (0.13 - 0.21) | 0.04 (0.02 - 0.08) | 0.01 (0 - 0.02) | 0.15 (0.11 - 0.19) |
| NS-SEC classes | Under 17 | 1 | 0.27 (0.25 - 0.29) | 0.17 (0.15 - 0.18) | 0 (0 - 0) | 0.01 (0.01 - 0.01) | 0.09 (0.08 - 0.11) |
| NS-SEC classes | Under 17 | 2 | 0.8 (0.76 - 0.84) | 0.29 (0.27 - 0.32) | 0.01 (0 - 0.01) | 0.24 (0.22 - 0.26) | 0.26 (0.24 - 0.28) |
| NS-SEC classes | Under 17 | 3 | 0.26 (0.24 - 0.29) | 0.12 (0.11 - 0.14) | 0 (0 - 0.01) | 0.03 (0.02 - 0.04) | 0.11 (0.09 - 0.13) |
| NS-SEC classes | Under 17 | 4 | 0.13 (0.12 - 0.15) | 0.05 (0.04 - 0.06) | 0 (0 - 0) | 0 (0 - 0.01) | 0.08 (0.07 - 0.09) |
| NS-SEC classes | Under 17 | 5 | 0.05 (0.04 - 0.06) | 0.03 (0.02 - 0.04) | 0 (0 - 0) | 0 (0 - 0) | 0.02 (0.01 - 0.02) |
| NS-SEC classes | Under 17 | 6 | 0.3 (0.28 - 0.32) | 0.11 (0.1 - 0.12) | 0 (0 - 0.01) | 0.02 (0.01 - 0.02) | 0.18 (0.16 - 0.2) |
| NS-SEC classes | Under 17 | 7 | 0.21 (0.19 - 0.23) | 0.08 (0.07 - 0.09) | 0 (0 - 0) | 0.02 (0.01 - 0.02) | 0.11 (0.1 - 0.13) |
| NS-SEC classes | Under 17 | Under 17 | 2.24 (2.13 - 2.36) | 0.63 (0.59 - 0.66) | 0.01 (0.01 - 0.02) | 0.96 (0.88 - 1.05) | 0.62 (0.57 - 0.67) |
| NS-SEC classes | Under 17 | Student | 0.28 (0.25 - 0.31) | 0.1 (0.09 - 0.11) | 0 (0 - 0.01) | 0.06 (0.05 - 0.08) | 0.12 (0.1 - 0.14) |
| NS-SEC classes | Under 17 | Retired | 0.1 (0.09 - 0.12) | 0.04 (0.03 - 0.05) | 0 (0 - 0) | 0 (0 - 0) | 0.07 (0.06 - 0.08) |
| NS-SEC classes | Under 17 | Unemployed | 0.37 (0.34 - 0.4) | 0.22 (0.2 - 0.24) | 0 (0 - 0) | 0.01 (0.01 - 0.01) | 0.14 (0.12 - 0.16) |
| NS-SEC classes | Student | 1 | 0.1 (0.05 - 0.16) | 0.05 (0.02 - 0.09) | 0 (0 - 0) | 0.02 (0 - 0.05) | 0.03 (0.01 - 0.06) |
| NS-SEC classes | Student | 2 | 0.37 (0.26 - 0.49) | 0.18 (0.11 - 0.25) | 0.01 (0 - 0.02) | 0.1 (0.05 - 0.17) | 0.07 (0.02 - 0.13) |
| NS-SEC classes | Student | 3 | 0.15 (0.08 - 0.23) | 0.04 (0.02 - 0.08) | 0.03 (0 - 0.07) | 0.01 (0 - 0.03) | 0.07 (0.03 - 0.13) |
| NS-SEC classes | Student | 4 | 0.12 (0.07 - 0.17) | 0.06 (0.03 - 0.1) | 0 (0 - 0) | 0.01 (0 - 0.04) | 0.04 (0.01 - 0.07) |
| NS-SEC classes | Student | 5 | 0.01 (0 - 0.02) | 0 (0 - 0) | 0 (0 - 0) | 0 (0 - 0) | 0.01 (0 - 0.02) |
| NS-SEC classes | Student | 6 | 0.45 (0.33 - 0.62) | 0.12 (0.07 - 0.17) | 0.01 (0 - 0.02) | 0.02 (0 - 0.04) | 0.34 (0.21 - 0.49) |
| NS-SEC classes | Student | 7 | 0.25 (0.17 - 0.33) | 0.06 (0.02 - 0.1) | 0 (0 - 0) | 0.02 (0 - 0.04) | 0.19 (0.11 - 0.27) |
| NS-SEC classes | Student | Under 17 | 0.37 (0.22 - 0.55) | 0.19 (0.12 - 0.28) | 0 (0 - 0) | 0.09 (0.01 - 0.2) | 0.08 (0.02 - 0.18) |
| NS-SEC classes | Student | Student | 1.24 (0.98 - 1.53) | 0.38 (0.25 - 0.54) | 0 (0 - 0) | 0.54 (0.35 - 0.78) | 0.3 (0.2 - 0.43) |
| NS-SEC classes | Student | Retired | 0.02 (0.01 - 0.05) | 0.01 (0 - 0.03) | 0 (0 - 0) | 0 (0 - 0) | 0.01 (0 - 0.04) |
| NS-SEC classes | Student | Unemployed | 0.38 (0.28 - 0.5) | 0.22 (0.15 - 0.3) | 0 (0 - 0) | 0.02 (0 - 0.06) | 0.13 (0.07 - 0.2) |
| NS-SEC classes | Retired | 1 | 0.14 (0.12 - 0.16) | 0.04 (0.03 - 0.05) | 0 (0 - 0.01) | 0 (0 - 0) | 0.1 (0.08 - 0.11) |
| NS-SEC classes | Retired | 2 | 0.33 (0.3 - 0.37) | 0.09 (0.07 - 0.11) | 0.02 (0.01 - 0.03) | 0.01 (0 - 0.02) | 0.22 (0.19 - 0.25) |
| NS-SEC classes | Retired | 3 | 0.18 (0.16 - 0.21) | 0.07 (0.06 - 0.08) | 0 (0 - 0.01) | 0 (0 - 0.01) | 0.1 (0.09 - 0.12) |
| NS-SEC classes | Retired | 4 | 0.15 (0.13 - 0.17) | 0.04 (0.03 - 0.05) | 0 (0 - 0) | 0 (0 - 0) | 0.11 (0.09 - 0.13) |
| NS-SEC classes | Retired | 5 | 0.05 (0.04 - 0.06) | 0.02 (0.01 - 0.03) | 0 (0 - 0) | 0 (0 - 0) | 0.03 (0.02 - 0.04) |
| NS-SEC classes | Retired | 6 | 0.48 (0.44 - 0.52) | 0.06 (0.04 - 0.07) | 0 (0 - 0.01) | 0 (0 - 0) | 0.43 (0.39 - 0.47) |
| NS-SEC classes | Retired | 7 | 0.34 (0.3 - 0.38) | 0.1 (0.08 - 0.11) | 0 (0 - 0) | 0 (0 - 0) | 0.24 (0.21 - 0.28) |
| NS-SEC classes | Retired | Under 17 | 0.2 (0.17 - 0.23) | 0.08 (0.07 - 0.1) | 0 (0 - 0) | 0.01 (0 - 0.01) | 0.1 (0.08 - 0.13) |
| NS-SEC classes | Retired | Student | 0.04 (0.03 - 0.05) | 0.02 (0.01 - 0.03) | 0 (0 - 0) | 0 (0 - 0) | 0.02 (0.01 - 0.03) |
| NS-SEC classes | Retired | Retired | 0.93 (0.87 - 0.99) | 0.36 (0.33 - 0.38) | 0.01 (0 - 0.01) | 0.01 (0 - 0.03) | 0.55 (0.49 - 0.61) |
| NS-SEC classes | Retired | Unemployed | 0.34 (0.3 - 0.37) | 0.15 (0.13 - 0.17) | 0.01 (0 - 0.01) | 0 (0 - 0.01) | 0.18 (0.15 - 0.21) |
| NS-SEC classes | Unemployed | 1 | 0.11 (0.09 - 0.13) | 0.05 (0.04 - 0.06) | 0 (0 - 0) | 0 (0 - 0) | 0.06 (0.05 - 0.08) |
| NS-SEC classes | Unemployed | 2 | 0.27 (0.23 - 0.3) | 0.1 (0.08 - 0.11) | 0.01 (0 - 0.02) | 0.03 (0.02 - 0.05) | 0.13 (0.11 - 0.15) |
| NS-SEC classes | Unemployed | 3 | 0.15 (0.13 - 0.18) | 0.06 (0.05 - 0.08) | 0.01 (0 - 0.01) | 0.01 (0 - 0.02) | 0.08 (0.06 - 0.1) |
| NS-SEC classes | Unemployed | 4 | 0.13 (0.11 - 0.15) | 0.04 (0.03 - 0.06) | 0 (0 - 0.01) | 0 (0 - 0.01) | 0.08 (0.06 - 0.1) |
| NS-SEC classes | Unemployed | 5 | 0.05 (0.03 - 0.06) | 0.03 (0.02 - 0.04) | 0 (0 - 0) | 0 (0 - 0) | 0.02 (0.01 - 0.02) |
| NS-SEC classes | Unemployed | 6 | 0.41 (0.37 - 0.46) | 0.11 (0.09 - 0.13) | 0 (0 - 0.01) | 0 (0 - 0.01) | 0.31 (0.27 - 0.35) |
| NS-SEC classes | Unemployed | 7 | 0.26 (0.22 - 0.3) | 0.1 (0.08 - 0.12) | 0.01 (0 - 0.01) | 0 (0 - 0.01) | 0.16 (0.13 - 0.19) |
| NS-SEC classes | Unemployed | Under 17 | 0.45 (0.39 - 0.5) | 0.32 (0.28 - 0.37) | 0.01 (0.01 - 0.02) | 0.02 (0.01 - 0.03) | 0.09 (0.07 - 0.11) |
| NS-SEC classes | Unemployed | Student | 0.13 (0.11 - 0.15) | 0.08 (0.07 - 0.1) | 0 (0 - 0.01) | 0.01 (0 - 0.01) | 0.04 (0.02 - 0.05) |
| NS-SEC classes | Unemployed | Retired | 0.28 (0.24 - 0.32) | 0.12 (0.1 - 0.14) | 0.01 (0 - 0.01) | 0 (0 - 0) | 0.16 (0.13 - 0.19) |
| NS-SEC classes | Unemployed | Unemployed | 0.47 (0.42 - 0.52) | 0.26 (0.22 - 0.29) | 0.01 (0 - 0.01) | 0.02 (0.01 - 0.03) | 0.18 (0.15 - 0.21) |

**Table S8:** Mean number of daily contacts between individuals of each age group, ethnicity, and NS-SEC class, stratified by setting (Total, Home, Work, School, Other), as estimated by the weighted negative binomial regression model.

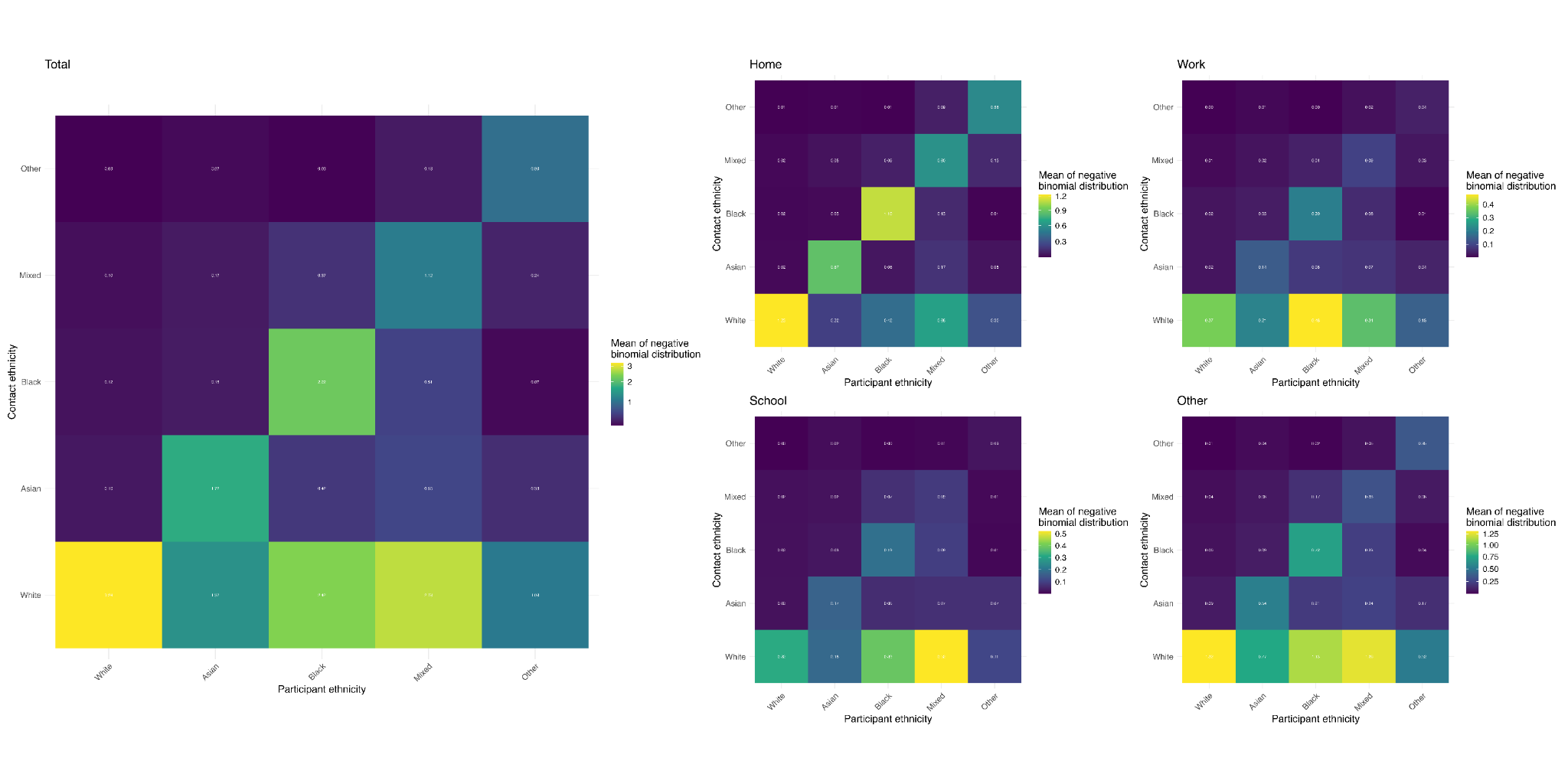

**Figure S8:** Mean number of contacts between individuals of each ethnicity, stratified by setting, as estimated by the weighted negative binomial regression model.

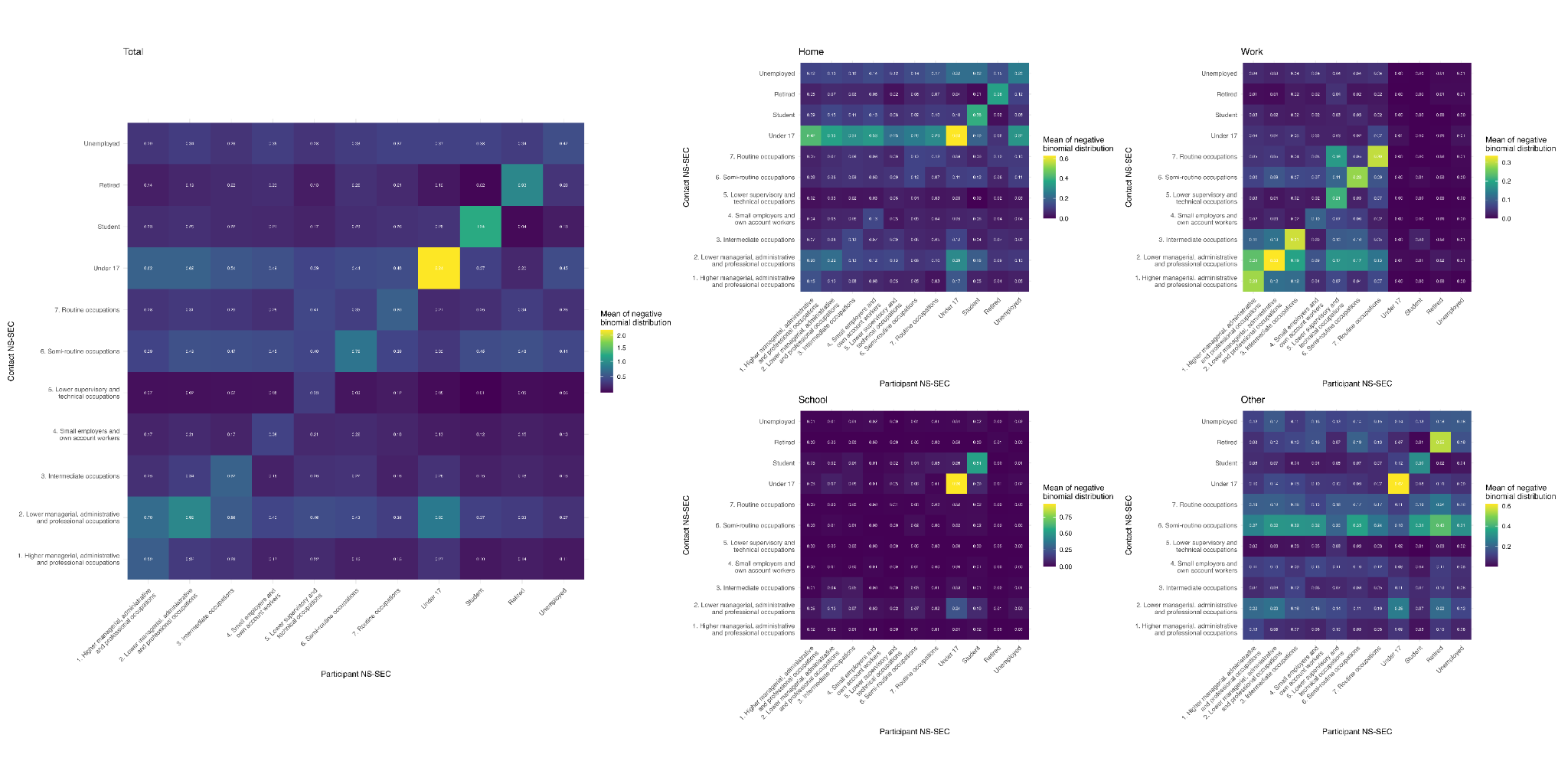

**Figure S9:** Mean number of contacts between individuals of each NS-SEC class, stratified by setting, as estimated by the weighted negative binomial regression model. These contact matrices are not adjusted for population-level reciprocity.

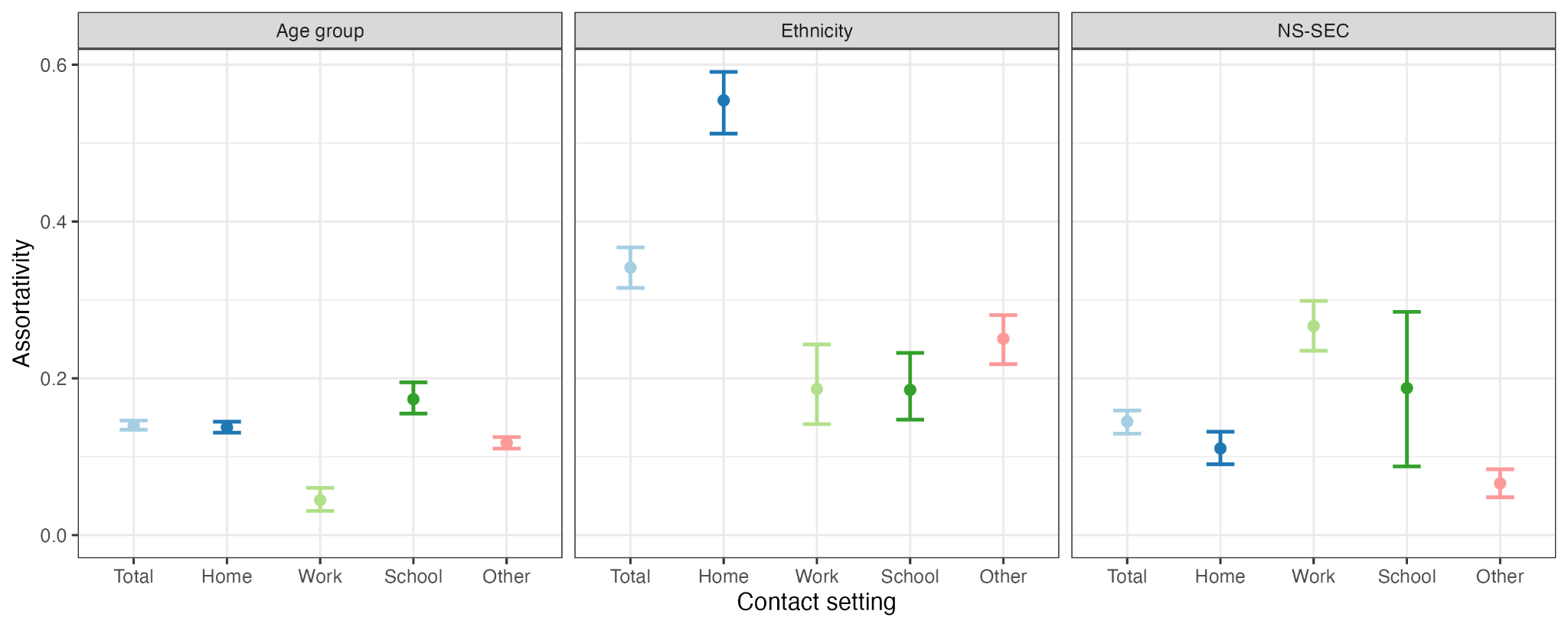

**Figure S10:** Mean assortativity of daily contacts between age groups, ethnic groups, and NS-SEC classes (only estimated using contacts between employed individuals, i.e. NS-SEC classes 1-7), using the assortativity measure *Q* as described by Keeling and Rohani [11], where *Q* = 1 describes complete assortativity, *Q* = 0 proportionate mixing, and *Q* = -1 complete disassortative mixing.

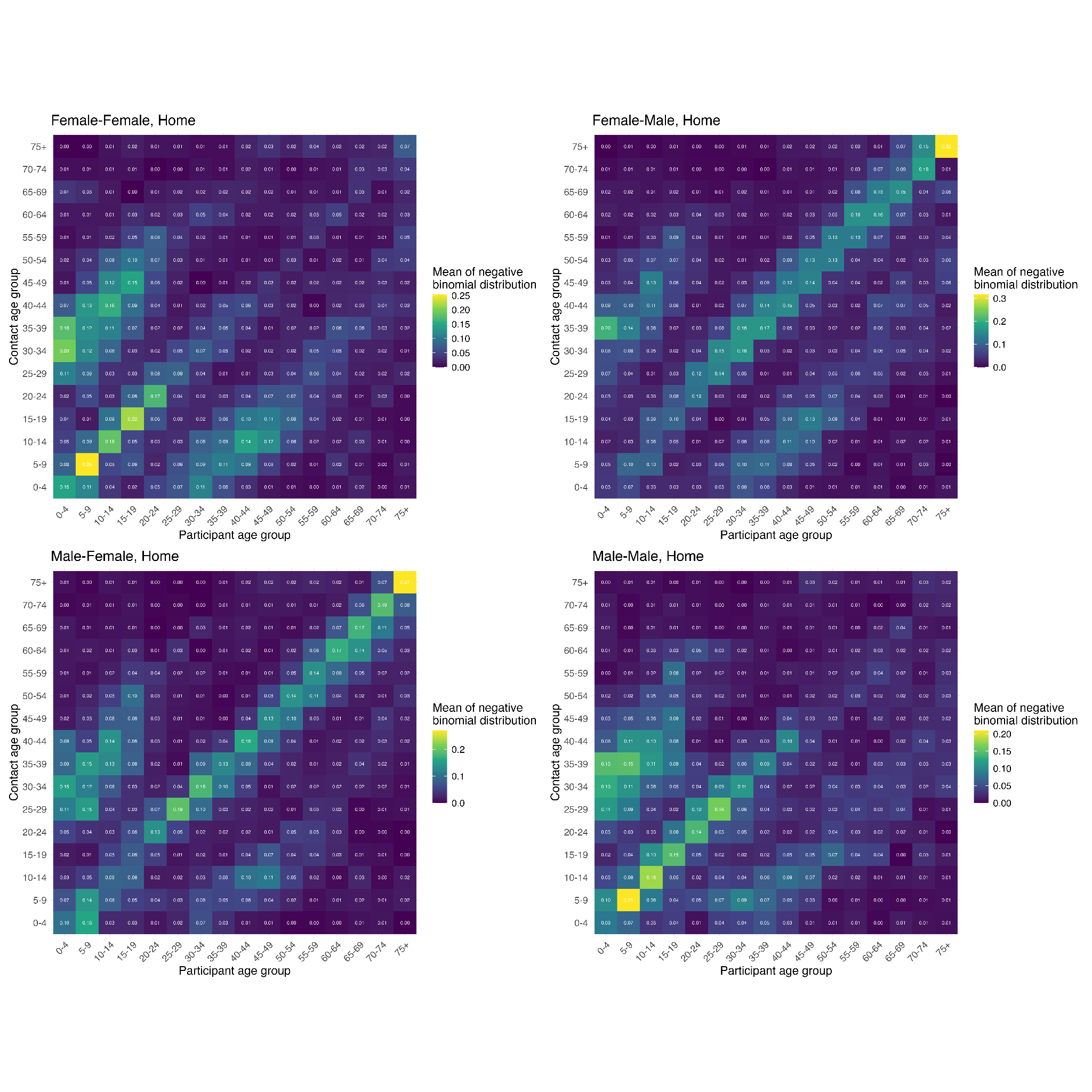

**Figure S11:** Mean number of contacts in the home between individuals of each age group, stratified by gender (restricted to male and female), calculated using the weighted negative binomial regression model (not normalised with respect to age- and gender-specific population structure).

#### g. Effect on infection risk

| **Analysis** | **Group** | **Infection Share (95% CI)** | **Risk Ratio (95% CI)** |
| --- | --- | --- | --- |
| Age only | 0-4 | 6.2% (5.8-6.6) | 1.56 (1.43-1.72) |
| Age only | 5-9 | 9.8% (9.2-10.5) | 2.28 (2.07-2.53) |
| Age only | 10-14 | 7.6% (7.1-8.1) | 1.73 (1.57-1.89) |
| Age only | 15-19 | 5.5% (5.2-5.8) | 1.32 (1.22-1.44) |
| Age only | 20-24 | 5.9% (5.6-6.2) | 1.34 (1.24-1.45) |
| Age only | 25-29 | 8.3% (8.0-8.6) | 1.74 (1.62-1.87) |
| Age only | 30-34 | 9.3% (9.0-9.7) | 1.84 (1.71-1.98) |
| Age only | 35-39 | 8.3% (8.0-8.6) | 1.71 (1.60-1.83) |
| Age only | 40-44 | 7.6% (7.3-7.9) | 1.66 (1.56-1.77) |
| Age only | 45-49 | 6.2% (5.9-6.4) | 1.33 (1.25-1.42) |
| Age only | 50-54 | 5.9% (5.6-6.2) | 1.17 (1.09-1.24) |
| Age only | 55-59 | 4.8% (4.5-5.1) | 0.98 (0.92-1.04) |
| Age only | 60-64 | 4.2% (4.0-4.5) | 1.00 (1.00-1.00) |
| Age only | 65-69 | 3.1% (2.9-3.3) | 0.87 (0.80-0.93) |
| Age only | 70-74 | 2.8% (2.6-3.0) | 0.76 (0.70-0.83) |
| Age only | 75+ | 4.5% (4.2-4.9) | 0.72 (0.66-0.79) |
| Ethnicity only | Asian | 8.5% (7.9-9.1) | 0.98 (0.90-1.06) |
| Ethnicity only | Black | 8.7% (7.9-9.5) | 2.29 (2.06-2.55) |
| Ethnicity only | Mixed/Other | 6.3% (5.7-7.0) | 1.35 (1.21-1.52) |
| Ethnicity only | White | 76.5% (75.2-77.9) | 1.00 (1.00-1.00) |
| NS-SeC only | 1 | 7.6% (7.3-7.9) | 1.00 (1.00-1.00) |
| NS-SeC only | 2 | 15.1% (14.6-15.6) | 1.36 (1.29-1.44) |
| NS-SeC only | 3 | 7.0% (6.6-7.3) | 1.16 (1.09-1.24) |
| NS-SeC only | 4 | 5.0% (4.6-5.4) | 0.85 (0.78-0.93) |
| NS-SeC only | 5 | 2.3% (2.1-2.6) | 0.81 (0.72-0.91) |
| NS-SeC only | 6 | 8.9% (8.4-9.4) | 1.52 (1.41-1.64) |
| NS-SeC only | 7 | 6.7% (6.2-7.1) | 1.07 (0.99-1.16) |
| NS-SeC only | Retired | 9.1% (8.6-9.6) | 0.64 (0.60-0.69) |
| NS-SeC only | Student | 4.9% (4.3-5.6) | 2.17 (1.90-2.49) |
| NS-SeC only | Under 17 | 27.8% (26.7-28.9) | 1.42 (1.33-1.51) |
| NS-SeC only | Unemployed | 5.7% (5.4-6.0) | 1.60 (1.50-1.71) |
| Gender only | Female | 53.8% (53.2-54.4) | 1.00 (1.00-1.00) |
| Gender only | Male | 46.2% (45.6-46.8) | 0.89 (0.87-0.92) |

**Table S9:** One-way stratified analysis results showing projected infection share and risk ratios by demographic group. Risk ratios are calculated relative to reference groups (60-64 age group, White ethnicity, NS-SeC 1, Female gender). Values are means with 95% confidence intervals from 1000 bootstrap simulations.
